# Automatic screening and characterization of patients with acquired neurological conditions from language

**DOI:** 10.1101/2025.08.28.25334096

**Authors:** Charalambos Themistocleous, Brielle Stark

## Abstract

Individuals with left-hemisphere damage (LHD), right-hemisphere damage (RHD), dementia, mild cognitive impairment (MCI), traumatic brain injury (TBI), and healthy controls are characterized by overlapping clinical profiles affecting communication and social interaction. Language provides a rich, non-invasive window into neurological health, yet objective and scalable methods to automatically differentiate between conditions with are lacking. This method aims to develop comprehensive neurolinguistic measures of these conditions, develop a machine learning multiclass screening and language assessment model (NeuroScreen) and offer a large comparative database of measures for other studies to build upon. We combined one of the largest databases, comprising 291 linguistic biomarkers calculated from speech samples produced by 1,394 participants: 536 individuals with aphasia secondary to LHD, 193 individuals with dementia, 107 individuals with MCI, 38 individuals with RHD, 58 individuals with TBI, and 498 Healthy Controls. Employing natural language processing (NLP) via the Open Brain AI platform (http://openbrainai.com), we extracted multiple linguistic features from the speech samples, including readability, lexical richness, phonology, morphology, syntax, and semantics. A Deep Neural Network architecture (DNN) classifies these conditions from linguistic features with high accuracy (up to 91%). A linear mixed-effects model approach was employed to determine the biomarkers of the neurological conditions, revealing distinct, quantitative neurolinguistic properties: LHD and TBI show widespread deficits in syntax and phonology; MCI is characterized by fine-grained simplification; patients with dementia present with specific lexico-semantic impairments; and RHD shows the most preserved profile. Ultimately, the outcomes provide an automatic detection and classification model of key neurological conditions affecting language, along with a novel set of validated neurological markers for facilitating differential diagnosis, remote monitoring, and personalized neurological care.

## 1 Introduction

Language is a distinctively human cognitive system that enables individuals to communicate, share information, and socialize. It includes a complex interplay of spoken, written, and signed modalities, drawing on multiple linguistic subsystems, including phonology (the sound structure of words), morphology (the internal structure of words), syntax (rules governing sentence structure), semantics (meaning), and pragmatics (the social use of language) (Hornstein et al., 2005; Levinson, 1983). Even simple tasks, such as ordering a meal, rely on the integration of these linguistic processes. Language is not only central to social participation but is also tightly linked to broader cognitive functions, including memory, attention, and executive functioning (Krakauer & Carmichael, 2017; Poeppel et al., 2008). Consequently, when language is disrupted due to neurological conditions such as left hemisphere damage (LHD), right hemisphere damage (LHD), dementia, mild cognitive impairment (MCI), or traumatic brain injury (TBI), the consequences extend beyond isolated cognitive deficits to independence, social participation, and overall quality of life. Yet, despite the critical role of language in human functioning, assessing and monitoring language functioning in clinical practice and properly treating it remains challenging.

The distinct underlying pathologies of LHD, RHD, dementia, MCI, and TBI produce unique behavioral profiles by differentially affecting receptive and expressive language (Goodglass & Kaplan, 1979; Lezak, 1995) (Table 1). These can serve as early linguistic markers that characterize these patients (Obler et al., 1991). Neurological research has shown that LHD primarily impacts language and other cognitive functions (Fridriksson et al., 2018; Hillis et al., 2018; Sebastian et al., 2018). Right hemisphere stroke can impair spatial awareness, emotions, and nonverbal and pragmatic communication (Joanette et al., 2015; Minga et al., 2023; Riès et al., 2016; Stockbridge, Sheppard, et al., 2021; Turkeltaub, 2015). Both LHD and RHD can language deficits, but the specific nature of these deficits differs (Caplan, 1987; Goodglass, 1993; Patel et al., 2018; Sidtis & Yang, 2017). MCI, an early cognitive decline, is typically amnestic in nature (affecting memory), but also typically impacts language and other critical cognitive domains, such as attention, and executive functions (Kim et al., 2024; Lyketsos et al., 2002; Nordlund et al., 2005; Park et al., 2011; Petersen et al., 1999; Petersen et al., 2001). Dementia is a progressive deterioration of the brain health due to neurodegeneration, affecting multiple cognitive domains, such as memory, language, attention, and movement (Faroqi-Shah et al., 2020; Mesulam, 1982; Tsapkini et al., 2018). TBI is a heterogeneous disorder, resulting in open or closed head trauma by an external force, such as a blow to the head, a fall, a car accident, and a penetrating injury. It can range from mild (e.g., concussion) to severe, with varying degrees of physical, cognitive, emotional, and behavioral effects (Birch et al., 2024).

**Table 1.**
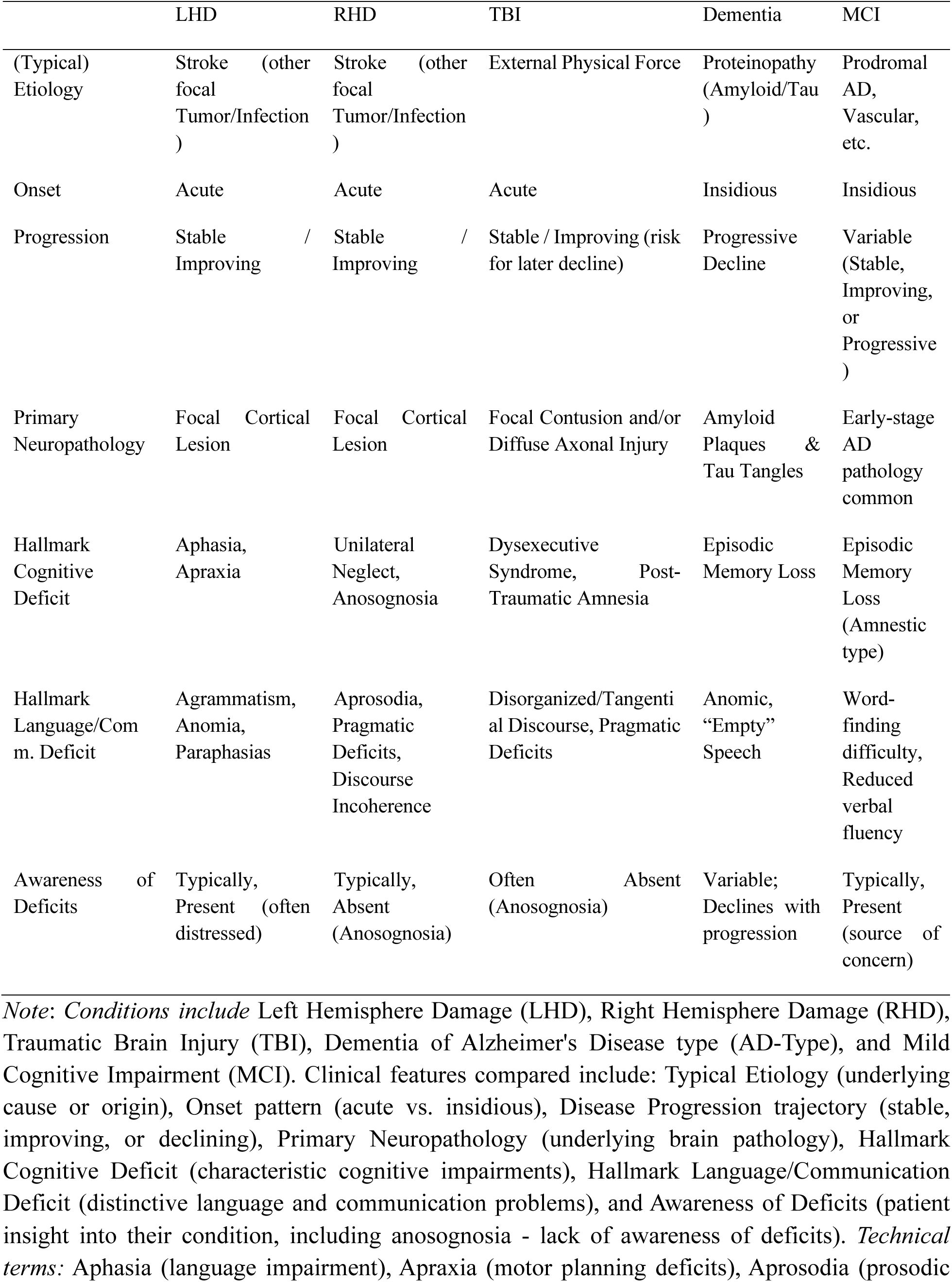
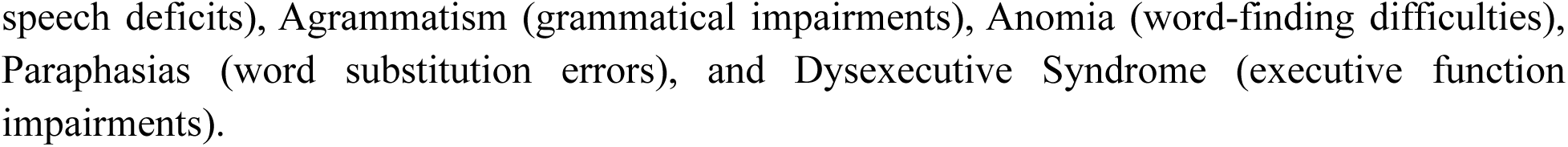
Comparative Table of Neurocognitive Conditions. Comprehensive comparison of five major neurocognitive conditions across key clinical and neurological characteristics. This comparative framework facilitates differential diagnosis and understanding of the distinct neuropsychological profiles associated with each condition.

### Conventional language assessment tools

Conventional language assessment tools, including structured tasks (e.g., Philadelphia Naming Test (Roach et al., 1996), Boston Naming Test (Kaplan et al., 2001), standardized batteries Western aphasia battery (revised) (WAB-R) (Kertesz, 2006), Quick Aphasia Battery (QAB) (Wilson et al., 2018) and the Boston Diagnostic Aphasia Examination (BDAE) (Goodglass et al., 2001), and patient– and clinician-rated evaluations (Frattali et al., 1995; Holland et al., 2018; Swinburn et al., 2019), are widely used to support diagnosis and guide treatment decisions. These methods often provide a narrow window into specific abilities like object naming, overlooking the multidimensional nature of everyday communication. Furthermore, their time-intensive and stressful nature makes them ill-suited for widespread screening. Clinicians may instead use general neurocognitive screeners like the MoCA or MMSE (Ciesielska et al., 2016; Davis et al., 2021; Tombaugh & McIntyre, 1992), but these still require in-person assessment and may not be sensitive enough to detect subtle language impairments characteristic of conditions like mild cognitive impairment (MCI). This creates a critical challenge for early detection and prognosis. A powerful solution lies in combining large-scale language corpora with computational methods such as Natural Language Processing (NLP) and Machine Learning (ML). This approach can enable automated screening and provide a deeper, comparative understanding of these conditions against each other and normative data from healthy individuals.

We address a critical limitation in neurolinguistics—the tendency to study conditions in isolation—by creating a unified analytical framework. Analyzing connected speech and discourse—how individuals use language in natural, extended communication—is widely regarded by researchers and clinicians as a best practice for assessing language abilities (Mueller et al., 2018). This approach captures real-world communicative competence and can reveal subtle linguistic deficits that standardized, isolated tasks often overlook. However, despite its advantages, discourse analysis remains underused in clinical practice due to its time-intensive nature, lack of scalable and standardized tools, and the manual effort required for transcription and coding (Stark et al., 2021). Moreover, traditional assessments are typically conducted in controlled clinical environments, which may not reflect an individual’s everyday communication patterns, thereby limiting ecological validity. Consequently, subtle or early-stage language impairments— especially those associated with heterogeneous conditions such as mild cognitive impairment (MCI) or early dementia—often go undetected until more pronounced cognitive or functional decline is evident.

Recent advances in artificial intelligence (AI), NLP, ML, and automated speech analysis have opened new possibilities for addressing these limitations. By leveraging AI-driven approaches to extract and quantify linguistic features from spontaneous speech, we and others have demonstrated their potential for objective, reproducible, and ecologically valid measures of language production from transcripts or audio files (Kim et al., 2024; Themistocleous et al., 2018; Themistocleous, Ficek, et al., 2020; Themistocleous, Webster, et al., 2020). These computational methods can quantify automatically domains of language disorder—spanning lexical diversity (Fergadiotis & and Wright, 2011), phonological structure (Barbieri et al., 2018; Croot et al., 2000), morphological patterns (Badecker et al., 1990; Caramazza & Hillis, 1991; Fridriksson et al., 2018; Hillis, 1989; Hillis et al., 2018; Stockbridge, Matchin, et al., 2021), syntactic complexity (Bastiaanse, 2013; Caramazza & Hillis, 1989; Mack et al., 2021; Thompson & Mack, 2014; Wilson et al., 2016), semantic content, and readability (Dale & Chall, 1948; Fitzsimmons et al., 2010; Klare, 1974; Themistocleous, 2024)—represent a promising class of digital biomarkers with the potential to support early detection (Fraser et al., 2018; Themistocleous et al., 2018; Themistocleous, Eckerström, et al., 2020), differential diagnosis (Fraser et al., 2019; Kim et al., 2024; König et al., 2015; Stark et al., 2025; Themistocleous, Ficek, et al., 2020), and ongoing monitoring of neurological conditions (Ahmed et al., 2013; Lavoie et al., 2023; Tuomiranta et al., 2025). Despite that these studies demonstrates that automated language analysis holds significant promise as a digital health tool, several challenges must be addressed before it can be fully integrated into clinical practice.

Automated language analysis is progressively recognized as a digital health tool (Beltrami et al., 2018; Fraser et al., 2015), yet its clinical translation is constrained by several critical gaps. For these computational tools to improve patient outcomes in a meaningful way, they must first move beyond the current landscape of proof-of-concept studies, which often use small, homogenous datasets from isolated clinical populations but rely on rigorous validation across large, diverse, and multi-condition populations is essential. This validation must also establish robust normative data from healthy controls, enabling clinicians to benchmark an individual’s performance to accurately assess pathology and severity. Also, the development of sophisticated computational pipelines must be paired with a focus on practical application: creating scalable, automated, and openly accessible tools that can integrate seamlessly into clinical workflows to reduce clinician burden and enhance diagnostic precision. Addressing these interconnected challenges is the essential next step toward realizing language as a clinically actionable digital biomarker.

### Study Aims

This study has an overarching aim to advance a novel paradigm for neurological assessment to corroborate existing neurological assessments and to establish spoken language as a scalable and clinical digital biomarker by evaluate a comprehensive set of measures from the key linguistic domains, readability, phonology, morphology, syntax, semantics, and lexicon (Supplementary Data 1 offers a detailed description see also the Methods section).

This provides a two-fold aim. The first aim is to develop a multi-class machine learning approach for neurological screening (NeuroScreen) that can discriminate patients from HCs and the subtype individual patient subgroups from each other. Ultimately, the MLs aim to answer two primary research questions (1) How well do the models distinguish patients and healthy controls? And (2) How well does the ML model distinguish each sub-group in the data? By answering these two questions, we will be able to determine how well the models can be employed in real-life scenarios for detecting patients and in the clinic to subtype patients, and which of them with high confidence. To achieve aim we have developed an end-to-end AI-driven procedure to analyze a large and diverse database of over 9,900 speech samples based on an end-to-end ML model that combines NLP pipelines that employ Open Brain AI (Themistocleous, 2024), a platform we have developed to extract the linguistic features. Subsequently, we preprocessed and standardized the calculated measures and passed them to a set of ML models, namely Random Forrest, Support Vector Machine, Logistic Regression, and Deep Neural Networks. These models were tuned through hyperparameter tuning and evaluated.

(2) The second aim is to provide explainable measures, namely the linguistic signatures of five major neurological conditions (LHD, RHD, dementia, MCI, and TBI). This is critical to understanding the effects of each condition on language and to providing therapeutic targets for novel clinical approaches. In other words, we will determine (1) Which linguistic measures differ most due to diagnostic groups? (2) Which are the distinctive features for each neurological condition compared to HC? And (3) What do language measures reveal for each patient group? To achieve this aim, we developed (generalized) linear mixed effect models while controlling for the effects of task and the participant.

This computational approach moves beyond prior research by leveraging ecologically valid data from everyday communicative tasks to create a comprehensive, multi-faceted portrait of how language changes in response to brain injury and disease, aiding in differential diagnosis, particularly for disorders with overlapping symptoms like MCI and early dementia, and offering a non-invasive, low burden means for monitoring disease progression and treatment response over time. Ultimately, this research contributes to the digital transformation of clinical practice by providing a validated set of open-access linguistic biomarkers, this study creates new opportunities for remote, low-burden monitoring of neurological health, supporting a future of more accessible, data-driven, and personalized care.

## 2 Methodology

### 2.1 Participants

The individuals for this study were drawn from Neural Databank collected and developed by the second author (Stark et al., 2023), now part of the Aphasia Bank, and data from the TalkBank consortium (https://talkbank.org), which following a similar protocol. Each clinical bank (e.g., AphasiaBank, RHDBank) has an established discourse protocol that elicits a variety of discourse genres (MacWhinney, 2025).

i. *Aphasia Bank:* The database contains spoken discourse samples from individuals with LHD and control participants, designed to study language production and its neural foundations. The research emphasizes connected speech (discourse) rather than single words or isolated sentences. Participants completed a full discourse protocol twice within a short timeframe to assess the test-retest reliability and stability of discourse measures. The participants contain both people with LHD (536 individuals) and HCs (359 individuals) (Stark et al., 2023).
ii. *Right Hemisphere Damage Bank (RHD Bank):* This is a specialized database focused on communication in individuals with RHD. The database serves as a resource for understanding and treating communication disorders following RHD, particularly focusing on pragmatic language abilities, discourse coherence, and real-world communication challenges (Minga et al., 2022). This bank includes 38 individuals with RHD and 40 Healthy Controls.
iii. *Traumatic Brain Injury (TBI Bank):* This is a multimedia database focused on studying communication disorders in individuals with TBI. TBIBank protocol includes discourse tasks such as the Cinderella story retell, following similar methodology to other TalkBank databases. The protocol consists of discourse genres including personal narratives, picture descriptions, story retelling, and procedural discourse. TBIBank is a longitudinal study in which brain injured people are videoed at 6 different time points post injury performing a uniform set of tasks, with the goal of identifying recovery patterns. The database enables automated language analysis, diagnostic profiling, comparative evaluation of treatment effects, and profiling of recovery patterns in TBI populations, supporting both research and clinical applications in understanding cognitive-communication disorders following brain injury. This bank includes 58 individuals with TBI.
iv. *Dementia Bank – Delaware MCI dataset:* This corpus is part of DementiaBank and includes language productions by 71 adults with MCI, from the Delaware Corpus and Baycrest Centre Corpus. This data contributes to early detection of subtle changes in language and cognition and provide insight into MCI subtypes based on discourse profiles (Lanzi Alyssa et al., 2023).
v. *Dementia Bank – Pitt Study (Pitt Study):* A comprehensive description of this dataset is provided in Becker et al. (1994).Briefly, the study includes a picture description task from the Boston Diagnostic Aphasia Examination (Goodglass & Kaplan, 1983), a widely used diagnostic tool for detecting language abnormalities. In this task, participants were shown the “Cookie Theft” picture stimulus and instructed to describe everything they observed. Their responses were audio-recorded and later transcribed verbatim. This study includes 193 individuals with Dementia and 99 Healthy Controls.

This study presents a comprehensive analysis of linguistic measures across various diagnostic groups by combining data from multiple discourse tasks, extending beyond the conventional Cookie Theft picture description. Our primary analysis provides a consolidated overview of these linguistic features (Table 2; Supplementary Data 2, provides a more comprehensive data breakdown of Data Count by Group, Project, and Task). Recognizing that different tasks may elicit distinct communication patterns, we have preemptively accounted for potential task-specific effects within our statistical models by adding the task in the random effects. To ensure full transparency and to allow for a more granular examination of these variations, we provide a detailed breakdown of the linguistic signatures for each task in the Supplementary Tables.

**Table 2.** Cognitive assessment tasks administered across diagnostic groups and research studies.

| Diagnosis | Task List |
| --- | --- |
| LHD | Cat, Cinderella, Flood, Important Event, Sandwich, Speech, Stroke, Umbrella, Window |
| MCI | Cookie Theft |
| HC | Cat, Cinderella, Cookie Theft, Flood, Illness, Important Event, Sandwich, Speech, Umbrella, Window |
| MCI | Cat, Cinderella, Cookie Theft, Rockwell, Sandwich, Umbrella, Window |
| Dementia | Cooke Theft |
| HC | Cookie Theft |
| RHS | Cat, Cinderella, Cookie Theft, Sandwich, Speech, Stroke |
| TBI | Brain Injury, Cat, Cinderella, Important Event, Recovery, Sandwich, Speech, Umbrella, Window |

Note: Cat = A description of a single picture, in which a cat is being rescued from a tree; Cinderella = retelling of the fictional narrative Cinderella, which is done after looking at a wordless picture book; Flood = A description of a single picture of a rescue during a flood; Important Event = a personal narrative about an important event; Sandwich = a procedural narrative describing how to make a peanut butter and jelly sandwich; Stroke or Recovery = A personal narrative about one's brain injury and recovery; Umbrella = A multiple scene picture sequence, in which a boy and mother interact about taking an umbrella into the rain; Window = A multiple scene picture sequence, in which a boy kicks a soccer ball through a man's window, shattering it; Cookie Theft = A description of a single picture, in which two kids steal a cookie; Rockwell = A description of a single picture, which is Norman Rockwell's "Coming and Going".

Participants participated in different tasks providing often more than one samples, the analysis is based on 9955 language samples drawn from multiple clinical databases produced by the individuals reported in

Table 3 (see also, Table 2 and Supplementary Data: Table 1). These databases exhibit significant clinical heterogeneity. For instance, the LHD database contains participant groups classified by subtype, including anomic, Wernicke’s, and Broca’s aphasia. The Pitt study’s dementia subgroup (N=193) further illustrates this diversity; it is composed primarily of patients with dementia (91%), who present with lower average Mini-Mental State Examination (MMSE) scores of 17–18, alongside individuals with MCI. We chose to incorporate these databases in their entirety for several reasons. This approach maintains the ecological validity of the data, ensuring our findings reflect the natural heterogeneity of clinical populations. Furthermore, it preserves the integrity of these standard corpora, which is crucial for the reproducibility and comparability of our results within the wider research community.

**Table 3.**
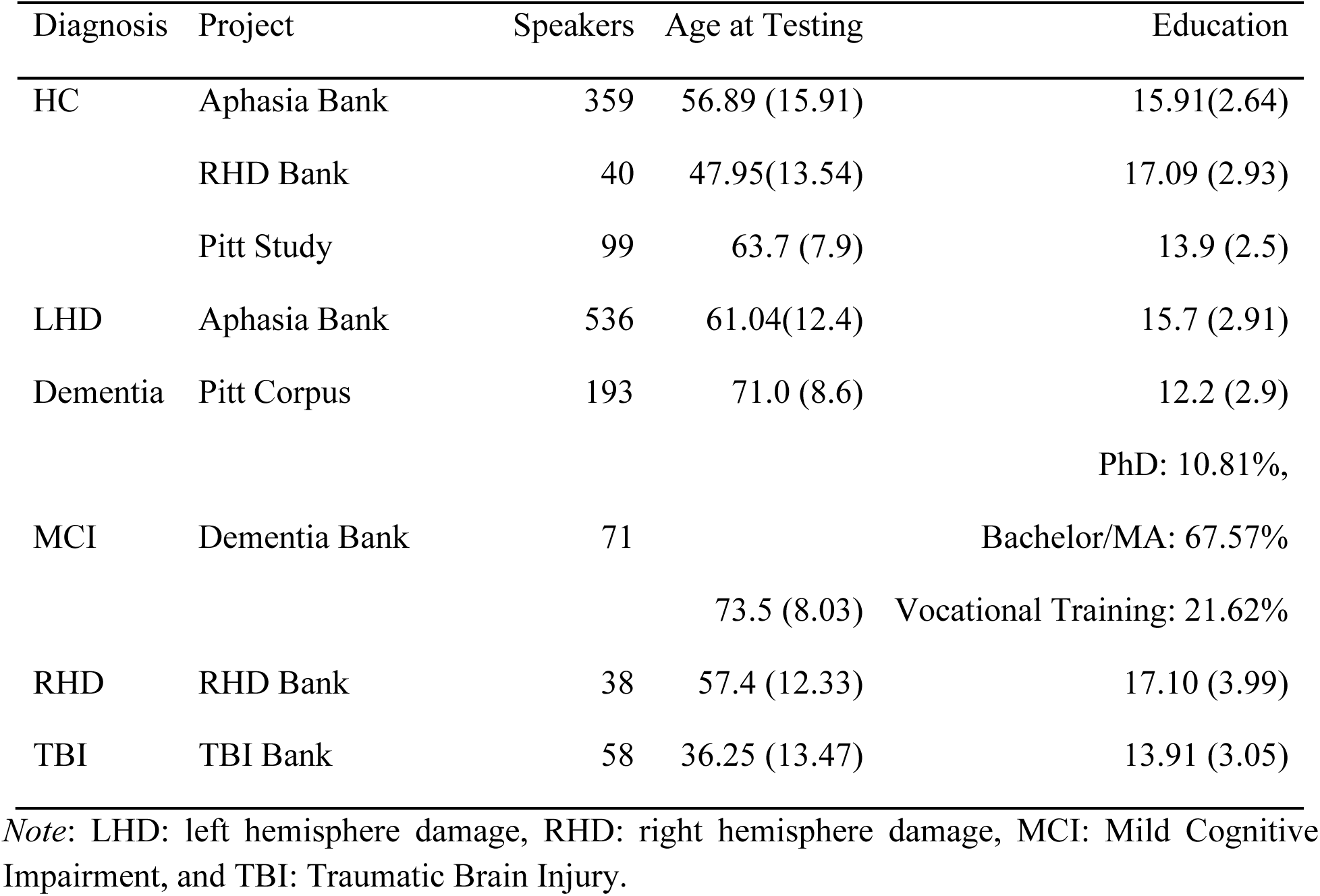
Participant demographics across diagnostic groups and research databases. The table presents sample sizes, mean ages, and educational attainment for participants in each diagnostic group across different research corpora. Age is reported as mean years (standard deviation). Education is reported as mean years of formal education (standard deviation) except for the MCI group where educational categories are presented as percentages.

### 2.2 Measures

Speech samples were analyzed using *Open Brain AI* (http://openbrainai.com; Figure 1), a custom clinical linguistics platform developed by the first author (Themistocleous, 2024) to facilitate automatic audio and linguistic analysis of texts. Unlike generic computational models, *Open Brain AI* was designed specifically for phenotyping of language features through a clinical lens, enabling hypothesis-driven research into speech pathology and neurogenic communication disorders. The platform calculates linguistic metrics in real-time as participants type or as clinicians transcribe speech samples, enabling immediate quantitative analysis of discourse features relevant to neurological conditions. Additional analysis modules accessible via the toolbar include syntactic complexity measures, semantic density calculations, and comparative normative data. This example demonstrates the platform’s capability to automatically extract objective linguistic measures from naturalistic discourse samples, facilitating evidence-based assessment of communication disorders across various neurological populations. *Open Brain AI* executed a cascade of NLP techniques. Core NLP steps included tokenization (segmenting text into individual words or tokens), part-of-speech tagging (assigning a grammatical category to each token), and dependency parsing (identifying the grammatical relationships between words and the syntactic structure of sentences). For each extracted feature, both raw counts and ratios (to normalize for variations in text length) are computed. These quantitative linguistic data were automatically exported by our computational platform as spreadsheet files, ready for statistical analysis.

**Figure 1.**
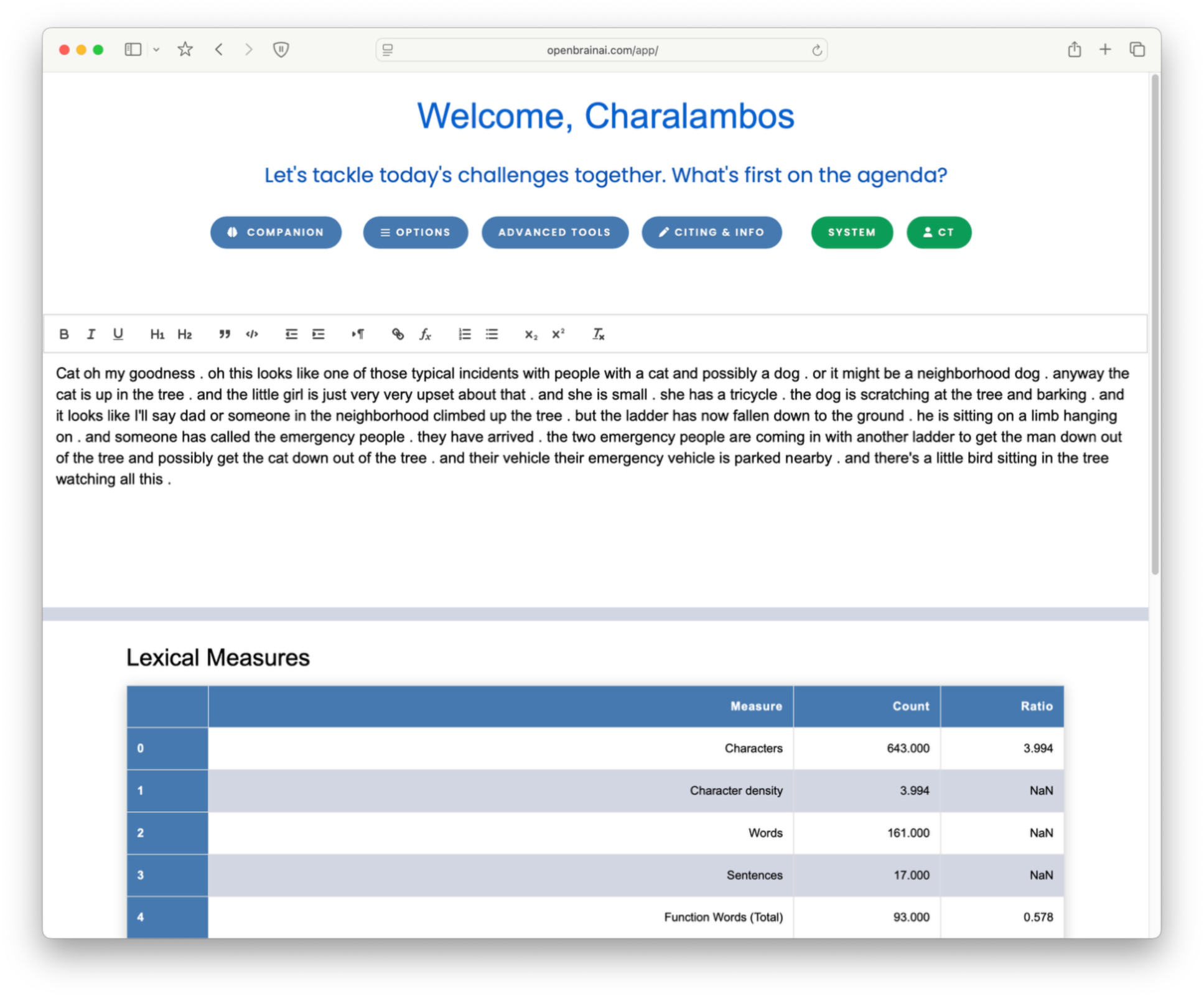
User interface of the Open Brain AI text analysis platform (Themistocleous, 2024) for neuropsychological assessment. The web-based platform provides real-time linguistic analysis of narrative discourse samples. The interface displays a text editor (top panel) containing a participant’s narrative description of the “Cat Rescue” picture stimulus, commonly used in aphasia and cognitive assessment batteries. The lower panel shows automated lexical measures including character count (643), word count (161), sentence count (17), and function word analysis (93 total function words, ratio 0.578).

From these foundational analyses, a comprehensive suite of linguistic measures was automatically extracted, quantifying aspects of (Supplementary Data 1 offers a detailed list of the measures):

i. *Readability.* Readability of text productions in patients with is a measure that has been evaluated for the first time concerning all these conditions in this study. Metrics assessing text complexity and perceived ease of understanding for a reader. *Readability* metrics include the Flesch-Kincaid Readability Tests, Gunning Fog Index, and SMOG Index (Dale & Chall, 1948; Fitzsimmons et al., 2010; Klare, 1974; Themistocleous, 2024) quantify how easy a text can be to be read and understood by a reader. It is typically influenced by factors such as sentence length, word complexity, and the overall structure of the text. Overall, we expect that patient speech should be simpler and more readable than that of healthy individuals.
ii. *Lexicon and Lexical Information.* We have designed features related to the vocabulary richness, diversity, and usage within the text. This includes measures like Type-Token Ratio, counts of content versus function words, and average word length. These measures explain the distribution of words and relationships between types and tokens that can quantify how words are used in different contexts and how they contribute to the overall meaning of a text such as lexical diversity measures (Fergadiotis & and Wright, 2011).
i. *Phonology*. Characteristics of sound structure, such as counts of words by syllable Number: (e.g., one-syllable, two-syllable words) and the distribution of various Consonant (C) and Vowel (V) syllable structures (e.g., CV, CVC, CCVC). We designed these measures to quantify how users employ speech sounds, the sound combinations, and the complexity of syllables. Comparing these measures across patients with different language impairments can reveal characteristics that pertain to the effects of impairment on the cognitive representation of sounds and speech production (Barbieri et al., 2018; Croot et al., 2000).
ii. *Morphology.* Analysis of word structure, encompassing both the distribution of parts of speech (e.g., counts and ratios of nouns, verbs, adjectives, and adverbs) and inflectional categories (e.g., tense, Number: Gender: case). Morphological measures quantify the structure and form of words, the distribution of parts of speech, and inflectional categories, such as tense, Number, Gender, and Case. Comparing patients with morphology impairments can reveal pathologies, like agrammatism and anomia (Badecker et al., 1990; Caramazza & Hillis, 1991; Fridriksson et al., 2018; Hillis, 1989; Hillis et al., 2018; Stockbridge, Matchin, et al., 2021).
ii. *Syntax.* Measures of sentence structure and grammatical complexity. This included quantification of various phrase types (e.g., Noun Phrases, Verb Phrases, Prepositional Phrases), analysis of core syntactic dependencies and relations (e.g., nominal subjects, direct objects, adverbial clause modifiers), and overall sentence complexity metrics (e.g., Average Sentence Length, T-units, and syntactic tree depth/Yngve load). These measures quantify impairments of sentence structure (e.g., subject-verb-object order), grammatical rules (e.g., agreement between subject and verb), and phrase structure (e.g., noun phrases, verb phrases) (Bastiaanse, 2013; Caramazza & Hillis, 1989; Mack et al., 2021; Thompson & Mack, 2014; Wilson et al., 2016).
iii. *Semantics.* Primarily focused on Named Entity Recognition (NER), which involves identifying and categorizing named entities in text into predefined classes such as persons, organizations, locations, dates, and quantities.

These grammatical analyses utilized the *Universal Dependencies* framework for standardized annotation (Nivre et al., 2020) and custom made metrics, which were systematically selected using both established measures based on established theoretical frameworks in clinical linguistics and their demonstrated sensitivity to pathological language changes in neurogenic communication disorders (like counts of nouns and verbs) and novel measures that aim to encompass microstructural elements (phonology, morphology), macrostructural components (syntax, semantics), and pragmatic dimensions.

Thus, these measures aim to provide a comprehensive characterization of language impairments that aligns with current models of linguistic breakdown in clinical populations. By capturing this full spectrum of linguistic variation, the analysis framework enables detection of subtle but clinically significant changes that might be overlooked by assessments targeting only isolated linguistic domains. A complete list of all measures and their detailed operational definitions is provided in Supplementary Data 1. Given this large feature set, the analyses presented in this paper prioritize a subset of measures selected for their demonstrated high sensitivity and specificity in distinguishing between the diagnostic groups (LHD, Dementia, MCI, RHS, TBI) and Healthy Controls, as well as differentiating the clinical groups from one another. An exhaustive output of all statistical results for every measure is available in the Supplementary Materials.

### 2.3 Machine Learning Pipelines

**Figure 2.**
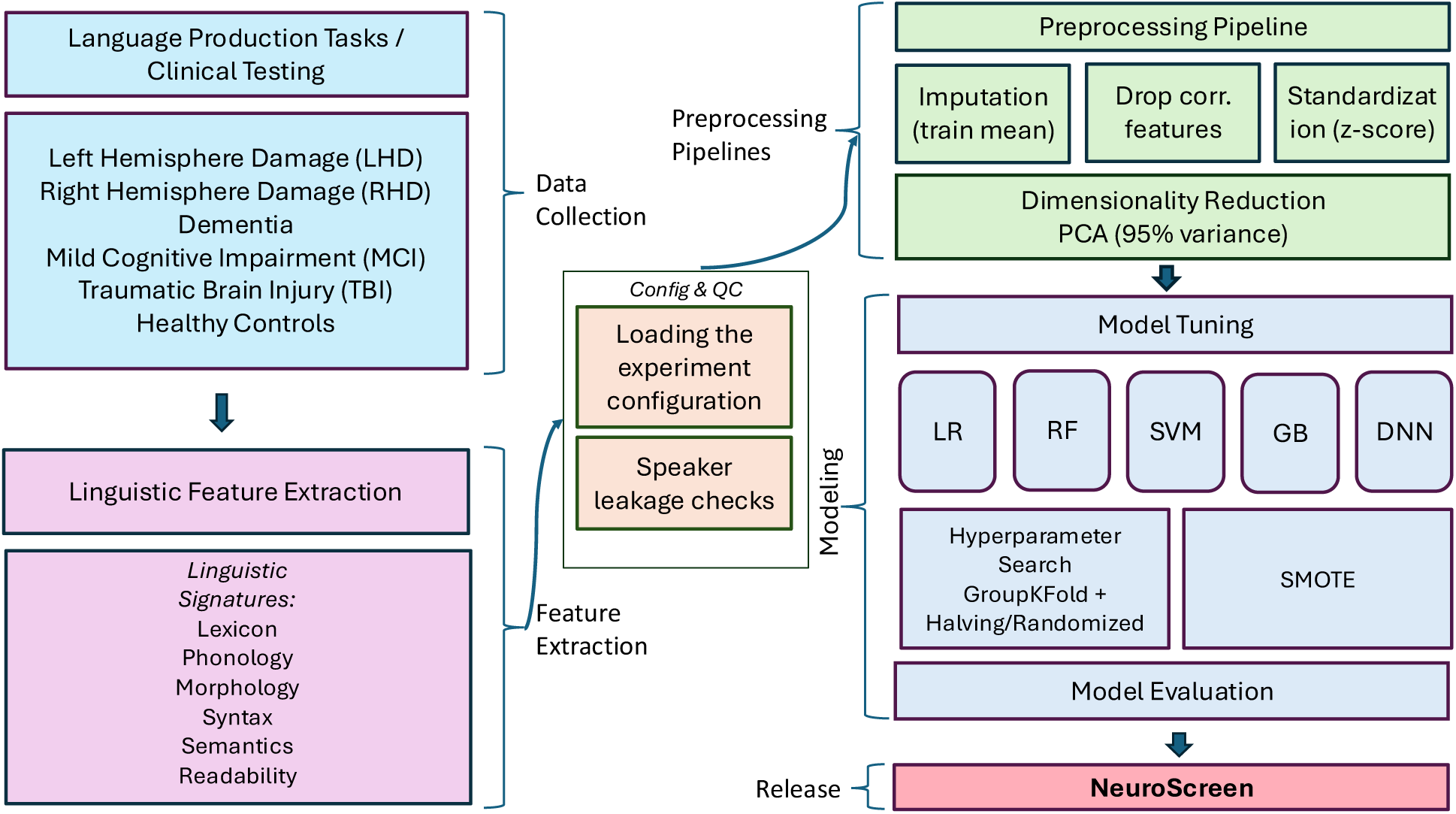
NeuroScreen machine learning pipeline architecture for automated neurological assessment. The comprehensive workflow shows the development and validation of a diagnostic system that analyzes language production to distinguish between neurological conditions. Input data comprises speech and text samples from participants across six diagnostic groups: Left Hemisphere Damage (LHD), Right Hemisphere Damage (RHD), Dementia, Mild Cognitive Impairment (MCI), Traumatic Brain Injury (TBI), and Healthy Controls. Language production tasks undergo automated linguistic feature extraction across six domains: Lexicon (vocabulary richness), Phonology (speech sound patterns), Morphology (word formation), Syntax (grammatical structure), Semantics (meaning content), and Readability (text complexity). The preprocessing pipeline includes quality control checks, speaker leakage detection, correlated feature removal, mean imputation for missing values, *z*-score standardization, and principal component analysis for dimensionality reduction (retaining 95% variance). Five machine learning algorithms are systematically evaluated: Logistic Regression (LR), Random Forest (RF), Support Vector Machine (SVM), Gradient Boosting (GB), and Deep Neural Network (DNN). Model optimization employs hyperparameter tuning with GroupKFold cross-validation and randomized/halving grid search. Synthetic Minority Oversampling Technique (SMOTE) addresses class imbalance. The validated models comprise the NeuroScreen diagnostic tool for objective, automated neurological assessment based on quantitative linguistic analysis.

We designed a machine learning pipeline to classify a speaker’s diagnosis into one of six categories based on statistical features derived from language productions on tasks, namely patient with LHD, RHD, dementia, MCI, TBI, and HCs. The pipeline is designed to manage speaker-dependent data, address class imbalance, and a provide comprehensive, comparative evaluation of multiple machine learning (ML) models, namely include Random Forest, Support Vector Machine (SVM), Logistic Regression, Gradient Boosting, and a Deep Neural Network (DNN). The entire process, from data preparation to model evaluation, was conducted in a Python environment utilizing pandas for data manipulation (McKinney, 2010), scikit-learn (Pedregosa et al., 2011) and imbalanced-learn (Lemaître et al., 2017) for machine learning algorithms. The deep learning component was build using Tensorflow (Abadi et al., 2016).

## 1. Data Preparation and Cohort Definition

The core of our methodology is built upon the principle of speaker-independent validation, which is crucial for developing models that can generalize to new, unseen individuals rather than memorizing characteristics of specific speakers in the training set. To facilitate this, a designated speaker identifier column was used to group data points belonging to the same individual. The dataset was then partitioned into features and the target variable.

To ensure that the model evaluation provides a realistic estimate of performance on new individuals, a strict speaker-independent splitting protocol was enforced. The dataset was divided randomly into a training set (80%) and a hold-out test set (20%) using the GroupShuffleSplit strategy. This method guarantees that all data points from any given speaker are confined to only one of the sets (either training or testing), completely preventing data leakage between them. This approach is critical for assessing the model’s ability to generalize beyond the specific speakers it was trained on.

## 3. Preprocessing and Feature Engineering Pipeline

A multi-step preprocessing pipeline was applied sequentially to the data. Crucially, all preprocessing steps were fitted only on the training data to prevent information from the test set from influencing the training process. The same fitted transformers were then used to transform both the training and test sets.

i. Missing values in the feature set were managed by imputing them with the mean of their respective columns, calculated from the training data.
ii. To reduce multicollinearity and model complexity, highly correlated features were removed. A Pearson correlation matrix was computed on the training set, and for any pair of features with a correlation coefficient and we evaluated various threshold features, for correlations greater than 0.90, one of the features was discarded.
iii. The features were standardized by removing the mean and scaling to unit variance using the StandardScaler (Pedregosa et al., 2011). This transformation ensures that features with larger scales do not disproportionately influence model training, which is particularly important for distance-based algorithms like SVM and regularization models like Logistic Regression.
iv. Principal Component Analysis (PCA) was employed as the final feature engineering step. PCA transforms the standardized features into a smaller set of uncorrelated principal components. The number of components was chosen to retain 95% of the original variance in the training data, effectively reducing noise and the dimensionality of the feature space while preserving most of the relevant information.

## 4. Model Training, Imbalance Handling, and Hyperparameter Optimization

We have evaluated five distinct classification models to explore a range of algorithmic approaches: Logistic Regression (LG), Random Forest, Support Vector Machine (SVM) with an RBF kernel, Gradient Boosting, and a feedforward Deep Neural Network (DNN). We selected these models to allow for a comprehensive analysis of the dataset and selection of a model that explain the data. More specifically, the following models were selected:

1. LG is a fundamental linear classification algorithm. It works by fitting a linear equation to the features and then applying a logistic function (or sigmoid function) to the output to return a probability between 0 and 1. This probability is then used to predict the class. LG serves as a baseline model (Hastie et al., 2009).
2. RFs is an ensemble learning method; it constructs many individual decision trees during training. It can capture complex, non-linear relationships in the data without requiring explicit transformations. It is generally robust to overfitting, especially when compared to a single decision tree as it averages the predictions of many trees (Breiman, 2001).
3. SVM models detect the optimal hyperplane (or decision boundary) that best separates the classes in the feature space. SVM can model both linear and non-linear boundary by mapping the data into a higher-dimensional space, with good generalization performance on unseen data (Cortes & Vapnik, 1995).
4. GB is another powerful ensemble technique like the RFs, which builds models sequentially. It starts with a simple model and then iteratively adds new decision trees that are specifically trained to correct the errors made by the previous ones. RFs, however, build trees independently and in parallel whereas GBs are sequential with an error-correcting approach leading to more powerful and flexible model (Hastie et al., 2009).
5. DNN consists of an input layer, multiple “hidden” layers of interconnected nodes (neurons), and an output layer. The network learns to detect complex patterns and features by adjusting the connection weights between neurons during training. The DNN approach can uncover patterns in the data than the other, more traditional machine learning models might miss (Themistocleous, 2019).

The data exhibited an imbalanced class distribution as there are fewer patients with MCI, RHD, and TBI, than patients with dementia, LHD, and HC. To mitigate the risk of models becoming biased towards the majority class, we integrated the SMOTE directly into our modeling pipeline (Chawla et al., 2002). For each model, a pipeline was constructed with SMOTE as the initial step. This approach ensures that over-sampling is performed correctly within each cross-validation fold: SMOTE is fitted and applied only to the training data partition of a fold, generating synthetic samples for the minority classes before the classifier is trained. The validation partition of the fold remains in its original, imbalanced state, providing an unbiased evaluation of the model’s performance. This in-pipeline application of SMOTE is crucial for preventing data leakage and obtaining a reliable estimate of model generalizability. We defined a custom DynamicSMOTE class to automatically adjust the *k* neighbors parameter, preventing errors in cross-validation folds where a minority class had very few samples.

To identify the optimal set of hyperparameters for each model, we employed a hybrid search strategy using GroupKFold cross-validation (with 5 folds) to maintain speaker independence. For the traditional models (Logistic Regression, Random Forest, SVM, Gradient Boosting), we used HalvingRandomSearchCV. This efficient method starts by evaluating many hyperparameter combinations on a small subset of the data and iteratively prunes fewer promising candidates, allocating more resources to the best-performing ones.

For the computationally intensive Deep Neural Network (DNN), we used RandomizedSearchCV to sample a fixed number of hyperparameter combinations from the search space. The performance of each combination was evaluated based on its default scoring metric. The best hyperparameters for SMOTE’s k neighbors parameter were also determined during this search. The DNN architecture was also part of the hyperparameter search. Key parameters tuned included the number of hidden layers, the number of neurons, the dropout rate, batch size, and the learning rate for the Adam optimizer. An “early stopping callback” was used to prevent overfitting by halting training when performance on the loss function stopped improving.

## 6. Model Evaluation

After hyperparameter tuning, the best-performing version of each model was evaluated on the completely unseen hold-out test set. Model performance was assessed using a comprehensive set of metrics to provide a holistic view of their classification capabilities:

1) Accuracy is the percentage of predictions that were correct out of all predictions made. If your model correctly predicts 85 out of 100 cases, your accuracy is 85%.
2) Balanced Accuracy solves this problem by averaging the accuracy within each class. It calculates the recall (true positive rate) for each class separately, then takes the average. In other words, the balanced accuracy is defined as the average of sensitivity (true-positive rate) and specificity (true-negative rate) for the two classes in a binary classification “Patient vs. Healthy Control (HC)”, the Specificity (HC Recall) (1) and the Sensitivity (Patient Recall) (2) is calculated. Then the Balanced Accuracy is the sum of the Specificity and Sensitivity divided by two (2), the number of classes in a binary classification.

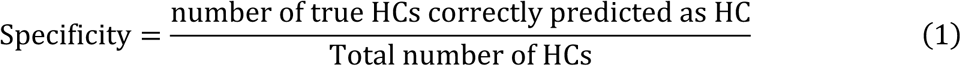

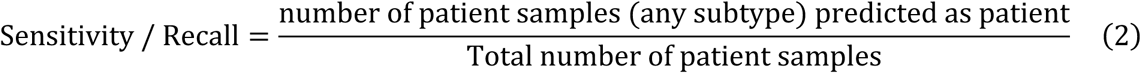
3) F1-Score (Weighted) addresses the trade-off between recall (2) and precision (3). The F1-score is the harmonic mean of these two, giving you a single number that balances both concerns. The weighted version calculates F1-scores for each class and then averages them based on how many samples each class has, making it appropriate for imbalanced datasets.

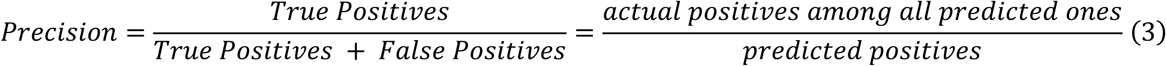
4) Cohen’s Kappa measures how much better your model performs compared to random chance. It is particularly valuable because it accounts for the possibility that some correct predictions might just be lucky guesses. Kappa values range from –1 to 1: 1.0: Perfect agreement beyond chance and 0.0: Agreement is no better than random chance; Negative values mean worse than random chance
5) AUC-ROC (Area Under the Receiver Operating Characteristic Curve). The ROC curve plots your model’s true positive rate against its false positive rate across all possible classification thresholds. The AUC-ROC tells you how well your model can distinguish between classes. AUC = 1.0: Perfect classifier
6) AUC-PR (Area Under the Precision-Recall Curve). ROC curves can often be optimistic on imbalanced datasets, precision-recall curves focus specifically on the positive class performance. This makes AUC-PR especially valuable when you care more about correctly identifying the minority class. The PR curve plots precision against recall at different thresholds. AUC-PR is particularly informative for imbalanced data.
7) Confusion matrices were generated for each model to visualize the distribution of correct and incorrect predictions across the different classes. For tree-based models (Random Forest, Gradient Boosting), feature importance scores were calculated and visualized to provide insights into the most influential principal components for classification. Finally, the best overall model, along with the fitted preprocessing transformers, was saved for potential future deployment.

### 2.4 Statistics

To assess the influence of clinical diagnosis on each linguistic outcome variable, we utilized an automated mixed-effects modeling pipeline. This analysis included participants from the five diagnostic groups (LHD, Dementia, MCI, RHS, TBI) and the Healthy Control (HC) group. The pipeline, developed in R (R Core Team, 2025) was designed to be flexible, data-driven, and robust to violations of statistical assumptions common in linguistic data.

For each linguistic variable, a mixed-effects model was implemented. *Diagnosis* was specified as a fixed effect to determine its influence on the outcome.

As discussed earlier there is variation in the subgroups within the participants and the tasks they perform, to appropriately account for the non-independence of data arising from the study design, and given the complexity of the databases, two random intercepts were included in the model:

1. The (1 | Speaker) term addresses that multiple observations (i.e., linguistic measures from one or more tasks) originate from the same individual. By including a random intercept for each speaker, the model accounts for individual-specific baseline differences in linguistic performance, thereby modeling the repeated measures dimension of the data.
2. The (1 | Task) term addresses the inherent variability across different elicitation tasks (e.g., “Cinderella,” “Flood,” and “Cookie Theft,” as listed in Table 1). Given that the study design involved diverse groups of participants undertaking varying subsets of these tasks, this random intercept allows the model to estimate an average deviation from the overall mean for each specific task. This effectively controls for baseline differences in how tasks might elicit certain linguistic features, regardless of the speaker or their diagnosis.

These random effects structure is robust to the unbalanced nature of task administration (i.e., not all participants completed all tasks, and tasks were not fully crossed with participants). It allows for the estimation of the fixed effect of ‘Diagnosis’ while simultaneously partitioning out variance attributable to individual speakers and specific tasks. The general model structure was:

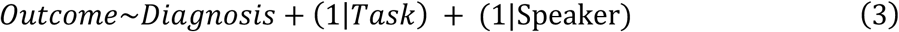

The analytical pipeline systematically selected the most appropriate statistical model based on the distribution of each dependent variable. This adaptive process involved fitting Gaussian Linear Mixed-Effects Models (LMMs) for continuous variables, using robust LMMs if residual diagnostics (via the *DHARMa* package (Hartig, 2016)) indicated violations of model assumptions, and employing Generalized Linear Mixed-Effects Models (GLMMs) with appropriate distributions (e.g., binomial, Poisson, or negative binomial) for binary or count data, including checks for overdispersion and zero-inflation. If a suitable model could not be fitted through these steps, a rank-based LMM was applied as a robust fallback. (Further details on the specific model selection criteria and R packages, such as *lmerTest* (Kuznetsova et al., 2016) and robustlmm (Koller, 2016).

When a significant main effect of ‘Diagnosis’ was found (typically p<.05), post-hoc pairwise comparisons were conducted between all diagnostic groups using estimated marginal means (via the *emmeans* package (Russell, 2020)). Tukey’s method was applied to adjust for multiple comparisons. Group means and confidence intervals are reported to aid in the interpretation of these differences.

To create a ranked list of linguistic signatures, a key statistic from the post-hoc analysis of your mixed-effects models. A larger z-ratio indicates a more robust and statistically significant difference. It simultaneously accounts for the size of the difference and the precision of the measurement. We use the absolute value of the z-ratio for ranking because we are interested in the *magnitude* of the difference, regardless of whether a feature’s value increased or decreased. This allows us to directly compare the most impactful features across all groups. The direction of the change (increase or decrease) is then indicated separately in the table with arrows.

## 3 Results

We examined the distinct linguistic production of each group on a comprehensive set of linguistic automated measures spanning lexical, morphological, phonological, readability, semantic, and syntactic domains.

### 3.1 How well do the models distinguish patients and healthy controls?

To assess how well the models distinguish patients and HCs, we have collapsed all five patient subtypes into one “Patient” group, and we can compute the results shown in Table 4. The plethora of available data for this classification enabled the models to perform exceptional well. LR is essentially perfect at flagging “Patient” vs. “HC” (balanced accuracy ≈ 99%). The DNN and the SVM both perform close to 95% thresholds; the RF and the GB (were close to 90%). Taking the best models into account (LR, DNN, and SVM), two main findings are important first, all the models can be employed for distinguishing patients from HCs in a real-life environment and second that the measures we employed have discriminatory power. Although these are multi-class rather than pure HC vs. Patient, their reported AUC-ROC and AUC-PR reflect overall separability.

**Table 4.**
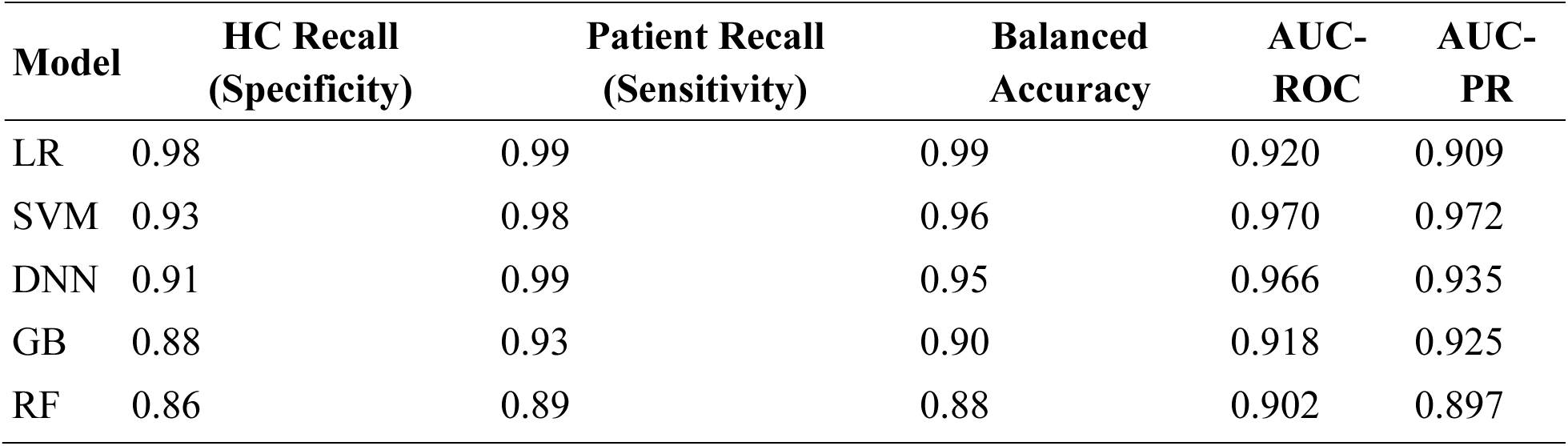
Model performance of the binary classification “Patient Group” vs. Healthy Controls.

### 3.2 How well does the ML model distinguish each sub-group in the data?

Above we collapsed all patients into one group, to determine howe well the model identifies patients from HCs. In this section, we discuss the performance of the models as multiclass classifiers, to determine how well the model distinguishes each group from each individual subgroup.

When examining the classifier’s performance on all categories, all models demonstrate robust performance with scores predominantly above 0.8 across most metrics (Figure 4 and Supplementary Data 4). For the detection of patients with LHD, all models excel here (F1 ≥ 0.92), with SVM slightly edging out the others (0.96) thanks to near-perfect precision (0.94) and recall (0.99). For patients with dementia LR is most balanced (F1 = 0.88), combining good precision (0.83) with high recall (0.94). The DNN overcalls patients (precision 0.61) despite high recall (0.98), yielding a lower F1. The detection of the HC within the LR again leads (F1 = 0.98), misclassifying only ∼2 % of controls, while the tree-based models lag (GB 0.85, RF 0.81). For the MCI, SVM outperformed the other models (F1 = 0.60) by balancing 0.63 precision with 0.56 recall. The detection of minority classes was poor, namely patients with RHD (DNN, F = 0.56 and low precision 0.17) comes at poor and TBI (SVM, F1 = 0.58, combining 0.71 precision with 0.50 recall). These suggests either both the need for more data or that language markers are overlapping so that the models are not discriminating these groups well. This will become evident from the following statistical analysis of markers associated with each condition in the following sections.

To address the problem of the minority classes, we collapsed the patient categories with MCI, RHD, and TBI into a category “Other Neurological Conditions”. In this way, the model has an exceptionally good performance, allowing the detection of patients with Dementia, LHD, and HCs and all the minority classes together. In this case, the model-specific performance across all categories. SVM demonstrates consistent performance with balanced precision and recall across LHD (precision: 0.94, recall: 0.99), Dementia (precision: 0.89, recall: 0.83), HC (precision: 0.94, recall: 0.93), and Other neurological conditions (precision: 0.94, recall: 0.72). In contrast, DNN exhibits perfect precision for LHD (1.00) but shows high recall sensitivity for Dementia (0.98) and Other Neurological conditions (0.93) at the cost of reduced precision (0.61 and 0.59, respectively). Support values indicate the sample sizes for each category: LHD (n=1173), HC (n=573), Other (n=211), and Dementia (n=47), with Dementia representing the smallest patient subgroup.

### 3.3 Which linguistic measures differ most due to diagnostic groups?

Healthy Controls (HC) served as the intercept, and the estimates for each diagnostic group (LHD, Dementia, MCI, RHD, TBI) represent the difference from this HC baseline. The analysis of various linguistic measures reveals that the *diagnosis* has a statistically significant and often substantial impact across a wide array of speech and language characteristics provides the top features with the largest explanatory power related to neurological condition. The complete results are shown in Appendix 2.

The strength of this impact, however, varies considerably among measures, as indicated by Partial Eta Squared (Partial *η*²) values for the Diagnosis and the Marginal R-squared (R² Marginal) for the overall fixed effects of the models is shown in Table 5. All p-values for the reported F-statistics are extremely small (e.g., *p <* .001), indicating high statistical significance for the effect of Diagnosis on these measures. Note that from the presentation below we have removed measures with extremely high Partial *η*² values but very low denominator degrees of freedom, suggesting their large effect sizes in this sample should be interpreted with caution due to potential model instability or low power for the inferential test despite the large point estimate of effect, also removed were measures with non-significant effects of diagnosis.

**Table 5.** Measures ranked by effect size, highlighting Large and Robust Effect Sizes (Partial *η*² > 0.15). The table presents the top 29 linguistic features ranked by partial eta-squared values, representing the proportion of variance in each measure explained by diagnostic group membership. Features are categorized into five linguistic domains: Morphology (word structure and grammatical forms), Lexicon (vocabulary and word usage), Phonology (sound patterns and syllable structure), and Syntax (grammatical relationships and phrase structure). F-statistics, degrees of freedom (Num DF = numerator, Den DF = denominator), p-values, partial *η*², and marginal R² values are reported for each measure. Morphological features dominate the top rankings, with Indefinite Count showing the largest effect size (partial *η*² = 0.29, F = 87.39, *p <* 0.001), followed by Cardinal Number Count (partial *η*² = 0.26, F = 90.88, *p <* 0.001). Lexical diversity measures (Types, Content Words Unique) and phonological complexity features (CVCC, syllable patterns) also demonstrate substantial discriminative power. All reported features achieved statistical significance (*p <* 0.001) with effect sizes meeting the threshold for practical significance in neurological assessment.

| | Measure | *Categor*<br>*y* | *F* | Num<br>DF | Den DF | *p*<br>value | Partial $\eta^2$ | $R^2$<br>Marginal |
| --- | --- | --- | --- | --- | --- | --- | --- | --- |
| 1 | Indefinite Count | Morpho<br>logy | 87.39 | 2.00 | 431.02 | 1.38E-32 | 0.29 | 0.26 |
| 2 | Cardinal<br>Number Count | Morpho<br>logy | 90.88 | 2.00 | 526.94 | 1.24E-34 | 0.26 | 0.24 |
| 3 | Types | Lexicon | 62.63 | 2.00 | 598.34 | 2.02E-25 | 0.17 | 0.19 |
| 4 | CVCC | Phonolo<br>gy | 63.29 | 2.00 | 630.04 | 8.96E-26 | 0.17 | 0.20 |
| 5 | Number Plural<br>Count | Morpho<br>logy | 56.81 | 2.00 | 568.22 | 3.22E-23 | 0.17 | 0.19 |
| 6 | Content Words<br>Unique | Lexicon | 57.43 | 2.00 | 577.53 | 1.79E-23 | 0.17 | 0.18 |
| 7 | Attribute Count | Syntax | 67.64 | 2.00 | 683.24 | 1.57E-27 | 0.17 | 0.24 |
| 8 | 2 syllables word | Phonology | 57.92 | 2.00 | 594.29 | 1.05E-23 | 0.16 | 0.19 |
| 9 | Appositional modifier Count | Syntax | 41.96 | 2.00 | 431.04 | 2.24E-17 | 0.16 | 0.21 |
| 10 | Degree Positive Count | Morphology | 48.38 | 2.00 | 499.73 | 6.19E-20 | 0.16 | 0.17 |
| 11 | Adjective Count | Morphology | 57.71 | 2.00 | 600.00 | 1.19E-23 | 0.16 | 0.19 |
| 12 | Adjective Phrases | Syntax | 57.47 | 2.00 | 597.62 | 1.48E-23 | 0.16 | 0.19 |
| 13 | Adjectival modifier Count | Syntax | 56.27 | 2.00 | 586.36 | 4.41E-23 | 0.16 | 0.19 |
| 14 | Numeral Count | Morphology | 88.21 | 2.00 | 941.26 | 7.68E-36 | 0.16 | 0.24 |
| 15 | Noun Count | Morphology | 52.31 | 2.00 | 570.44 | 1.39E-21 | 0.15 | 0.17 |
| 16 | Expletive Count | Morphology | 16.51 | 2.00 | 180.42 | 2.61E-07 | 0.15 | 0.11 |
| 17 | Syllables | Phonology | 53.61 | 2.00 | 591.15 | 4.03E-22 | 0.15 | 0.18 |
| 18 | CVC | Phonology | 53.58 | 2.00 | 593.36 | 4.07E-22 | 0.15 | 0.17 |
| 19 | Content Words Total | Lexicon | 51.06 | 2.00 | 576.36 | 3.82E-21 | 0.15 | 0.18 |
| 20 | Total Characters in Text Letters Only | Lexicon | 51.64 | 2.00 | 585.38 | 2.22E-21 | 0.15 | 0.17 |
| 21 | Corrected TTR CTTR | Lexicon | 82.39 | 2.00 | 937.05 | 1.09E-33 | 0.15 | 0.20 |
| 22 | Prepositional modifier Count | Syntax | 51.04 | 2.00 | 581.51 | 3.78E-21 | 0.15 | 0.18 |
| 23 | Prepositional Phrases | Syntax | 51.79 | 2.00 | 590.31 | 1.88E-21 | 0.15 | 0.18 |
| 24 | Unclassified dependent Count | Syntax | 10.19 | 2.00 | 117.38 | 8.29E-05 | 0.15 | 0.05 |
| 25 | Adposition Count | Morphology | 51.23 | 2.00 | 593.33 | 3.00E-21 | 0.15 | 0.18 |
| 26 | CV | Phonology | 59.48 | 2.00 | 693.62 | 1.44E-24 | 0.15 | 0.18 |
| 27 | Verb Phrases | Syntax | 50.38 | 2.00 | 590.91 | 6.28E-21 | 0.15 | 0.17 |
| 28 | Direct object<br>Count | Syntax | 46.34 | 2.00 | 543.58 | 2.62E-19 | 0.15 | 0.17 |
| 29 | Words Tokens | Lexicon | 49.53 | 2.00 | 582.53 | 1.36E-20 | 0.15 | 0.17 |

D*iagnosis* demonstrates a widespread influence on a multitude of linguistic measures. The strongest differentiating features (those with large Partial *η*² values and robust model fits) are concentrated in areas of semantic content (especially numerical and definiteness marking), overall lexical production and diversity, counts of various morphological categories (nouns, adjectives, plurals), and basic phonological/syllable structure counts. Additionally, measures of syntactic complexity and certain readability characteristics also show substantial impact.

These findings highlight that the neurological conditions under study manifest with distinct and quantifiable linguistic profiles. The identified measures with the largest effect sizes are prime candidates for inclusion in diagnostic models or for tracking linguistic changes associated with these conditions. The high *R*² Marginal values for many of these top-ranking measures further underscore the explanatory power of Diagnosis in accounting for the observed linguistic variations. A substantial number of linguistic measures demonstrated large and robust effects of Diagnosis, indicating these are strong candidates for differentiating between the groups. These involve all the aspects of grammar like phonology, morphology, syntax and semantics, lexical usage, and readability that is text difficulty.

#### Measures with Medium Effects (Partial *η*² ∼0.06 – 0.13)

Beyond the large effects, a broad range of other measures showed medium-sized effects of Diagnosis. These span across all linguistic domains, which we included like the total Number of Function Words (Partial *η*² = 0.14), phonology, such as the different syllable types, like VC and CCVCC (Partial *η*² = 0.14), morphology including the Number of Verbs (Partial *η*² = 0.14), syntax like the number of Complex thematic units (*T* units), the number of matrix sentences (Root), dependent clauses, and the object of preposition. As discussed below although the readability measures did not make it to the list shown in Table 5, several readability measures remain important as they achieve a Partial *η*² between 0.14 and 0.13; these include the Estimated Reading Time (sec), Smog Index, Total Classical Yngve Load, Difficult Words; the latter is a measure based on a standardized dictionary (Themistocleous, 2024).

### 3.4 Which are the distinctive features for each neurological condition compared to HC?

In this section, we summarize the high-level “linguistic signatures” that distinguish each group. Table 6 below synthesizes the results for each neurological condition, by highlighting the top ten (10) linguistic features that most strongly distinguish it from Healthy Controls by using the magnitude of the z scores from the post-hoc analysis (emmeans). The complete list of distinctive linguistic features is provided in the Supplementary Table 5.

**Table 6.**
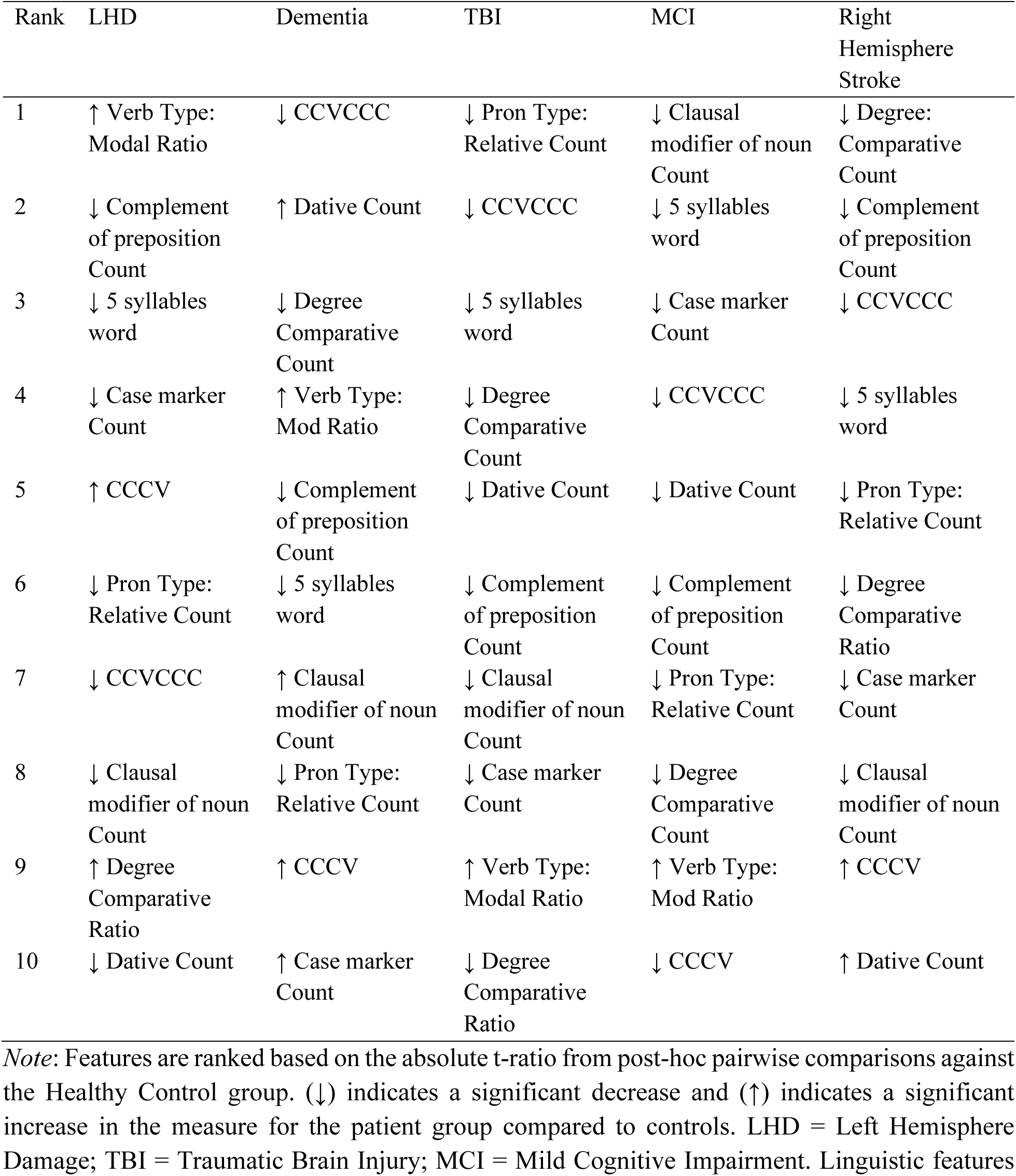
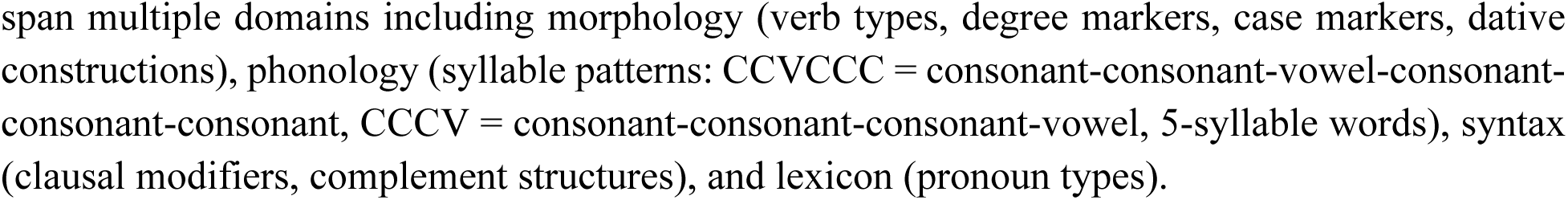
Top 10 distinctive linguistic features for each neurological condition compared to healthy controls. Features are ranked by absolute t-ratio values from post-hoc pairwise comparisons, identifying the most diagnostically discriminative linguistic markers for each condition. Upward arrows (↑) indicate significantly increased measures in patient groups relative to healthy controls; downward arrows (↓) indicate significantly decreased measures.

| Rank | LHD | Dementia | TBI | MCI | Right Hemisphere Stroke |
| --- | --- | --- | --- | --- | --- |
| 1 | ↑ Verb Type: Modal Ratio | ↓ CCVCCC | ↓ Pron Type: Relative Count | ↓ Clausal modifier of noun Count | ↓ Degree: Comparative Count |
| 2 | ↓ Complement of preposition Count | ↑ Dative Count | ↓ CCVCCC | ↓ 5 syllables word | ↓ Complement of preposition Count |
| 3 | ↓ 5 syllables word | ↓ Degree Comparative Count | ↓ 5 syllables word | ↓ Case marker Count | ↓ CCVCCC |
| 4 | ↓ Case marker Count | ↑ Verb Type: Mod Ratio | ↓ Degree Comparative Count | ↓ CCVCCC | ↓ 5 syllables word |
| 5 | ↑ CCCV | ↓ Complement of preposition Count | ↓ Dative Count | ↓ Dative Count | ↓ Pron Type: Relative Count |
| 6 | ↓ Pron Type: Relative Count | ↓ 5 syllables word | ↓ Complement of preposition Count | ↓ Complement of preposition Count | ↓ Degree Comparative Ratio |
| 7 | ↓ CCVCCC | ↑ Clausal modifier of noun Count | ↓ Clausal modifier of noun Count | ↓ Pron Type: Relative Count | ↓ Case marker Count |
| 8 | ↓ Clausal modifier of noun Count | ↓ Pron Type: Relative Count | ↓ Case marker Count | ↓ Degree Comparative Count | ↓ Clausal modifier of noun Count |
| 9 | ↑ Degree Comparative Ratio | ↑ CCCV | ↑ Verb Type: Modal Ratio | ↑ Verb Type: Mod Ratio | ↑ CCCV |
| 10 | ↓ Dative Count | ↑ Case marker Count | ↓ Degree Comparative Ratio | ↓ CCCV | ↑ Dative Count |

*Note:* Features are ranked based on the absolute t-ratio from post-hoc pairwise comparisons against the Healthy Control group. (↓) indicates a significant decrease and (↑) indicates a significant increase in the measure for the patient group compared to controls. LHD = Left Hemisphere Damage; TBI = Traumatic Brain Injury; MCI = Mild Cognitive Impairment. Linguistic features
span multiple domains including morphology (verb types, degree markers, case markers, dative constructions), phonology (syllable patterns: CCVCCC = consonant-consonant-vowel-consonant-consonant-consonant, CCCV = consonant-consonant-consonant-vowel, 5-syllable words), syntax (clausal modifiers, complement structures), and lexicon (pronoun types).

Several key patterns emerge from the statistical analysis. Individuals with LHD are characterized by a widespread disruption across multiple linguistic domains. While the most discriminating feature is an increased ratio of verb-modifying word types, the majority of the top-10 features are decreases (↓), reflecting a reduction in phonological complexity (e.g., 5 syllables word, CCVCCC), syntactic structures (Complement of preposition), and the use of specific word types (Pronoun Type: Relative Pronouns). Individuals with Dementia show a pattern of impairment that is also broad but appears centered on the use of specific content and function words (Verb Type: Modal) and a decrease relative to HCs on measures of phonological complexity (5 syllables word, CCVCCC). Individuals with TBI present a mixed profile of mostly decreases in its top features, suggesting a unique pattern of linguistic disruption with a notable increases in of ratio of Modal Verbs. Individuals with MCI is uniquely distinguished by a strong decrease in measures that associated with increased production complexity like five (5) syllable-words, syllables with complex articulatory patterns (CCVCCC, CCCV) and complex syntactic patterns such as the number of Clausal Modifier of Nouns and Complement of Prepositions. This pattern of decreased production in several of the top-ranking features supports the hypothesis that individuals with MCI more general disruptions in language and domains like memory that can explain their use of simpler patterns. Individuals with RHD shows the most subtle linguistic profile. Its top discriminators are related to the diminished production of the number Comparative Adjectives, Complements of Prepositions and complex syllable patterns (CCVCCC) and phonological structures (5 syllable-words).

The primary linguistic signatures based on measures that resulted in statistical significance are described in the next section.

### 3.5 What do language measures reveal about each patient group?

The detailed statistical analyses reveal distinct linguistic profiles for each diagnostic group when compared to healthy controls. The nature of these differences—whether reductions in production, complexity, or alterations in proportional usage of linguistic features—varies by measure and group, underscoring the unique impact of each neurological condition on language processing. We present the results only for the measures that had an overall significant effect with large and medium effect sizes. For a comprehensive list of all statistical comparisons for every measure, please refer to the Supplementary Data 6.

#### 3.5.1 Lexical Markers

Analysis of lexical features revealed distinct profiles across the diagnostic groups when compared to Healthy Controls (HC). The findings indicate that phonological impairments follow a severity gradient, with LHD and individuals with TBI showing the most pronounced deficits, individuals with MCI displaying moderate impairments, and dementia and individuals with RHD maintaining relatively preserved abilities compared to healthy controls.

**Figure 3.**
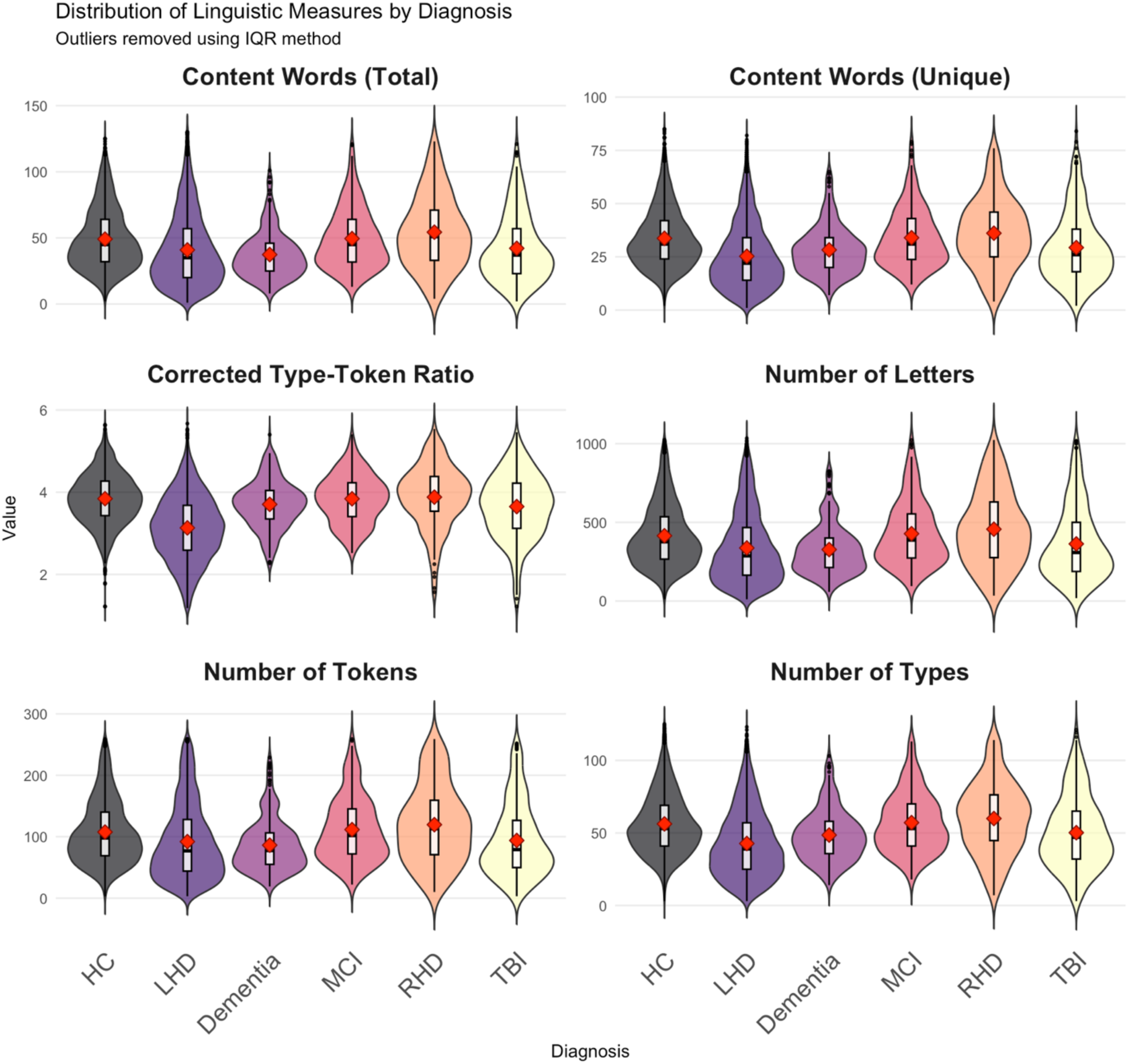
shows key lexical measures selected based on the effect size (*η*^2^).

**Figure 3.**
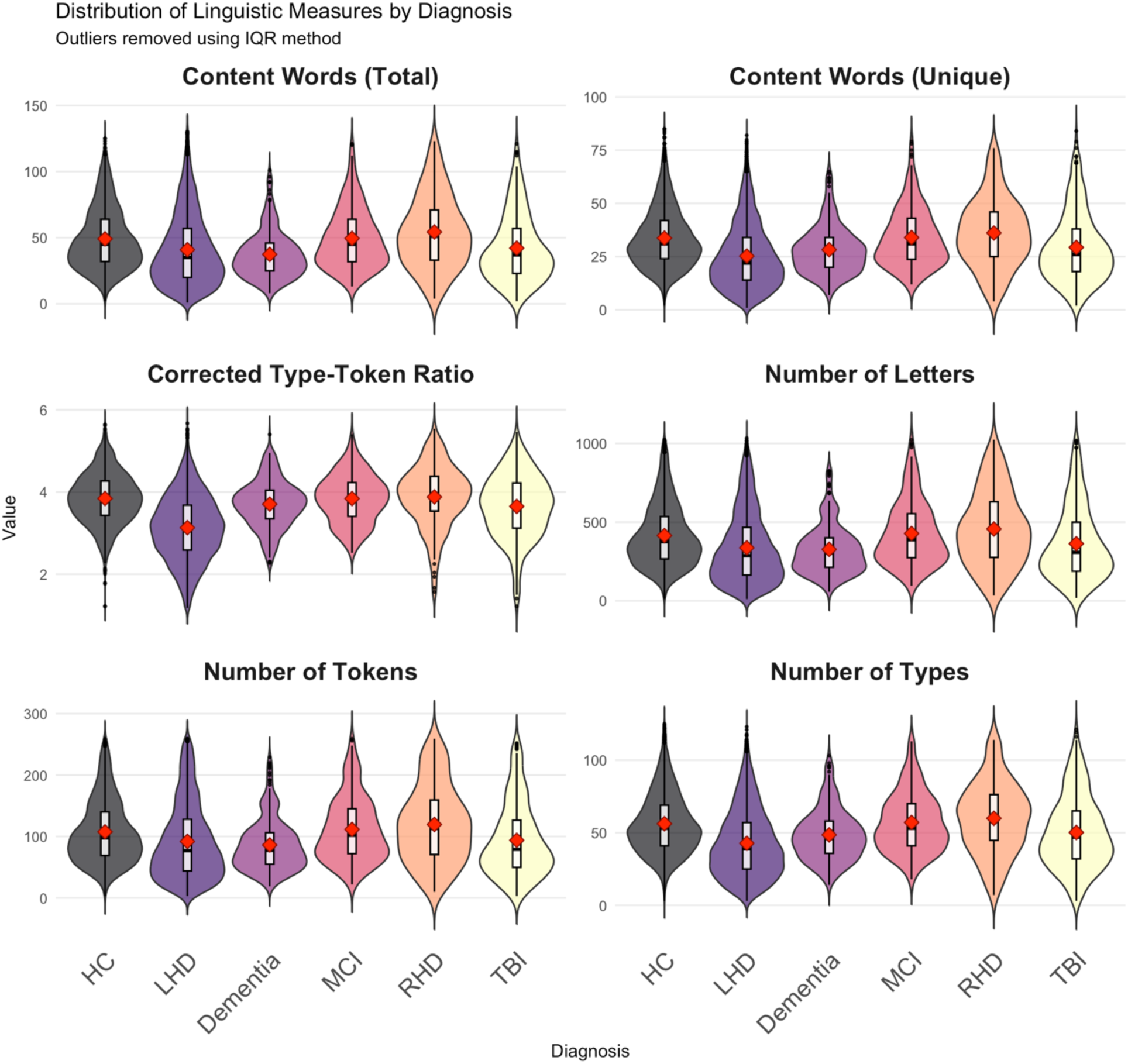
Distribution of key linguistic measures across neurological diagnostic groups. Violin plots display the probability density distribution of six fundamental linguistic features, with outliers removed using the interquartile range (IQR) method. Each violin shows the full distribution shape, with embedded box plots indicating median (white line), first and third quartiles (box boundaries), and range (whiskers). Red dots represent mean values for each group. Diagnostic groups include HC = Healthy Controls, LHD = Left Hemisphere Damage, Dementia, MCI = Mild Cognitive Impairment, RHD = Right Hemisphere Damage, TBI = Traumatic Brain Injury. The measures represent core aspects of language production: Content Words (Total) measures semantic word usage; Content Words (Unique) reflects lexical diversity; Corrected Type-Token Ratio (CTTR) quantifies vocabulary richness adjusted for text length; Number of Letters indicates overall text production; Number of Tokens represents total word output; Number of Types measures vocabulary breadth. Notable patterns include reduced lexical diversity in dementia (lower CTTR and unique content words), variable text production across conditions (Number of Letters and Tokens), and preserved vocabulary breadth in some conditions despite reduced overall output. Healthy controls generally show the most consistent distributions, while neurological conditions exhibit increased variability and condition-specific patterns. These distributions demonstrate the diagnostic potential of quantitative linguistic analysis for differentiating neurological conditions based on language production characteristics.

##### 3.5.1.1 Individuals with LHD

The LHD group demonstrated a consistent and statistically significant reduction in performance across nearly all measures of lexical productivity and diversity compared to healthy controls. This group produced significantly fewer total words (*β* = –69.67, *p <* .001), content words (*β* = –33.49, *p <* .001), and unique word types (*β* = –28.47, *p <* .001). Furthermore, their lexical diversity was significantly lower, as measured by the Corrected Type-Token Ratio (CTTR; *β* = –0.81, *p <* .001). An interesting exception was observed in Maas’s TTR, where the LHD group scored significantly *higher* than controls (*β* = 0.007, *p <* .001), suggesting a more complex pattern of lexical usage.

##### 3.5.1.2 Individuals with Dementia

The Dementia group exhibited a pattern of lower, albeit less pronounced, lexical performance. They showed a statistically significant reduction in average word length (*β* = –0.06, *p =* .011) and Corrected TTR (*β* = –0.13, *p =* .003) compared to the HC group. On measures of word productivity, such as the total number of content words or unique types, the Dementia group scored lower than controls, but these differences did not reach statistical significance (e.g., for unique content words, *p =* .126).

##### 3.5.1.3 Individuals with MCI

The MCI group displayed a consistent and significant reduction in lexical productivity. This group produced significantly fewer total words (*β* = –58.59, *p <* .001), content words (*β* = –26.46, *p <* .001), function words (*β* = –30.12, *p <* .001), and unique word types (*β* = –14.36, *p <* .001) than their healthy peers. In contrast, their lexical diversity, as measured by the standard Type-Token Ratio (TTR), was significantly *higher* than the HC group (*β* = 0.02, *p =* .008).

##### 3.5.1.4 Individuals with TBI

The TBI group showed a performance profile marked by significantly reduced lexical productivity but increased lexical diversity. They produced significantly fewer total words (*β* = –65.50, *p <* .001), unique content words (*β* = –15.10, *p <* .001), and unique function words (*β* = –4.30, *p <* .001). Concurrently, their TTR was significantly *higher* than that of the HC group (*β* = 0.04, *p <* .001), suggesting that while they produced fewer words overall, the vocabulary they used was more varied.

##### 3.5.1.5 Individuals with RHD

Across all lexical measures analyzed, the RHD group’s performance was not statistically different from that of the healthy control group. The estimates for this group were consistently small and associated with non-significant *p*-values (all *p*s > .14), indicating a comparable lexical profile to the healthy controls on these specific tasks.

#### 3.5.2 Phonological Markers

Phonological measures relate to sound structures, syllable complexity, and word length in terms of characters and syllables. Individuals with different diagnosis differed in the number of distinctively characteristics measures.

**Figure 4.**
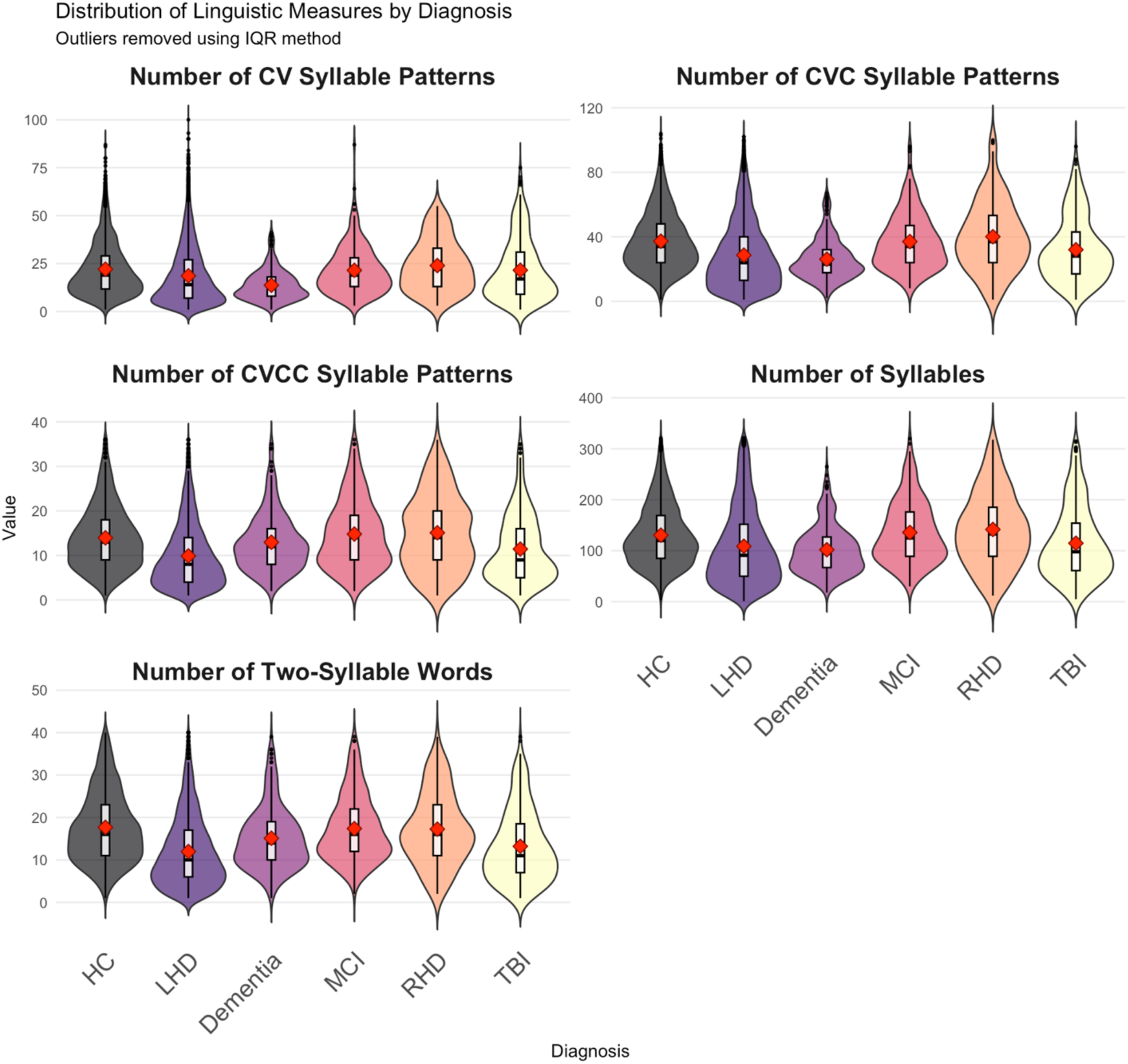
shows key lexical measures selected based on the effect size (*η*^2^).

**Figure 4.**
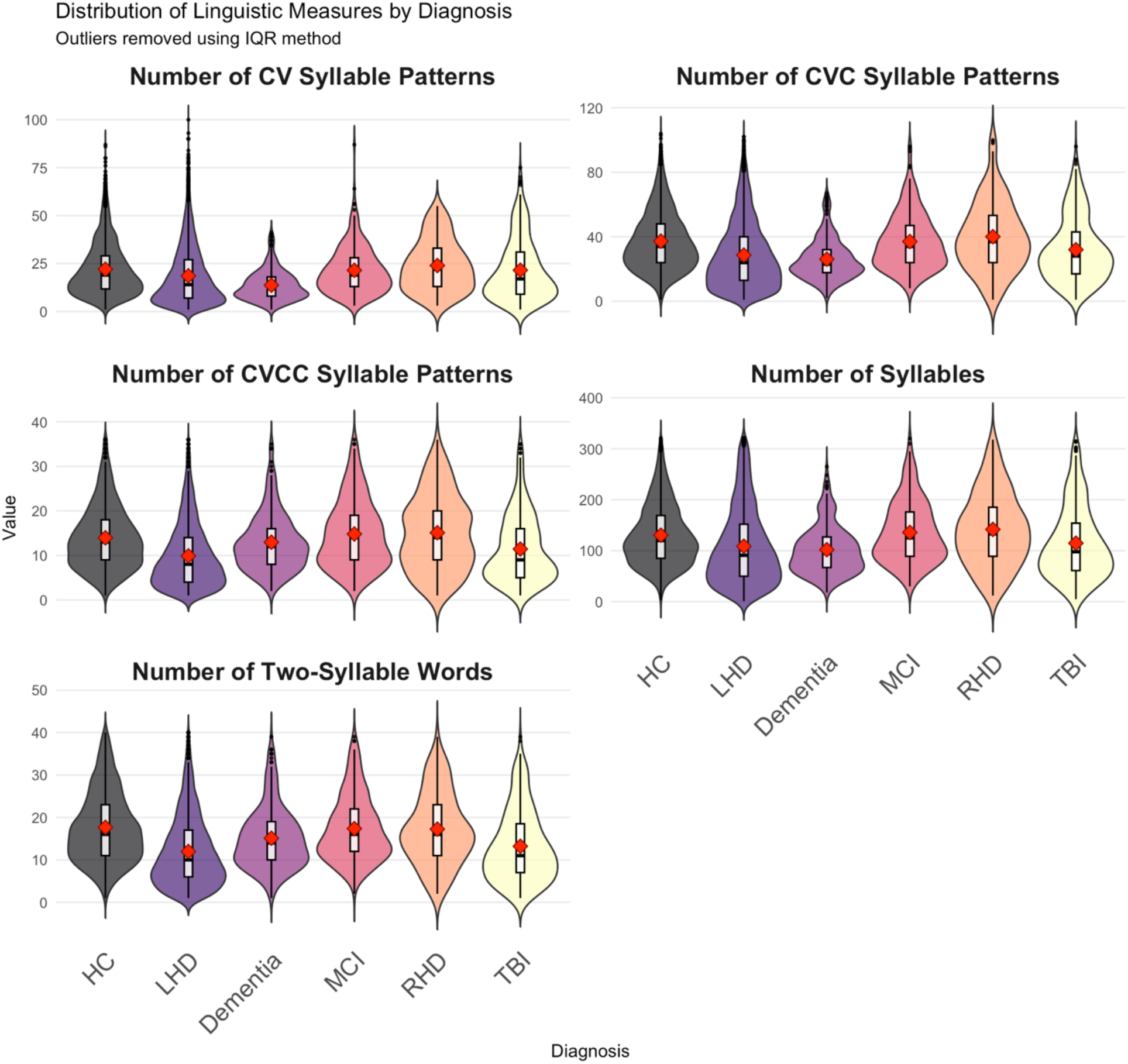
Violin plots showing distribution density with boxplots and mean values (red diamonds) of phonology measures by diagnosis. Distribution of phonological complexity measures across neurological diagnostic groups. Violin plots display the probability density distribution of five phonological features that capture syllable structure complexity and phonological processing abilities, with outliers removed using the interquartile range (IQR) method. Each violin shows the full distribution shape, with embedded box plots indicating median (white line), first and third quartiles (box boundaries), and range (whiskers). Red dots represent mean values for each group. Diagnostic groups include HC = Healthy Controls, LHD = Left Hemisphere Damage, Dementia, MCI = Mild Cognitive Impairment, RHD = Right Hemisphere Damage, TBI = Traumatic Brain Injury. Phonological measures include CV (Consonant-Vowel) patterns representing the simplest syllable structure; CVC (Consonant-Vowel-Consonant) patterns indicating moderate complexity; CVCC (Consonant-Vowel-Consonant-Consonant) patterns reflecting higher phonological complexity; Number of Syllables measuring overall phonological output; Number of Two-Syllable Words indicating specific syllable length preferences. Notable patterns include preserved simple syllable structures (CV, CVC) across most conditions, while complex syllable patterns (CVCC) show greater variability and potential reduction in certain neurological conditions. RHD and TBI groups demonstrate elevated usage of complex syllable structures compared to other patient groups, possibly reflecting compensatory strategies or preserved phonological complexity. These phonological measures provide insights into the articulatory and phonological planning abilities affected differentially across neurological conditions, complementing lexical and syntactic analyses for comprehensive linguistic assessment.

##### 3.5.2.1 Individuals with LHD

Individuals with LHD had the most severe phonological impairments across virtually all measures. Significant reductions were observed in word production across syllable lengths, with 1-syllable words showing a 48.27-point decrease (*p <* 0.001), 2-syllable words decreasing by 15.47 points (*p <* 0.001), and 4-syllable words declining by 3.49 points (*p <* 0.001). Total syllable production was reduced by 107.13 (*p <* 0.001), and overall character output decreased by 391.83 characters (*p <* 0.001). Most syllable structures showed significant impairments, including CV patterns (*β* = –23.79, *p <* 0.001) and CVC patterns (*β* = –36.56, *p <* 0.001). Notably, individuals with LHD showed a rare positive effect in CCCV patterns (+0.92, *p <* 0.001).

##### 3.5.2.2 Individuals with Dementia

Individuals with dementia showed relatively preserved phonological abilities compared to other groups. Most measures showed no significant differences from healthy controls, with p-values consistently exceeding 0.05. While numerical trends suggested slight reductions in some areas (e.g., *β* = –7.30 for 1-syllable words, *β* = –15.88 for total syllables), these differences were not statistically significant.

##### 3.5.2.3 Individuals with MCI

Individuals with MCI showed moderate but consistent phonological difficulties across most measures. Significant reductions were found in 1-syllable words (*β* = –43.47, *p <* 0.001), 2-syllable words (*β* = –9.78, *p <* 0.001), and 4-syllable words (*β* = –1.58, *p <* 0.001). Total syllable production was reduced by 74.87 (*p <* 0.001), with character output decreasing by 302.66 characters (*p <* 0.001). Several syllable structures showed significant deficits, including CV patterns (*β* = –15.64, *p <* 0.001) and CVC patterns (*β* = –22.47, *p <* 0.001).

##### 3.5.2.4 Individuals with TBI

Individuals with TBI exhibited substantial phonological deficits similar in magnitude to LHD patients. Significant reductions included 1-syllable words (*β* = –48.52, *p <* 0.001), 2-syllable words (*β* = –12.38, *p <* 0.001), and 4-syllable words (*β* = –2.55, *p <* 0.001). Total syllable production decreased by 90.62 (*p <* 0.001), with corresponding reductions in character output (*β* = –342.62, *p <* 0.001). Multiple syllable structures showed significant impairments, including CV (*β* = –19.18, *p <* 0.001) and CVC patterns (*β* = –28.61, *p <* 0.001).

##### 3.5.2.5 Individuals with RHD

Individuals with RHD showed largely intact phonological functioning with minimal significant effects. Most measures revealed no significant differences from controls, suggesting that phonological processing remains relatively preserved following right hemisphere injury. Only 4-syllable word production showed a small but significant reduction (*β* = –1.35, *p =* 0.020).

#### 3.5.3 Morphological Markers

Morphology provides insights into how different patient populations structure their utterances at the word level.

**Figure 5.**
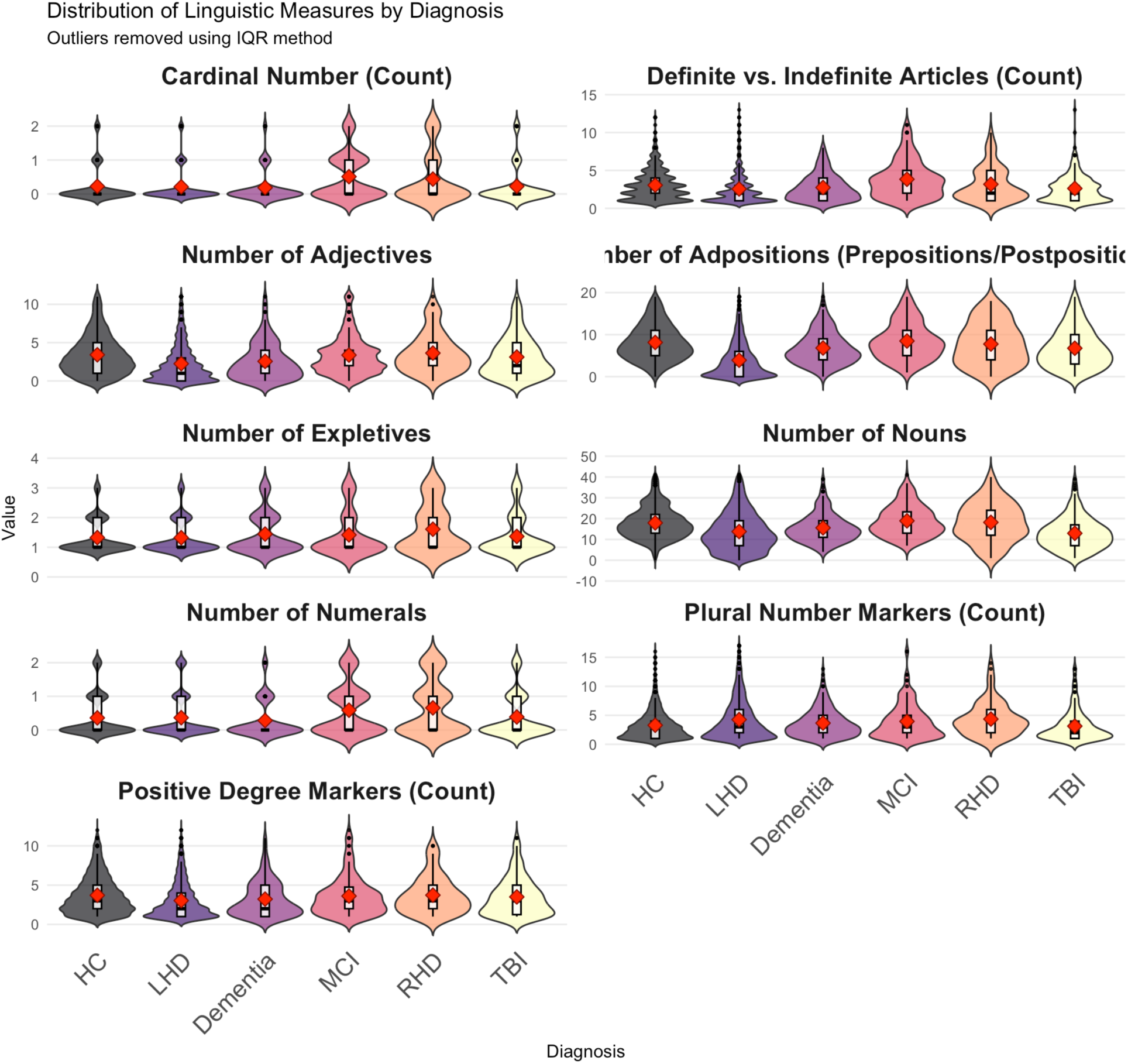
shows the distribution of the morphological measures selected based on the effect size (*η*^2^). In the following, we discuss the part of speech measures first, and as we see the individual linguistic profile for each diagnostic group had significantly varied with high effect size for adjectives, expletives, nouns, and numerals.

**Figure 5.**
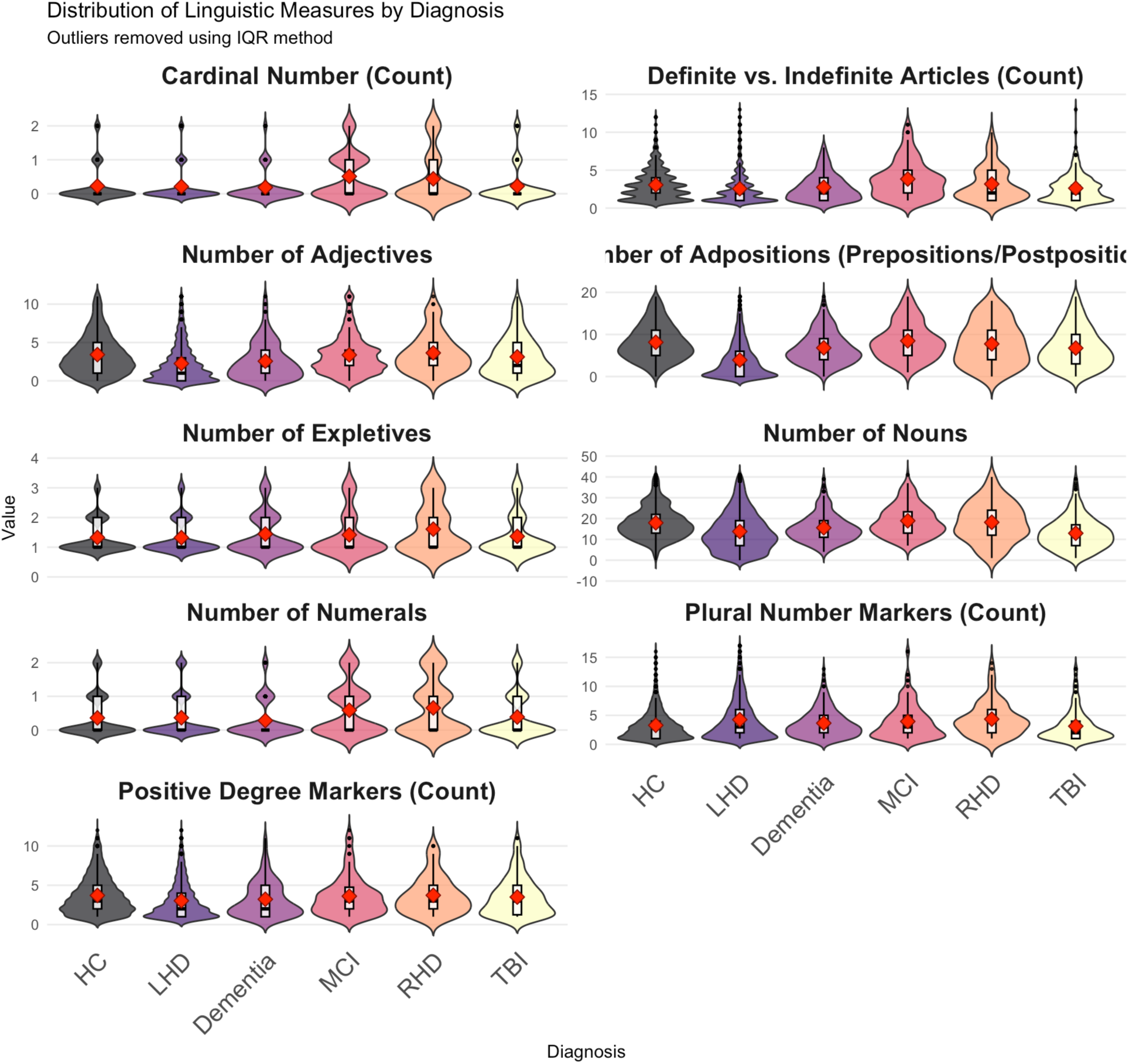
Distribution of morphological complexity measures across neurological diagnostic groups. Violin plots display the probability density distribution of nine phonological features that capture syllable structure complexity and phonological processing abilities, with outliers removed using the interquartile range (IQR) method: Cardinal Number (Count), ranging from 0-2; Definite vs. Indefinite Articles (Count), ranging from 0-15; Number of Adjectives, ranging from 0-10; Number of Adpositions (Prepositions/Postpositions), ranging from 0-20; Number of Expletives, ranging from 0-4; Number of Nouns, ranging from –10 to 50; Number of Numerals, ranging from 0-2; Plural Number Markers (Count), ranging from 0-15; and Positive Degree Markers (Count), ranging from 0-10. Each violin shows the full distribution shape, with embedded box plots indicating median (white line), first and third quartiles (box boundaries), and range (whiskers). Red dots represent mean values for each group. Diagnostic groups include HC = Healthy Controls, LHD = Left Hemisphere Damage, Dementia, MCI = Mild Cognitive Impairment, RHD = Right Hemisphere Damage, TBI = Traumatic Brain Injury.

##### 3.5.3.1 Individuals with LHD

Individuals with LHD displayed a mixed profile. They showed a significantly lower ratio of adjectives, adverbs, conjunctions, and determiners. For example, the Adjective Ratio was significantly reduced (*β* = –0.008, *p <* .001). Conversely, they demonstrated a significantly higher ratio of nouns, proper nouns, and interjections. The proportional use of proper nouns, for instance, was markedly increased (*β* = 0.03, *p <* .001).

##### 3.5.3.2 Individuals with Dementia

Individuals with dementia had largely comparable proportional use of morphology to HCs, with only a few exceptions, such as a higher ratio of nouns in the nominative case (*β* = 0.003, *p =* .017).

##### 3.5.3.3 Individuals with MCI

Individuals with MCI showed significant differences on several ratios. They used a lower proportion of adverbs (*β* = –0.008, *p <* .001) and coordinating conjunctions (*β* = –0.005, *p =* .023) but a higher proportion of plural nouns (*β* = 0.004, *p =* .025) and nouns in the accusative case (*β* = 0.002, *p =* .007).

##### 3.5.3.4 Individuals with TBI

Individuals with TBI also exhibited a mixed pattern. They showed a significantly lower ratio of coordinating conjunctions (*β* = –0.009, *p <* .001) but a higher ratio of nouns in the accusative case (*β* = 0.003, *p <* .001) and words with perfective aspect (*β* = 0.004, *p <* .001) and passive voice (*β* = 0.002, *p <* .001).

##### 3.5.3.5 Individuals with RHD

Individuals with RHD was again broadly like HCs, though they showed a significantly lower use of second-person pronouns (*β* = –0.006, *p =* .001) and a higher use of first-person pronouns (*β* = 0.007, *p <* .001).

#### 3.5.4 Syntactic Characteristics

Syntactic measures evaluate sentence structure, complexity, and the use of different phrase and clause types.

**Figure 6.**
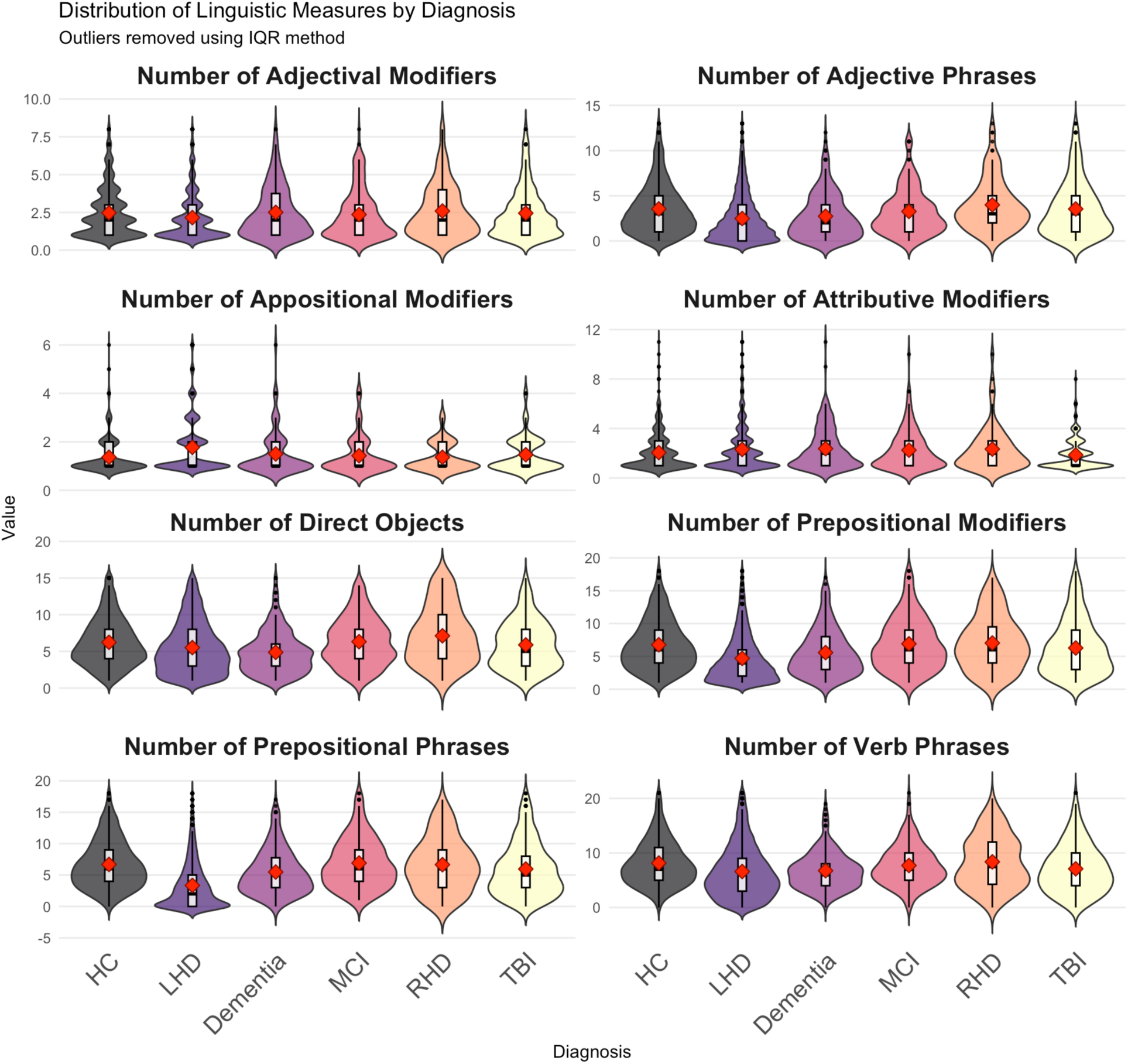
shows the distribution of the key syntactic measures by diagnosis selected based on the model’s effect size (*η*^2^).

**Figure 6.**
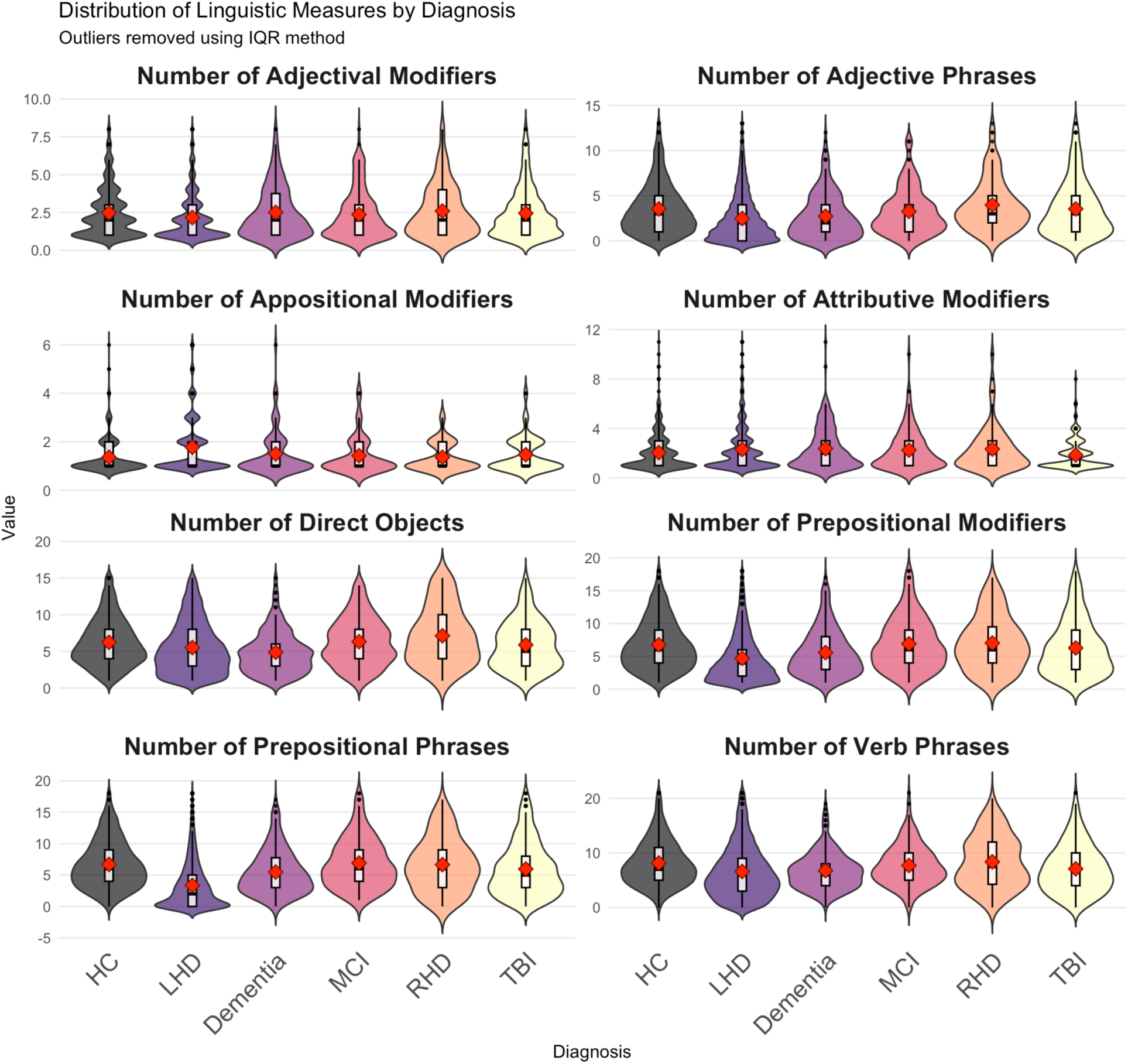
Violin plots showing distribution density with boxplots and mean values (red diamonds) of syntactic measures by diagnosis. Each violin shows the full distribution shape, with embedded box plots indicating median (white line), first and third quartiles (box boundaries), and range (whiskers). Red dots represent mean values for each group. Diagnostic groups include HC = Healthy Controls, LHD = Left Hemisphere Damage, Dementia, MCI = Mild Cognitive Impairment, RHD = Right Hemisphere Damage, TBI = Traumatic Brain Injury.

These findings reveal a clear hierarchy of syntactic impairment severity, with LHD and individuals with TBI showing the most pronounced deficits, individuals with MCI displaying moderate impairments, and dementia and individuals with RHD maintaining relatively preserved syntactic abilities compared to healthy controls.

##### 3.5.4.1 Individuals with LHD

Individuals with LHD demonstrated the most severe and comprehensive syntactic deficits across virtually all measures. Significant reductions were observed in phrase-level structures, including adjectival complements (*β* = –1.12, *p <* 0.001), adjectival modifiers (*β* = –2.63, *p <* 0.001), adjective phrases (*β* = –3.93, *p <* 0.001), and adverbial phrases (*β* = –6.52, *p <* 0.001). Sentence complexity was markedly reduced, with average sentence length decreasing by 2.45 words (*p <* 0.001) and average tree height declining by 0.75 units (*p <* 0.001). Complex syntactic structures showed substantial impairments, including total complex nominals (*β* = –11.21, *p <* 0.001), prepositional phrases (*β* = –8.46, *p <* 0.001), and total dependent clauses (*β* = –2.06, *p <* 0.001). Interestingly, individuals with LHD showed compensatory increases in certain simpler structures, including compound modifiers (*β* = 1.91, *p <* 0.001) and appositional modifiers (*β* = 0.94, *p <* 0.001), suggesting possible strategic shifts toward less complex syntactic patterns.

##### 3.5.4.2 Individuals with Dementia

Individuals with dementia demonstrated relatively preserved syntactic abilities compared to other groups, with most measures showing no significant differences from healthy controls. While some isolated significant effects emerged, such as reduced adverbial clause modifier ratios (*β* = – 0.00216, *p =* 0.007) and prepositional modifier ratios (*β* = –0.00389, *p =* 0.022), the overall pattern suggested substantially better preservation of syntactic structure compared to LHD, TBI, and MCI groups.

##### 3.5.4.3 Individuals with MCI

Individuals with MCI showed moderate but consistent syntactic difficulties across many domains. Significant reductions were found in adjectival complements (*β* = –1.16, *p <* 0.001), adjectival modifiers (*β* = –1.65, p< 0.001), and adjective phrases (*β* = –2.60, *p <* 0.001). Sentence length was modestly reduced (*β* = –0.42 words, *p =* 0.020), while complex structures showed notable impairments, including total complex nominals (*β* = –6.67, *p <* 0.001), prepositional phrases (*β* = –4.01, *p <* 0.001), and coordinate phrases (*β* = –4.68, *p <* 0.001). Clause-level complexity was affected, with reductions in total clauses (*β* = –3.84, *p <* 0.001) and dependent clauses (*β* = –1.51, *p <* 0.001).

##### 3.5.4.4 Individuals with TBI

Individuals with TBI exhibited substantial syntactic impairments comparable in severity to LHD patients. Significant deficits included reduced adjectival complements (*β* = –1.05, *p <* 0.001), adjectival modifiers (*β* = –1.99, p< 0.001), and adjective phrases (*β* = –3.11, *p <* 0.001). Sentence structure was compromised, with average sentence length decreasing by 1.19 words (*p <* 0.001) and average tree height declining by 0.32 units (*p <* 0.001). Complex syntactic constructions showed marked reductions, including total complex nominals (*β* = –9.32, *p <* 0.001), prepositional phrases (*β* = –4.86, *p <* 0.001), and coordinate phrases (*β* = –6.16, *p <* 0.001). Overall syntactic complexity was reduced across multiple measures, including total dependent clauses (*β* = –1.95, *p <* 0.001) and complex T-units (−1.86, *p <* 0.001).

##### 3.5.4.5 Individuals with RHD

Individuals with RHD showed largely intact syntactic functioning with minimal significant impairments. Most syntactic measures remained within normal ranges, with only occasional significant effects such as reduced complement of preposition counts (−0.41, *p =* 0.023) and number modifier counts (*β* = –1.03, *p =* 0.011). This pattern indicates that syntactic processing remains relatively preserved following right hemisphere injury.

#### 3.5.5 Readability Metrics

Readability formulas estimate the level of education (e.g., first year student, second year student) needed to understand a text. Although the readability measures had a lower effect size, their effect was statistically significant. The results are presented in Table 7. The Smog Index had the highest effect size (Partial *η*² > 0.15) among the readability measures. The findings suggest that clinical populations differ not only in speech and cognitive abilities but also in the structural and lexical complexity of their language production, as quantified by standardized readability metrics.

**Figure 7.**
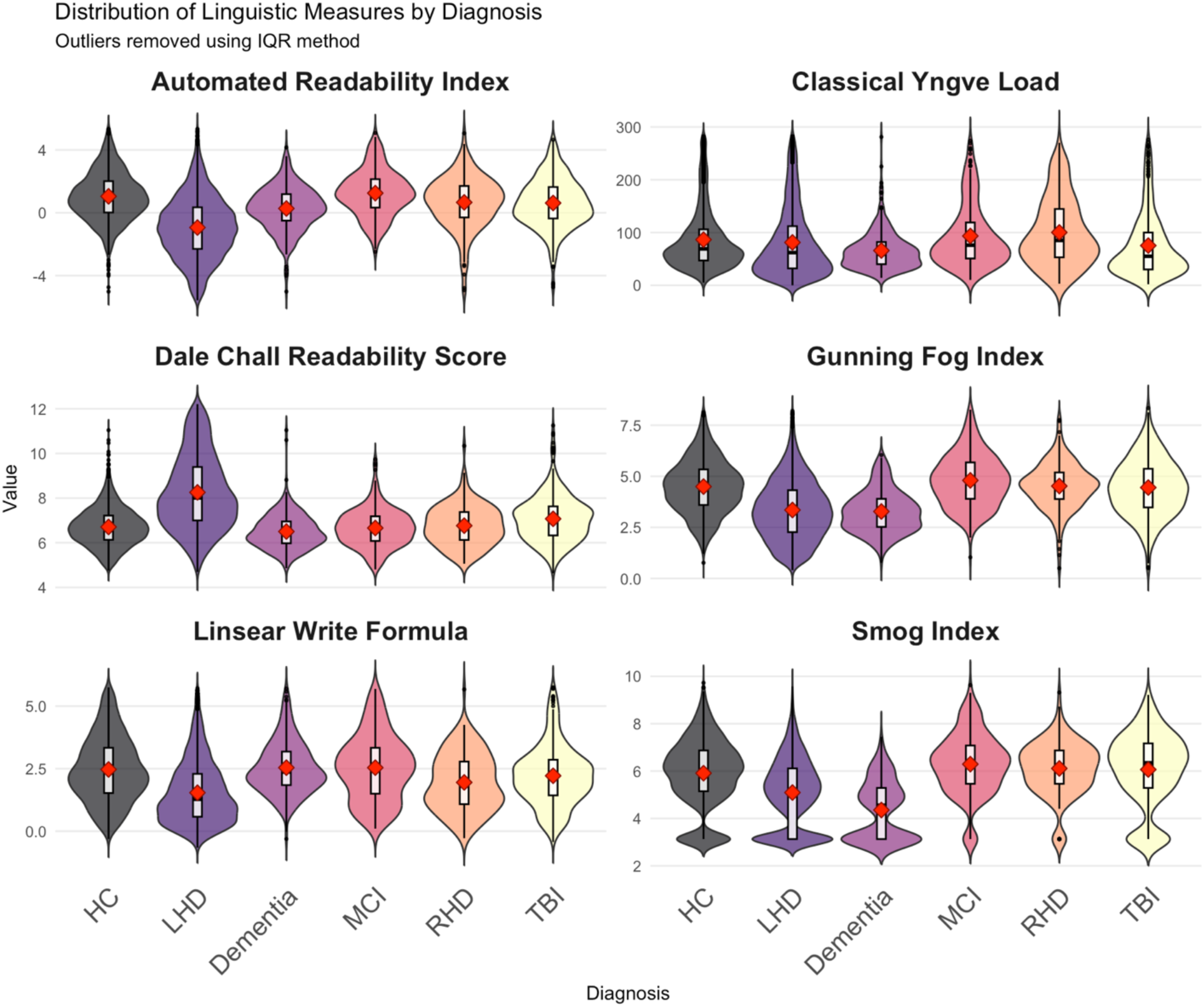
shows the violin plots displaying the distribution density with boxplots and mean values (red diamonds) of readability measures by diagnosis.

**Figure 7.**
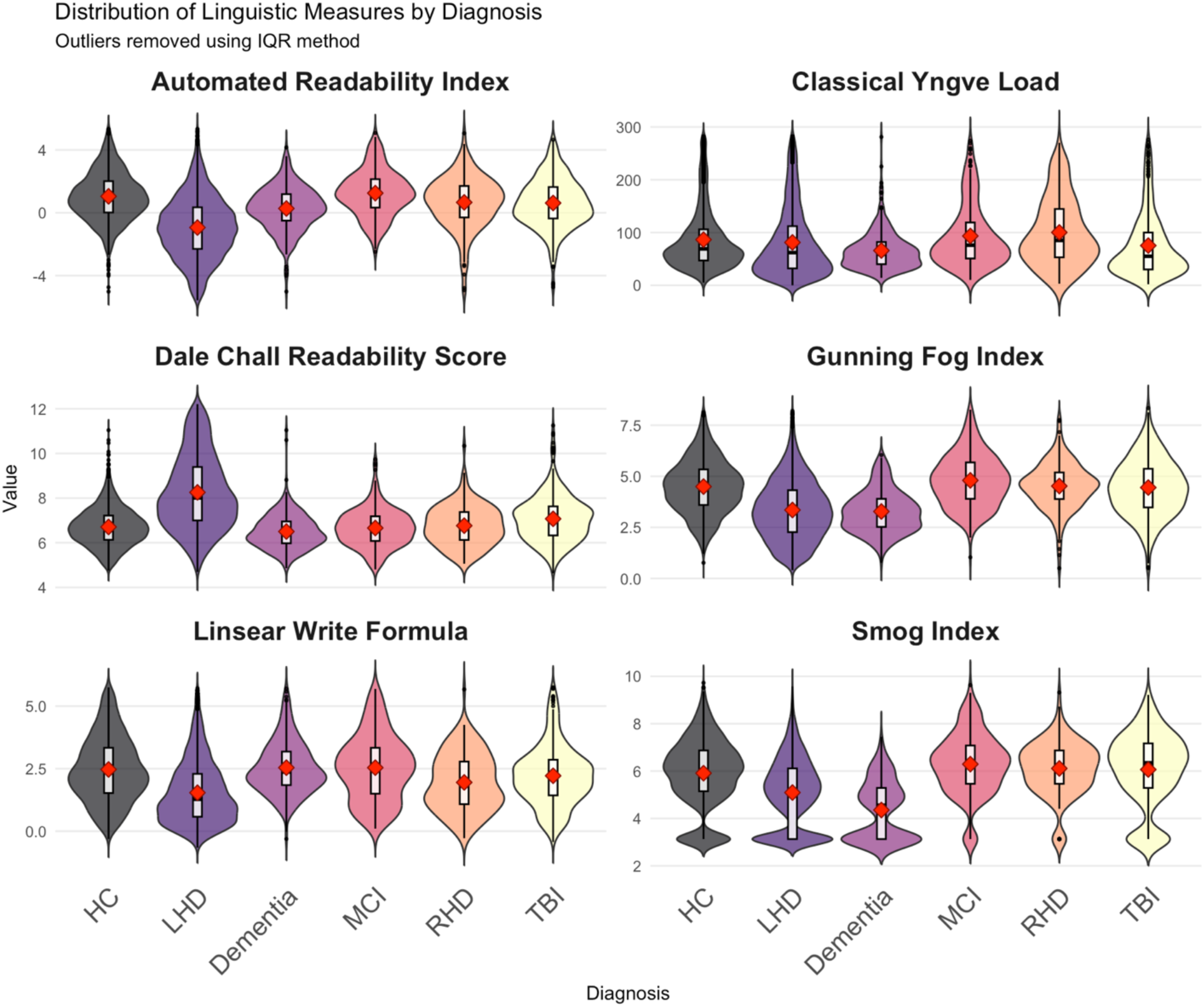
Violin plots showing distribution density with boxplots and mean values (red diamonds) of readability measures by diagnosis (HC: Healthy Controls, LHD: Left Hemisphere Damage, MCI: Mild Cognitive Impairment, RHD: Right Hemisphere Damage, TBI: Traumatic Brain Impairment).

**Figure 10.**
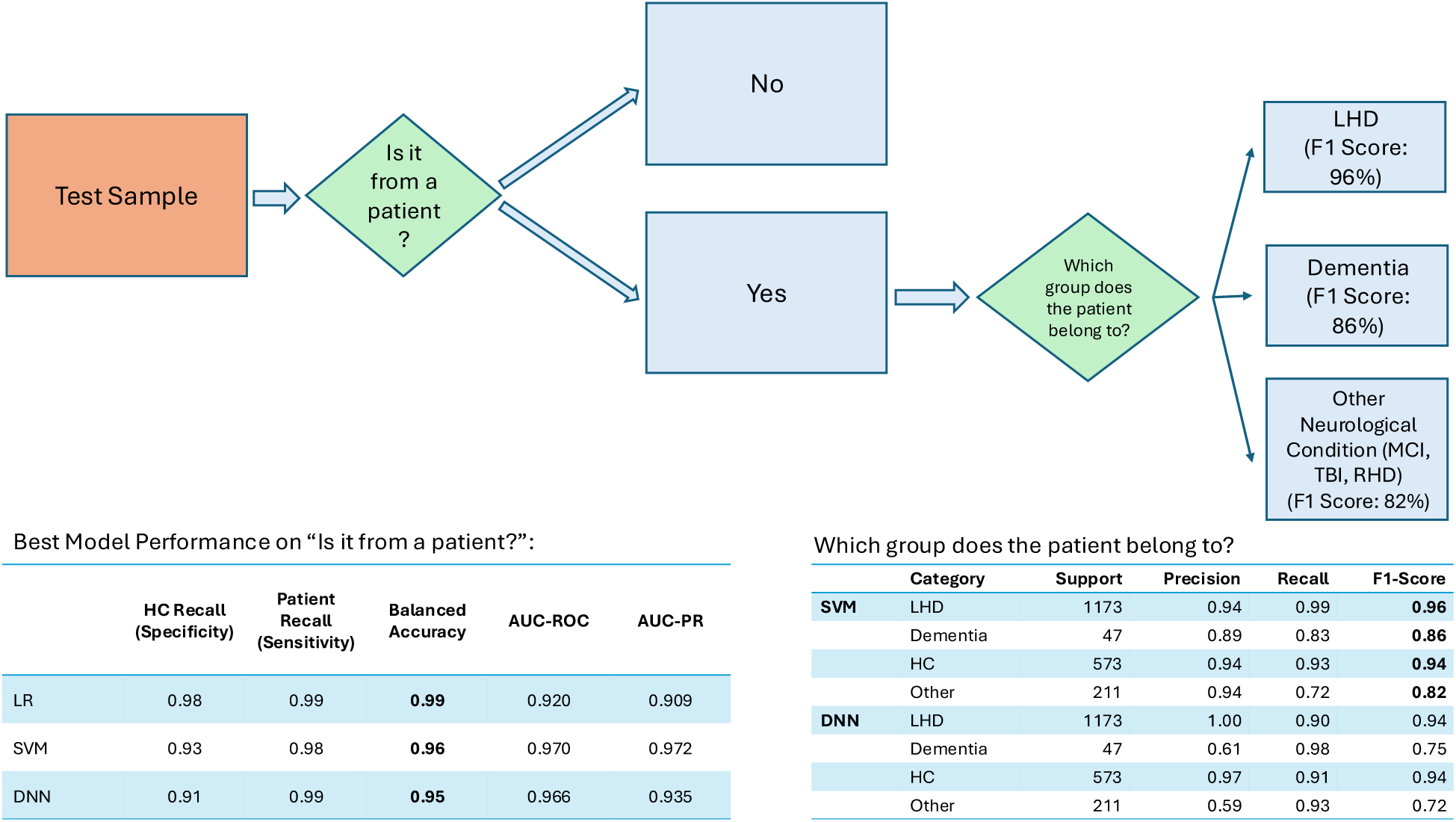
Two-stage hierarchical classification system for distinguishing neurological patients from healthy controls and subsequent patient subgroup classification. The flowchart illustrates a binary decision tree where test samples are first classified as either patient or healthy control (HC), followed by multi-class classification of patient samples into specific neurological conditions. The first stage achieves high performance with F1 scores of 96% for patient detection. Patients are subsequently classified into Left Hemisphere Damage (LHD, F1 = 96%), Dementia (F1 = 86%), or Other Neurological Conditions including Mild Cognitive Impairment (MCI), Traumatic Brain Injury (TBI), and Right Hemisphere Damage (RHD) (F1 = 82%).

**Table 7.** Table 5. Effect Size of Readability Measures Across Diagnostic Groups. This table presents the results of statistical analyses examining the differences in various readability measures across clinical diagnostic categories. Each row represents a separate readability metric, including both traditional indices (e.g., Smog Index, Gunning Fog Index) and computational linguistic measures (e.g., Yngve Load, Difficult Words). The table reports the F-statistic, numerator and denominator degrees of freedom (Num DF, Den DF), p-values (in scientific notation), and partial eta-squared (*η*²) as a measure of effect size.

| Measure | Category | F | Num DF | Den DF | *p* value | Partial $\eta^2$ |
| --- | --- | --- | --- | --- | --- | --- |
| Estimated Reading Time (sec) | 47.42 | 2.00 | 575.91 | 8.60E-20 | 0.14 | 0.16 |
| Smog Index | 86.37 | 2.00 | 1,085.65 | 1.55E-35 | 0.14 | 0.14 |
| Total Classical Yngve Load | 42.47 | 2.00 | 575.78 | 6.27E-18 | 0.13 | 0.15 |
| Difficult Words | 64.47 | 2.00 | 875.73 | 7.58E-27 | 0.13 | 0.20 |
| Gunning Fog Index | 27.21 | 2.00 | 1,036.36 | 3.04E-12 | 0.05 | 0.05 |
| Linsear Write Formula | 16.89 | 2.00 | 931.51 | 6.22E-08 | 0.03 | 0.08 |
| Dale Chall Readability Score | 14.23 | 2.00 | 981.68 | 8.06E-07 | 0.03 | 0.07 |

Readability findings indicate that neurological conditions, particularly those with LHD, result in the production of linguistically simpler text that requires lower reading levels and is more accessible to general audiences. This simplification likely reflects both the direct effects of language impairments and potential compensatory strategies employed by patients to maintain communicative effectiveness despite underlying linguistic deficits.

##### 3.5.5.1 Individuals with LHD

Left Hemisphere Damage (LHD) patients demonstrated the most pronounced shift toward simplified text readability across virtually all measures. Significant reductions in complexity were observed in the Automated Readability Index (*β* = –2.12, *p <* 0.001), Coleman Liau Index (*β* = – 3.21, *p <* 0.001), Flesch-Kincaid Grade Level (*β* = –1.38, *p <* 0.001), Gunning Fog Index (*β* = –1.50, *p <* 0.001), and SMOG Index (*β* = –1.22, *p <* 0.001). Conversely, the Flesch Reading Ease score increased significantly (*β* = 4.40, *p <* 0.001), indicating easier-to-read text. The Dale-Chall Readability Score showed a significant increase (*β* = 1.68, *p <* 0.001), suggesting greater use of common vocabulary. Estimated reading time was substantially reduced (−4.67 seconds, *p <* 0.001), reflecting shorter, simpler content.

##### 3.5.5.2 Individuals with Dementia

Individuals with dementia demonstrated moderate simplification across several readability measures. Significant decreases were observed in the Automated Readability Index (*β* = –0.43, *p <* 0.001), Coleman Liau Index (*β* = –0.47, *p =* 0.008), Flesch-Kincaid Grade Level (−0.22, *p =* 0.012), Gunning Fog Index (*β* = –0.22, *p =* 0.017), and SMOG Index (*β* = –0.35, *p <* 0.001). Flesch Reading Ease increased significantly (*β* = 1.06, *p =* 0.020). However, estimated reading time and difficult word usage showed no significant changes.

##### 3.5.5.3 Individuals with MCI

Individuals with MCI showed moderate readability changes with a mixed pattern. While some complexity measures decreased significantly, including the Coleman Liau Index (*β* = –0.40, *p =* 0.036) and Flesch-Kincaid Grade Level (*β* = –0.21, *p =* 0.020), others remained unchanged or showed minimal effects. Flesch Reading Ease increased (*β* = 0.97, *p =* 0.039), and difficult word usage decreased significantly (−7.40, *p =* 0.018). Estimated reading time was reduced by 3.39 seconds (*p <* 0.001). Interestingly, the Linsear Write Formula showed a small but significant increase (*β* = 0.20, *p =* 0.040).

##### 3.5.5.4 Individuals with TBI

Individuals with TBI exhibited substantial simplification in text complexity, though to a lesser degree than LHD patients. Significant decreases were found in the Automated Readability Index (*β* = –0.52, *p <* 0.001), Flesch-Kincaid Grade Level (*β* = –0.56, *p <* 0.001), Gunning Fog Index (*β* = –0.60, *p <* 0.001), and SMOG Index (*β* = –0.56, *p <* 0.001). Flesch Reading Ease increased significantly (*β* = 1.45, *p <* 0.001), while the Dale-Chall score increased (*β* = 0.46, *p*< 0.001), indicating simpler vocabulary usage. Estimated reading time decreased by 3.94 seconds (*p <* 0.001).

##### 3.5.5.5 Individuals with RHD

Individuals with individuals with RHD showed minimal changes in text readability, with most measures remaining within normal ranges. Only the Flesch Reading Ease showed a significant increase (*β* = 1.50, *p =* 0.038) and the Linsear Write Formula decreased slightly (*β* = –0.38, *p =* 0.012). This pattern suggests that text complexity remains largely preserved following right hemisphere injury.

#### 3.5.6 Semantic Markers (Named Entity Analysis)

The semantic findings reveal a markedly different pattern from phonological and syntactic results, with some patient groups showing compensatory increases in certain semantic categories. The preservation or enhancement of organization and person references in several groups suggests that semantic processing may be more resilient to neurological damage, with patients potentially shifting toward concrete, socially relevant semantic content when other linguistic abilities are compromised.

##### 3.5.6.1 Individuals with LHD

Individuals with LHD demonstrated a complex pattern of semantic changes characterized by both impairments and compensatory increases. Significant reductions were observed in cardinal number usage (count: –0.29, *p <* 0.001; ratio: –0.052, *p <* 0.001) and date references (count: –0.80, *p <* 0.001; ratio: –0.070, *p <* 0.001). However, individuals with LHD showed notable increases in organization references (count: +0.78, *p <* 0.001; ratio: +0.103, *p <* 0.001) and person reference ratios (+0.069, *p <* 0.001), suggesting possible compensatory semantic strategies or shifts toward more concrete, person-centered discourse.

##### 3.5.6.2 Individuals with Dementia

Individuals with dementia demonstrated mild semantic changes with the most notable effect being a significant reduction in cardinal number ratios (*β* = –0.060, *p <* 0.001) and date ratios (*β* = –0.030, *p =* 0.029). Interestingly, organization ratios showed a significant increase (*β* = 0.074, *p <* 0.001), potentially reflecting preserved or compensatory use of organizational semantic categories. Overall, the semantic profile in dementia appeared less severely affected than in other patient groups.

##### 3.5.6.3 Individuals with MCI

Individuals with MCI exhibited selective semantic difficulties with a distinctive pattern. Cardinal number ratios showed a significant increase (*β* = 0.132, *p <* 0.001), while date ratios decreased (*β* = –0.039, *p =* 0.003). Organization ratios were reduced (*β* = –0.054, *p =* 0.005), and person counts declined (*β* = –0.61, *p =* 0.038). This pattern suggests specific difficulties with temporal and organizational semantic categories while potentially overcompensating with numerical references.

##### 3.5.6.4 Individuals with TBI

Individuals with TBI showed moderate semantic impairments primarily in numerical and personal reference domains. Significant reductions included cardinal number counts (*β* = –0.45, *p =* 0.002) and ratios (−0.041, *p =* 0.008), as well as person counts (*β* = –0.57, *p =* 0.033). Date and organization references remained relatively preserved, indicating more selective semantic deficits compared to other linguistic domains.

##### 3.5.6.5 Individuals with RHD

Individuals with RHD showed minimal semantic impairments with only cardinal number counts significantly reduced (−0.53, *p =* 0.012) and organization counts increased (+1.06, *p =* 0.010). Most other semantic measures remained within normal ranges, suggesting that semantic processing is largely preserved following right hemisphere injury.

## 4 Discussion

Language is an extraordinarily complex a distributed network, interfacing with human faculties and cognitive processes such as memory, attention, executive functions, and emotions (Margulies & Petrides, 2013; Stanford & Delage, 2023; Themistocleous, 2025). Damage in brain areas responsible for language or areas affecting these cognitive systems is intrinsically reflected in an individual’s language (Hickok & Poeppel, 2007). An impaired cognitive function is often the earliest indication of neurological conditions, like mild cognitive impairment (MCI) and dementia, or can designate another acquired damage like left (LHD) and right hemisphere stroke (RHD), and traumatic brain injury (TBI) and can manifest as a subtle or severe change in linguistic expression, lexical choice, syntactic structure, acoustic properties, and discourse coherence. This makes speech and language a uniquely rich, non-invasive, and continuously available source of medical information, offering a veritable window into an individual’s brain health and cognitive status. The potential to harness this data for diagnostic and prognostic purposes is immense. Traditional approaches are time-consuming, require controlled clinical settings, and can be stressful to the patients. These drawbacks of traditional methods can be addressed by the recent advancements in Machine Learning (ML) and Natural Language Processing (NLP), demonstrating remarkable capabilities in pattern recognition, data analysis, and predictive modeling. In our previous research, we have already shown that ML techniques can enhance the diagnostic accuracy for neurodegenerative disorders by identifying complex patterns in clinical and neuroimaging data that often elude conventional analytical approaches (Kim et al., 2024; Themistocleous et al., 2018; Themistocleous, Ficek, et al., 2020; Themistocleous, Webster, et al., 2020). Additionally, this underscores the methodological capacity of AI algorithms to manage and interpret intricate medical data, a capability directly transferable to the complexities of speech. In this study, we employed NLP, ML, and robust statistical approach to extract relevant linguistic information and detect signatures for text productions of patients in a variety of discoursal tasks. That resulted into an analysis of 292 linguistic measures from distinct language domains.

### 4.1 Language Discriminates Diverse Neurological Conditions: NeuroScreen

Having a high-performing, end-to-end model is critical for its real-world usefulness in a clinical setting. The excellent performance metrics demonstrate that this system is not just a theoretical exercise but a potentially powerful diagnostic tool. The model’s ability to distinguish between patients and healthy controls with up to 99% accuracy is its most crucial feature. This near-perfect performance means the system can function as a reliable screening tool for early detection and characterization of neurological conditions. The model excels at identifying common and distinct conditions like LHD with a 96% F1 score and Dementia with an 88% F1 score. This provides a strong basis as a useful tool in the clinic to advice the diagnostic process. While the model struggled with rarer or less linguistically distinct conditions (like RHD and TBI), by grouping them into “Other Neurological Conditions” category offers a smart, practical approach as in this way the category still achieves a high F1 score of 82%. In this way the ML flags these patients for more specialized expert review without overstating its diagnostic precision.

Clinicians can trust the model to accurately flag individuals who need further evaluation, minimizing the chances of missing a patient with a neurological condition. It automates the initial assessment, saving valuable time for specialists and allowing healthcare systems to screen more people, more quickly. Beyond simply identifying a patient, the model’s strength lies in its ability to differentiate between specific neurological conditions. Knowing which condition a person has is essential for providing the right treatment. In essence, this two-stage, end-to-end performance creates a complete and practical workflow. It reliably filters the general population and then provides a highly accurate differential diagnosis for common conditions while intelligently triaging more complex cases. This makes the system on of the most powerful and scalable tool for clinical decision support (Ahmed et al., 2013; Fraser et al., 2019; Fraser et al., 2018; Kim et al., 2024; König et al., 2015; Lavoie et al., 2023; Themistocleous et al., 2018; Themistocleous, Eckerström, et al., 2020; Themistocleous, Ficek, et al., 2020; Tuomiranta et al., 2025).

The reasons are twofold, we rely on a large dataset and on the large number of computational measures that we have develop and provide within Open Brain AI (Themistocleous, 2024) covering a wide range of language domains spanning from textual *readability* (Dale & Chall, 1948; Fitzsimmons et al., 2010; Klare, 1974; Themistocleous, 2024), *Lexicon and Lexical Information* (Fergadiotis & and Wright, 2011), *Phonology* (Barbieri et al., 2018; Croot et al., 2000), *Morphology* (Badecker et al., 1990; Caramazza & Hillis, 1991; Fridriksson et al., 2018; Hillis, 1989; Hillis et al., 2018; Stockbridge, Matchin, et al., 2021), *Syntax* (Bastiaanse, 2013; Caramazza & Hillis, 1989; Mack et al., 2021; Thompson & Mack, 2014; Wilson et al., 2016), and *Semantics.* Finaly, this approach demonstrates the importance of these metrics to function as linguistic signatures indicating that symptoms associated with neurological conditions can both facilitate diagnosis and function as therapeutic targets. The characteristics of these language signatures and their patterns are discussed next.

### 4.2 Overall language characteristics

The findings revealed condition-specific distinct patterns of linguistic impairments. The most significant differences were observed in individuals with LH stroke and dementia, TBI, MCI, and finally RHD, which showed the most preserved language.

Concerning the lexical markers and the vocabulary usage, we found that individuals with LHD and TBI showed significant reductions in the number of words produced and lexical diversity. Patients with dementia also exhibited reduced word production and diversity, though to a lesser extent while patients with MCI and RHD lexical profile was closer to that of HCs. Concerning the phonological measurements, such as key syllable patterns and syllable complexity, patients with LHD, TBI, and dementia groups produced fewer words of varying syllable lengths and less complex syllable structures. Patients with RHD produced similar phonological patterns to HCs.

In addition to the lexicon and phonology, key morphological measures that involve both the distribution of part of speech (POS) production and inflectional morphology presented key differences among group in the distribution of these measures (Kiran, 2012; Kiran et al., 2014; Kiran et al., 2009; Thompson et al., 2003). Patients with LHD and TBI demonstrated widespread reductions in the use of most word classes, including determiners, adjectives, nouns, and verbs. Patients with dementia also showed a decline in the use of several word classes whereas patients with RHD showed relatively minor differences compared to HCs.

In line with earlier findings (den Ouden et al., 2016; Den Ouden et al., 2019; Thompson et al., 2013), syntactic complexity was significantly reduced in individuals with LHD and TBI, who produced shorter and structurally simpler sentences. Patients with dementia also showed notable reductions in syntactic complexity. The MCI group presented mostly reductions of the core syntactic measures whereas patients with RHD provided fewer distinct patterns compared to HCs.

The statistical models about the readability of the text, a novel measure that we employed in this study, reveal several important insights about the language production in the patient groups. Individuals with LHD, TBI, and dementia was generally rated as less complex and easier to read by various readability indices. Patients with LHD, TBI, and dementia groups used fewer named entities like cardinal numbers and dates.

### 4.3 Overall Patterns Across Diagnostic Groups

A global view of the results reveals distinct patterns of linguistic alteration across the diagnostic groups.

Expectedly, individuals with LHD consistently demonstrated the most extensive and pronounced differences from HCs across nearly all linguistic categories as detailed in the results section. The majority of these were characterized by significantly lower scores (negative estimates), particularly in measures of lexical production and diversity, morphological complexity, phonological output, and syntactic complexity. These findings corroborate our existing understanding about the grammatical difficulties (Matchin et al., 2014), reduced lexical diversity (Fergadiotis & and Wright, 2011), and impaired phonological output (Miceli et al., 1980), but at the same time they offer a broader understanding, given the extensive coverage our measures provide of the language domain andthe systematic integration of features spanning the entire linguistic hierarchy—from phonological structures to discourse-level semantics. Unlike traditional clinical assessments that typically focus on isolated linguistic domains (e.g., naming tests for semantics, sentence repetition for syntax), whereas this approach captures the complex interplay between linguistic levels that characterizes real-world communication.

Importantly, the results highlight previously underappreciated compensatory strategies, such as increased reliance on proper nouns, socially salient references (e.g., persons, organizations), and syntactic simplification through appositional and compound modifiers. This suggests that individuals with LHD are not merely producing less language but may be restructuring their output (whether consciously or unconsciously) to maximize communicative success within their impaired linguistic system. Furthermore, the readability metrics provide novel, ecologically relevant evidence that the language produced by individuals with LHD is objectively simpler and more accessible, supporting the interpretation that both deficits and adaptations co-occur in spontaneous language use.

Individuals with TBI also exhibited a broad range of significant differences from HCs, which lies upon with prior evidence that has also found reductions in linguistic output (e.g., total words, content words, unique words), complexity (e.g., Corrected TTR), and various syntactic counts (Coelho, 2016; Lê & Coelho, 2024; Marini et al., 2011). In several measures, the magnitude of these differences was comparable to or, in some specific instances, even exceeded those seen in dementia. At the same time, the TBI group displayed increased lexical diversity and preserved, or even compensatory, use of certain morphological and syntactic features, indicating strategic adaptations rather than uniform linguistic degradation. The semantic profile of TBI also revealed selective vulnerabilities, particularly in numerical and personal references, suggesting domain-specific disruptions in meaning construction rather than global semantic impairment. Importantly, the readability metrics demonstrate that language produced by individuals with TBI is objectively simplified, mirroring patterns seen in aphasia and underscoring the functional consequences of these linguistic changes for everyday communication. Together, these results contribute novel, objective evidence that TBI disrupts language in ways that are both overlapping with and distinct from classical aphasia profiles.

While global cognitive impairment is a hallmark of dementia, this study demonstrates that spontaneous language production in this group is relatively preserved across many core linguistic domains, particularly in phonology, syntax, and overall lexical productivity (this is the case in amnestic dementia, but not necessarily in primary progressive aphasia, which is not a syndrome studied her). However, subtle but meaningful disruptions emerged in specific areas which echo prior findings, notably reduced lexical diversity (Williams et al., 2023), simplified word choice (e.g., shorter average word length), and decreased use of complex syntactic and semantic structures (Le et al., 2011; Meteyard et al., 2014; Snowdon, 2002). The readability findings further underscore this pattern, showing a moderate shift toward simpler, more accessible language that likely reflects both cognitive decline and simplification strategies. While the pattern was generally one of decreased scores compared to HCs, the effects were often less pronounced and less uniformly distributed across measures compared to the LHD group, reflecting high variation in this group (Le et al., 2011; Meteyard et al., 2014; Snowdon, 2002).

Unlike LHD aphasia or TBI, MCI was characterized by a subtler but systematic pattern of linguistic simplification, which has been shown previously, particularly evident in reduced lexical productivity, decreased syntactic complexity, and phonological impairments (Kim et al., 2019; Sanborn et al., 2022; Sung et al., 2020). The findings reveal that even at this early disease stage, individuals with MCI produced fewer total words, content words, and unique word types, accompanied by reductions in sentence length and the use of complex syntactic structures such as dependent clauses and prepositional phrases. Interestingly, lexical diversity (standard TTR) was increased compared to HCs, reflecting a compensatory pattern where speakers produce fewer words overall but rely on a more varied vocabulary within their reduced output. Readability metrics further indicated that MCI speakers produce objectively simpler, more accessible language than HCs, likely reflecting both cognitive constraints and emerging compensatory strategies.

These results provide new, quantitative evidence reinforcing and extending long-standing but often inconsistently documented observations that language production following RHD is relatively preserved in terms of core linguistic structure, but may still exhibit subtle disruptions, particularly in semantic, pragmatic, and higher-order discourse features. The present analyses reveal that individuals with RHD performed comparably to healthy controls across most lexical, phonological, morphological, syntactic, and readability measures, supporting prior research showing that RHD does not typically produce the overt language breakdown observed in left hemisphere stroke or TBI. However, the detection of reduced use of specific structures, such as comparative adjectives, complex syllable patterns, and second-person pronouns, along with a selective reduction in certain semantic categories (e.g., cardinal numbers), highlights that RHD may subtly affect aspects of language tied to complexity, perspective-taking, or relational meaning. These findings align with previous evidence that while RHD does not result in classical aphasia, it can impact elements of discourse organization, inferencing, and pragmatic language, often in ways that evade detection by standard language batteries.

A key insight from these findings is that while language simplification emerges as a common consequence of neurological damage, the specific linguistic signature varies systematically across disorders, reflecting both the nature of the underlying neural disruption and the ways in which language production shifts in response to these deficits. Across conditions such as LHD, TBI, MCI, and dementia, individuals consistently produced simpler language characterized by reduced lexical output, diminished syntactic complexity, and lower readability. Yet, the precise linguistic domains affected, and the nature of these changes differed. For example, individuals with MCI and TBI showed increased lexical diversity within reduced output, while LHD and dementia speakers exhibited greater reliance on proper nouns and socially salient references. These patterns suggest that language production does not decline uniformly but instead reflects a combination of impairment and adaptive linguistic shifts, whether conscious or automatic. Even in the context of cognitive or neural decline, measurable alterations in language use indicate preserved linguistic capacity and potential compensatory processes. Capturing both these deficits and adaptations provides a more complete and clinically informative picture of how language reflects the complex interaction between neural damage, cognitive constraints, and preserved linguistic mechanisms across neurological conditions.

### 4.4 Limitations and Future Research

Although this study marks a critical starting point for comparing more than one and especially often conditions that are dissimilar in their underlying pathology making this comparison possible there are several that are inherent to this approach. First, for many neurological conditions, especially rare disorders or the initial stages of more common ones like MCI, large-scale speech datasets are lacking, especially for languages other than English, so shared corpora like DementiaBank and TalkBank are crucial.

A second issue is the need for more fine-grained distinctions between the populations. Although the categories we have presented here like LHD, or dementia correspond to a broader diagnosis, there is an important variation within the population because of their condition, the potential influence of medication and other comorbidities on the linguistic profiles. So, there is a need for a greater understanding through subtyping the populations into subgroups, like individuals with anomic aphasia and conduction aphasia and individuals with different severity levels.

Understanding disease progression and the evolution of linguistic signatures over time necessitates longitudinal data collection, where individuals are assessed repeatedly. Such data, as used in the MCI-to-AD progression study, is invaluable but expensive and time-consuming to acquire. The noted lack of longitudinal AD speech data, particularly at the MCI stage, and DementiaBank’s aim for longitudinal tracking highlight this ongoing need.

A key limitation of the current study is that we collapsed language data across multiple discourse tasks, despite well-established evidence that different tasks elicit distinct linguistic profiles (Stark & Fukuyama, 2021; Stark Brielle, 2019). While this approach maximized statistical power and facilitated broad comparisons across diagnostic groups, it may have obscured task-specific linguistic patterns that are clinically and theoretically meaningful. We have planned for future work that will systematically examine how task type interacts with diagnosis to influence linguistic profiles.

Finally, there is an increasing need to have acoustic data along with linguistic data. For example, previous research has shown that the extension of information provided be even a single sound is incredible. As we have learnt from our research, the way speakers pronounce their vowels (Themistocleous, 2017), consonants (Themistocleous, 2019), voice quality and prosody (Themistocleous, Eckerström, et al., 2020) reveal aspects of speakers’ identity, like their dialects, sociolects and pathology. Our future research will intergrade these different concepts together and provide multimodal systems for understanding language and cognition. Future research should also prioritize the continued expansion of this dataset, enhancing its diversity and generalizability. Integrating multimodal signatures, such as neuroimaging data, alongside these linguistic measures will be the next frontier, promising even greater precision and clinical utility. Ultimately, this open library provides the essential groundwork for a future where language analysis is a core component of neurological care.

### 4.5 Conclusion

This study represents a critical step toward transforming language analysis from a research tool into a scalable, clinically actionable digital biomarker for neurological disorders. By applying automated, computational linguistic analysis to one of the largest and most diverse databases of spoken language, we demonstrate that distinct, quantifiable linguistic profiles can differentiate between individuals with left hemisphere damage, right hemisphere damage, dementia, MCI, TBI, and healthy controls. These findings not only advance scientific understanding of language impairments but also establish a practical foundation for integrating language-based digital biomarkers into routine neurological assessment.

Importantly, the architecture of Open Brain AI provides a clear pathway for translation beyond the research setting. With further development, this platform could be scaled into an accessible, secure application deployable by researchers, speech-language pathologists, and clinicians worldwide. Such a tool could enable real-time, automated language analysis in clinical environments, telemedicine, or even remote monitoring contexts—delivering objective, reproducible language metrics that augment clinical decision-making. The naturalistic, low-burden nature of speech samples makes this approach uniquely suited to scalable, patient-friendly assessment.

Looking ahead, the integration of Open Brain AI into clinical workflows, combined with regulatory-compliant development and continued dataset expansion, holds the potential to redefine how language is used to detect, monitor, and personalize care for individuals with neurological conditions. By moving beyond proof-of-concept and toward scalable, validated tools, this work contributes to the broader goal of leveraging AI and language as accessible, ecologically valid biomarkers in digital medicine.

## Supplementary Information

### Author Contributions

CT conducted the statistical analyses, developed the analysis code and software, wrote the first draft of the manuscript, and contributed to subsequent manuscript editing. BS contributed to data curation and reviewed and edited the manuscript. All authors reviewed and approved the final version of the manuscript.

### Data Availability

The study used openly available human data that were originally located at TalkBank (https://talkbank.org). The analysis included individual-level raw behavioral data in the form of transcripts from patients with acquired neurological conditions. All individual-level data were fully de-identified prior to analysis and prior to inclusion in the TalkBank library. The raw behavioral data are available through membership to the TalkBank consortium. Additional data used in this study are available from the authors upon request with a proper Data Use Agreement in place. The analysis code is openly available at https://github.com/themistocleous/neuroscreen.

## Ethics Statement

This work used only existing, publicly available de-identified data from TalkBank. Therefore, no approval or waiver from an institutional review board (IRB) was required.

## References

1. Abadi, M., Agarwal, A., Barham, P., Brevdo, E., Chen, Z., Citro, C., Corrado, G. S., Davis, A., Dean, J., Devin, M., Ghemawat, S., Goodfellow, I. J., Harp, A., Irving, G., Isard, M., Jia, Y., Józefowicz, R., Kaiser, L., Kudlur, M.,…Zheng, X. (2016). *TensorFlow: Large-Scale Machine Learning on Heterogeneous Distributed Systems* 12th USENIX Symposium on Operating Systems Design and Implementation (OSDI’16), Savannah, GA. https://www.usenix.org/conference/osdi16/technical-sessions/presentation/abadi

2. Ahmed, S., Haigh, A.-M. F., de Jager, C. A., & Garrard, P. (2013). Connected speech as a marker of disease progression in autopsy-proven Alzheimer’s disease. Brain, 136(12), 3727–3737. 10.1093/brain/awt269

3. Badecker, W., Hillis, A., & Caramazza, A. (1990). Lexical morphology and its role in the writing process: evidence from a case of acquired dysgraphia. Cognition, 35(3), 205–243.

4. Barbieri, Z., Fernández, M. A., Newbury, D. F., & Villanueva, P. (2018). Family aggregation of language impairment in an isolated Chilean population from Robinson Crusoe Island. Int J Lang Commun Disord, 53(3), 643–655. 10.1111/1460-6984.12377

5. Bastiaanse, R. (2013). Why reference to the past is difficult for agrammatic speakers [Conference Paper]. Clinical Linguistics & Phonetics, 27(4), 244–263. 10.3109/02699206.2012.751626

6. Becker, J. T., Boiler, F., Lopez, O. L., Saxton, J., & McGonigle, K. L. (1994). The natural history of Alzheimer’s disease: description of study cohort and accuracy of diagnosis. Archives of Neurology, 51(6), 585–594. https://jamanetwork.com/journals/jamaneurology/article-abstract/592905

7. Beltrami, D., Gagliardi, G., Rossini Favretti, R., Ghidoni, E., Tamburini, F., & Calzà, L. (2018). Speech Analysis by Natural Language Processing Techniques: A Possible Tool for Very Early Detection of Cognitive Decline? [Original Research]. Frontiers in Aging Neuroscience, Volume 10 – 2018. 10.3389/fnagi.2018.00369

8. Birch, E. S., C., S. B., & and Neumann, D. (2024). Factors related to social inferencing performance in moderate-severe, chronic TBI. Brain Injury, 38(12), 992–1003. 10.1080/02699052.2024.2361634

9. Breiman, L. (2001). Random Forests. Machine Learning, 45(1), 5–32. 10.1023/A:1010933404324

10. Caplan, D. (1987). Neurolinguistics and linguistic aphasiology: an introduction. Cambridge University Press. Sample text http://www.loc.gov/catdir/samples/cam034/86031049.html

11. Publisher description http://www.loc.gov/catdir/description/cam023/86031049.html

12. Table of contents http://www.loc.gov/catdir/toc/cam024/86031049.html

13. Caramazza, A., & Hillis, A. E. (1989). The disruption of sentence production: some dissociations. Brain and Language, 36(4), 625–650.

14. Caramazza, A., & Hillis, A. E. (1991). Lexical organization of nouns and verbs in the brain. Nature, 349(6312), 788–790.

15. Chawla, N. V., Bowyer, K. W., Hall, L. O., & Kegelmeyer, W. P. (2002). SMOTE: synthetic minority over-sampling technique. Journal of artificial intelligence research, 16, 321–357.

16. Ciesielska, N., Sokołowski, R., Mazur, E., Podhorecka, M., Polak-Szabela, A., & Kędziora-Kornatowska, K. (2016). Is the Montreal Cognitive Assessment (MoCA) test better suited than the Mini-Mental State Examination (MMSE) in mild cognitive impairment (MCI) detection among people aged over 60? Meta-analysis. Psychiatr Pol, 50(5), 1039–1052.

17. Coelho, C. A. (2016). Discourse analysis in traumatic brain injury. In Communication disorders following traumatic brain injury (pp. 55–79). Psychology press.

18. Cortes, C., & Vapnik, V. (1995). Support-Vector Networks. Machine Learning, 20.

19. Croot, K., Hodges, J. R., Xuereb, J., & Patterson, K. (2000). Phonological and Articulatory Impairment in Alzheimer’s Disease: A Case Series. Brain and Language, 75(2), 277–309. 10.1006/brln.2000.2357

20. Dale, E., & Chall, J. S. (1948). A formula for predicting readability: Instructions. Educational research bulletin, 37-54.

21. Davis, D. H. J., Creavin, S. T., Yip, J. L. Y., Noel-Storr, A. H., Brayne, C., & Cullum, S. (2021). Montreal Cognitive Assessment for the detection of dementia. Cochrane Database of Systematic Reviews (7). 10.1002/14651858.CD010775.pub3

22. den Ouden, D. B., Dickey, M. W., Anderson, C., & Christianson, K. (2016). Neural correlates of early-closure garden-path processing: Effects of prosody and plausibility. Q J Exp Psychol (Hove), 69(5), 926–949. 10.1080/17470218.2015.1028416

23. Den Ouden, D. B., Malyutina, S., Basilakos, A., Bonilha, L., Gleichgerrcht, E., Yourganov, G., & Fridriksson, J. (2019). Cortical and structural connectivity damage correlated with impaired syntactic processing in aphasia. Human Brain Mapping, 40(7), 2153–2173. 10.1002/hbm.24514

24. Faroqi-Shah, Y., Treanor, A., Ratner, N. B., Ficek, B., Webster, K., & Tsapkini, K. (2020). Using narratives in differential diagnosis of neurodegenerative syndromes. J Commun Disord, 85, 1–13. 10.1016/j.jcomdis.2020.105994

25. Fergadiotis, G., & and Wright, H. H. (2011). Lexical diversity for adults with and without aphasia across discourse elicitation tasks. Aphasiology, 25(11), 1414–1430. 10.1080/02687038.2011.603898

26. Fitzsimmons, P. R., Michael, B. D., Hulley, J. L., & Scott, G. O. (2010). A readability assessment of online Parkinson’s disease information. J R Coll Physicians Edinb, 40(4), 292–296. 10.4997/JRCPE.2010.401

27. Fraser, K. C., Linz, N., Li, B., Lundholm Fors, K., Rudzicz, F., Konig, A., Alexandersson, J., Robert, P., & Kokkinakis, D. (2019). Multilingual prediction of {A}lzheimer{’}s disease through domain adaptation and concept-based language modelling. Proceedings of the 2019 Conference of the North {A}merican Chapter of the Association for Computational Linguistics: Human Language Technologies, Volume 1 (Long and Short Papers), 3659–3670. https://www.aclweb.org/anthology/N19-1367

28. Fraser, K. C., Lundholm Fors, K., Eckerström, M., Themistocleous, C., & Kokkinakis, D. (2018). Improving the Sensitivity and Specificity of MCI Screening with Linguistic Information. Proceedings of the LREC 2018 Workshop “Resources and ProcessIng of linguistic, para-linguistic and extra-linguistic Data from people with various forms of cognitive/psychiatric impairments (RaPID-2)”(2015), 19–26. http://demo.spraakdata.gu.se/svedk/pbl/rapid-2-final.pdf%0Ahttp://lrec-conf.org/workshops/lrec2018/W31/pdf/3_W31.pdf

29. Fraser, K. C., Meltzer, J. A., & Rudzicz, F. (2015). Linguistic Features Identify Alzheimer’s Disease in Narrative Speech. Journal of Alzheimer’s Disease, 49(2), 407–422. 10.3233/JAD-150520

30. Frattali, C., American, S., & Hearing, A. (1995). Functional Assessment of Communication Skills for Adults: ASHA FACS. In (pp. 1 text (118 pages: illustrations; 128 cm), demographic section booklet ([112] pages; 128 cm), rating key scale, 111 computer disc (113 111/112 in.), license agreement). Rockville, MD: ASHA.

31. Fridriksson, J., den Ouden, D. B., Hillis, A. E., Hickok, G., Rorden, C., Basilakos, A., & Bonilha, L. (2018). Anatomy of aphasia revisited. Brain, 141(3), 848–862. 10.1093/brain/awx363

32. Goodglass, H. (1993). Understanding aphasia. Academic Press.

33. Goodglass, H., & Kaplan, E. (1979). Assessment of cognitive deficit in the brain-injured patient. In Neuropsychology (pp. 3–22). Springer.

34. Goodglass, H., & Kaplan, E. (1983). Boston diagnostic aphasia examination (BDAE). Lea & Febiger.

35. Goodglass, H., Kaplan, E., & Barresi, B. (2001). BDAE-3: Boston Diagnostic Aphasia Examination–Third Edition. Lippincott Williams & Wilkins Philadelphia, PA.

36. Hartig, F. (2016). DHARMa: residual diagnostics for hierarchical (multi-level/mixed) regression models. CRAN: Contributed Packages.

37. Hastie, T., Tibshirani, R., & Friedman, J. (2009). The Elements of Statistical Learning: Data Mining, Inference, and Prediction. (2 ed.). Springer.

38. Hickok, G., & Poeppel, D. (2007). The cortical organization of speech processing. Nature Reviews Neuroscience, 8, 393–402. https://www.nature.com/articles/nrn2113.pdf

39. Hillis, A. E. (1989). Efficacy and generalization of treatment for aphasic naming errors. Archives of Physical Medicine and Rehabilitation, 70(8), 632–636.

40. Hillis, A. E., Beh, Y. Y., Sebastian, R., Breining, B., Tippett, D. C., Wright, A., Saxena, S., Rorden, C., Bonilha, L., & Basilakos, A. (2018). Predicting Recovery in Acute Post-stroke Aphasia. Annals of Neurology.

41. Holland, A. L., Wozniak, L., & Fromm, D. (2018). CADL-3: communication activities of daily living (Third edition ed.). Pro-Ed.

42. Hornstein, N., Nunes, J., & Grohmann, K. K. K. K. (2005). Understanding minimalism. 405.

43. Joanette, Y., Ferré, P., & Wilson, M. A. (2015). Right hemisphere damage and communication. In The Cambridge Handbook of Communication Disorders (pp. 247–265).

44. Kaplan, E., Goodglass, H., & Weintraub, S. (2001). Boston Naming Test. Pro-ed.

45. Kertesz, A. (2006). Western aphasia battery-revised (WAB-R). Pearson.

46. Kim, B. S., Kim, Y. B., & Kim, H. (2019). Discourse measures to differentiate between mild cognitive impairment and healthy aging. Frontiers in Aging Neuroscience, 11, 221. https://www.frontiersin.org/journals/aging-neuroscience/articles/10.3389/fnagi.2019.00221/pdf

47. Kim, H., Hillis, A. E., & Themistocleous, C. (2024). Machine Learning Classification of Patients with Amnestic Mild Cognitive Impairment and Non-Amnestic Mild Cognitive Impairment from Written Picture Description Tasks. Brain Sciences, 14(7).

48. Kiran, S. (2012). What is the nature of poststroke language recovery and reorganization? ISRN neurology, 2012, 786872.

49. Kiran, S., Balachandran, I., & Lucas, J. (2014). The nature of lexical-semantic access in bilingual aphasia. Behavioural Neurology, 2014, 389565.

50. Kiran, S., Sandberg, C., & Abbott, K. (2009). Treatment for lexical retrieval using abstract and concrete words in persons with aphasia: Effect of complexity. Aphasiology, 23(7), 835–853.

51. Klare, G. R. (1974). Assessing Readability. Reading Research Quarterly, 10(1), 62–102. 10.2307/747086

52. Koller, M. (2016). robustlmm: an R package for robust estimation of linear mixed-effects models. Journal of Statistical Software, 75, 1–24.

53. König, A., Satt, A., Sorin, A., Hoory, R., Toledo-Ronen, O., Derreumaux, A., Manera, V., Verhey, F., Aalten, P., Robert, P. H., & David, R. (2015). Automatic speech analysis for the assessment of patients with predementia and Alzheimer’s disease. *Alzheimer’s and Dementia: Diagnosis*, Assessment and Disease Monitoring, 1(1), 112–124. 10.1016/j.dadm.2014.11.012

54. Krakauer, J. W., & Carmichael, S. T. (2017). Broken movement: the neurobiology of motor recovery after stroke. The MIT Press.

55. Kuznetsova, A., Bruun Brockhoff, P., & Haubo Bojesen Christensen, R. (2016). lmerTest: Tests in Linear Mixed Effects Models. In R Foundation for Statistical Computing. https://CRAN.R-project.org/package=lmerTest

56. Lanzi Alyssa, M., Saylor Anna, K., Fromm, D., Liu, H., MacWhinney, B., & Cohen Matthew, L. (2023). DementiaBank: Theoretical Rationale, Protocol, and Illustrative Analyses. American Journal of Speech-Language Pathology, 32(2), 426–438. 10.1044/2022_AJSLP-22-00281

57. Lavoie, M., E., B. S., F., T.-W. D., L., G. N., S., S., M., F., C., L., & and Rochon, E. (2023). Longitudinal changes in connected speech over a one-year span in the nonfluent/agrammatic variant of Primary Progressive Aphasia. Aphasiology, 37(8), 1186–1197. 10.1080/02687038.2022.2084707

58. Lê, K., & Coelho, C. (2024). Discourse characteristics in Traumatic Brain Injury. In Spoken Discourse Impairments in the Neurogenic Populations: A State-of-the-Art, Contemporary Approach (pp. 65–80). Springer.

59. Le, X., Lancashire, I., Hirst, G., & Jokel, R. (2011). Longitudinal detection of dementia through lexical and syntactic changes in writing: a case study of three British novelists. Literary and Linguistic Computing, 26(4), 435–461. 10.1093/llc/fqr013

60. Lemaître, G., Nogueira, F., & Aridas, C. K. (2017). Imbalanced-learn: a python toolbox to tackle the curse of imbalanced datasets in machine learning. J. Mach. Learn. Res., 18(1), 559–563.

61. Levinson, S. C. (1983). Pragmatics. Cambridge University Press.

62. Lezak, M. D. (1995). Neuropsychological assessment (Vol. null).

63. Lyketsos, C. G., Lopez, O., Jones, B., Fitzpatrick, A. L., Breitner, J., & DeKosky, S. (2002). Prevalence of Neuropsychiatric Symptoms in Dementia and Mild Cognitive ImpairmentResults From the Cardiovascular Health Study. JAMA, 288(12), 1475–1483. 10.1001/jama.288.12.1475

64. Mack, J. E., Barbieri, E., Weintraub, S., Mesulam, M. M., & Thompson, C. K. (2021). Quantifying grammatical impairments in primary progressive aphasia: Structured language tests and narrative language production. Neuropsychologia, 151, 107713. 10.1016/j.neuropsychologia.2020.107713

65. MacWhinney, B. (2025). Understanding Language Through TalkBank. Current Directions in Psychological Science, 09637214241304345.

66. Margulies, D. S., & Petrides, M. (2013). Distinct parietal and temporal connectivity profiles of ventrolateral frontal areas involved in language production. The Journal of neuroscience: the official journal of the Society for Neuroscience, 33(42), 16846–16852. 10.1523/JNEUROSCI.2259-13.2013

67. Marini, A., Galetto, V., Zampieri, E., Vorano, L., Zettin, M., & Carlomagno, S. (2011). Narrative language in traumatic brain injury. Neuropsychologia, 49(10), 2904–2910. https://www.sciencedirect.com/science/article/abs/pii/S0028393211002983?via%3Dihub

68. Matchin, W., Sprouse, J., & Hickok, G. (2014). A structural distance effect for backward anaphora in Broca’s area: An fMRI study. Brain and Language, 138, 1–11. http://www.scopus.com/inward/record.url?eid=2-s2.0-84907857117&partnerID=40&md5=f5743ec29334a7384d0ddb8a2b70a0a0, http://ac.els-cdn.com/S0093934X14001278/1-s2.0-S0093934X14001278-main.pdf?_tid=35a12192-b903-11e4-a4ad-00000aacb35d&acdnat=1424438596_c71f1fca7f2c0087ec91c6af64302ccd

69. McKinney, W. (2010). Data Structures for Statistical Computing in Python. 10.25080/Majora-92bf1922-00a

70. Mesulam, M. M. (1982). Slowly progressive aphasia without generalized dementia. Annals of Neurology, 11(6), 592–598. https://onlinelibrary.wiley.com/doi/pdfdirect/10.1002/ana.410110607?download=t rue

71. Meteyard, L., Quain, E., & Patterson, K. (2014). Ever decreasing circles: Speech production in semantic dementia. Cortex, 55, 17–29. https://www.sciencedirect.com/science/article/abs/pii/S0010945213000695?via%3Dihub

72. Miceli, G., Gainotti, G., Caltagirone, C., & Masullo, C. (1980). Some aspects of phonological impairment in aphasia. Brain and Language, 11(1), 159–169. https://www.sciencedirect.com/science/article/abs/pii/0093934X80901170?via%3Dihub

73. Minga, J., Sheppard, S. M., Johnson, M., Hewetson, R., Cornwell, P., & Blake, M. L. (2023). Apragmatism: The renewal of a label for communication disorders associated with right hemisphere brain damage. International Journal of Language & Communication Disorders, 58(2), 651–666. 10.1111/1460-6984.12807

74. Minga, J., Stockbridge Melissa, D., Durfee, A., & Johnson, M. (2022). Clinical Guidelines for Eliciting Discourse Using the RHDBank Protocol. American Journal of Speech-Language Pathology, 31(5), 1949–1962. 10.1044/2022_AJSLP-22-00097

75. Mueller, K. D., Bruce, H., Jonilda, M., & and Turkstra, L. S. (2018). Connected speech and language in mild cognitive impairment and Alzheimer’s disease: A review of picture description tasks. Journal of Clinical and Experimental Neuropsychology, 40(9), 917–939. 10.1080/13803395.2018.1446513

76. Nivre, J., de Marneffe, M.-C., Ginter, F., Hajič, J., Manning, C. D., Pyysalo, S., Schuster, S., Tyers, F., & Zeman, D. (2020, May). Universal Dependencies v2: An Evergrowing Multilingual Treebank Collection. In N. Calzolari, F. Béchet, P. Blache, K. Choukri, C. Cieri, T. Declerck, S. Goggi, H. Isahara, B. Maegaard, J. Mariani, H. Mazo, A. Moreno, J. Odijk, & S. Piperidis, Proceedings of the Twelfth Language Resources and Evaluation Conference Marseille, France.

77. Nordlund, A., Rolstad, S., Hellström, P., Sjögren, M., Hansen, S., & Wallin, A. (2005). The Goteborg MCI study: mild cognitive impairment is a heterogeneous condition. *Journal of Neurology*, Neurosurgery & Psychiatry, 76(11), 1485–1490.

78. Obler, L. K., Fein, D., Nicholas, M., & Albert, M. L. (1991). Auditory comprehension and aging: Decline in syntactic processing. Applied Psycholinguistics, 12(4), 433–452. 10.1017/S0142716400005865

79. Park, K. W., Kim, H. S., Cheon, S.-M., Cha, J.-K., Kim, S.-H., & Kim, J. W. (2011). Dementia with Lewy Bodies versus Alzheimer’s Disease and Parkinson’s Disease Dementia: A Comparison of Cognitive Profiles. J Clin Neurol, 7(1), 19–24. 10.3988/jcn.2011.7.1.19, https://www.ncbi.nlm.nih.gov/pmc/articles/PMC3079155/pdf/jcn-7-19.pdf

80. Patel, S., Oishi, K., Wright, A., Sutherland-Foggio, H., Saxena, S., Sheppard, S. M., & Hillis, A. E. (2018). Right hemisphere regions critical for expression of emotion Through Prosody. Frontiers in Neurology, 9, 224. https://www.ncbi.nlm.nih.gov/pmc/articles/PMC5897518/pdf/fneur-09-00224.pdf

81. Pedregosa, F., Varoquaux, G., Gramfort, A., Michel, V., Thirion, B., Grisel, O., Blondel, M., Prettenhofer, P., Weiss, R., Dubourg, V., Vanderplas, J., Passos, A., Cournapeau, D., Brucher, M., Perrot, M., & Duchesnay, E. (2011). Scikit-learn: Machine Learning in Python. Journal of Machine Learning Research, 12, 2825–2830.

82. Petersen, R. C., Smith, G. E., Waring, S. C., Ivnik, R. J., Tangalos, E. G., & Kokmen, E. (1999). Mild cognitive impairment: clinical characterization and outcome. Archives of Neurology, 56(3), 303–308.

83. Petersen, R. C., Stevens, J. C., Ganguli, M., Tangalos, E. G., Cummings, J. L., & DeKosky, S. T. (2001). Practice parameter: Early detection of dementia: Mild cognitive impairment (an evidence-based review) – Report of the Quality Standards Subcommittee of the American Academy of Neurology. Neurology, 56. 10.1212/WNL.56.9.1133

84. Poeppel, D., Idsardi, W. J., & Van Wassenhove, V. (2008). Speech perception at the interface of neurobiology and linguistics [Review]. Philosophical Transactions of the Royal Society B: Biological Sciences, 363(1493), 1071–1086. 10.1098/rstb.2007.2160

85. R Core Team. (2025). R: A Language and Environment for Statistical Computing. In R Foundation for Statistical Computing. https://www.R-project.org/

86. Riès, S. K., Dronkers, N. F., & Knight, R. T. (2016). Choosing words: Left hemisphere, right hemisphere, or both? Perspective on the lateralization of word retrieval [Article]. Annals of the New York Academy of Sciences, 1369(1), 111–131. 10.1111/nyas.12993

87. Roach, A., Schwartz, M. F., Martin, N., Grewal, R. S., & Brecher, A. (1996). The Philadelphia naming test: scoring and rationale. Clinical Aphasiology, 24, 121–133.

88. Russell, L. (2020). emmeans: Estimated Marginal Means, aka Least-Squares Means. In R Foundation for Statistical Computing. https://CRAN.R-project.org/package=emmeans

89. Sanborn, V., Ostrand, R., Ciesla, J., & Gunstad, J. (2022). Automated assessment of speech production and prediction of MCI in older adults. Applied Neuropsychology: Adult, 29(5), 1250–1257. https://pmc.ncbi.nlm.nih.gov/articles/PMC8243401/

90. Sebastian, R., Thompson, C. B., Wang, N. Y., Wright, A., Meyer, A., Friedman, R. B., Hillis, A. E., & Tippett, D. C. (2018). Patterns of Decline in Naming and Semantic Knowledge in Primary Progressive Aphasia. Aphasiology, 32(9), 1010–1030. 10.1080/02687038.2018.1490388

91. Sidtis, D. L., & Yang, S. Y. (2017). Formulaic language performance in left– and right-hemisphere damaged patients: structured testing. Aphasiology, 31(1), 82–99. https://www.scopus.com/inward/record.uri?eid=2-s2.0-84961233762&doi=10.1080%2f026870382016.1157136&partnerID=40&md5=1c47d53601059c17c1cf48c5ecf28e7f

92. Snowdon, D. (2002). Aging with grace: What the nun study teaches us about leading longer, healthier, and more meaningful lives. Bantam.

93. Stanford, E., & Delage, H. (2023). The language-cognition interface in atypical development: Support for an integrative approach. Folia phoniatrica et logopaedica, 1–1. 10.1159/000533685

94. Stark, B. C., Alexander, J. M., Hittson, A., Doub, A., Igleheart, M., Streander, T., & Jewell, E. (2023). Test–retest reliability of microlinguistic information derived from spoken discourse in persons with chronic aphasia. Journal of Speech, Language, and Hearing Research, 66(7), 2316–2345.

95. Stark, B. C., Dalton, S. G., & Lanzi, A. M. (2025). Access to context-specific lexical-semantic information during discourse tasks differentiates speakers with latent aphasia, mild cognitive impairment, and cognitively healthy adults [Original Research]. Frontiers in Human Neuroscience, Volume 18 – 2024. 10.3389/fnhum.2024.1500735

96. Stark, B. C., Dutta, M., Murray Laura, L., Fromm, D., Bryant, L., Harmon Tyson, G., Ramage Amy, E., & Roberts Angela, C. (2021). Spoken Discourse Assessment and Analysis in Aphasia: An International Survey of Current Practices. Journal of Speech, Language, and Hearing Research, 64(11), 4366–4389. 10.1044/2021_JSLHR-20-00708

97. Stark, B. C., & Fukuyama, J. (2021). Leveraging big data to understand the interaction of task and language during monologic spoken discourse in speakers with and without aphasia. Language, Cognition and Neuroscience, 36(5), 562–585.

98. Stark Brielle, C. (2019). A Comparison of Three Discourse Elicitation Methods in Aphasia and Age-Matched Adults: Implications for Language Assessment and Outcome. American Journal of Speech-Language Pathology, 28(3), 1067–1083. 10.1044/2019_AJSLP-18-0265

99. Stockbridge, M. D., Matchin, W., Walker, A., Breining, B., Fridriksson, J., Hickok, G., & Hillis, A. E. (2021). One cat, Two cats, Red cat, Blue cats: Eliciting morphemes from individuals with primary progressive aphasia. Aphasiology, 35(12), 1–12. 10.1080/02687038.2020.1852167

100. Stockbridge, M. D., Sheppard, S. M., Keator, L. M., Murray, L. L., Lehman Blake, M., Right Hemisphere Disorders working group, E.-B. C. R. C. A. o. N. C. D., & Sciences. (2021). Aprosodia Subsequent to Right Hemisphere Brain Damage: A Systematic Review and Meta-Analysis. J Int Neuropsychol Soc, 1–27. 10.1017/S1355617721000825

101. Sung, J. E., Choi, S., Eom, B., Yoo, J. K., & Jeong, J. H. (2020). Syntactic complexity as a linguistic marker to differentiate mild cognitive impairment from normal aging. Journal of Speech, Language, and Hearing Research, 63(5), 1416–1429.

102. Swinburn, K., Best, W., Beeke, S., Cruice, M., Smith, L., Pearce Willis, E., Ledingham, K., Sweeney, J., & McVicker, S. J. (2019). A concise patient reported outcome measure for people with aphasia: The aphasia impact questionnaire 21. Aphasiology, 33(9), 1035–1060.

103. Themistocleous, C. (2017). The Nature of Phonetic Gradience across a Dialect Continuum: Evidence from Modern Greek Vowels [Journal Article]. Phonetica, 74(3), 157–172. 10.1159/000450554)

104. Themistocleous, C. (2019). Dialect Classification From a Single Sonorant Sound Using Deep Neural Networks. Frontiers in Communication, 4, 1–12. 10.3389/fcomm.2019.00064

105. Themistocleous, C. (2024). Open Brain AI and language assessment. Frontiers in Human Neuroscience, 18. 10.3389/fnhum.2024.1421435

106. Themistocleous, C. (2025). Linguistic and Emotional Prosody: A Systematic Review and ALE Meta-Analysis. Neuroscience & Biobehavioral Reviews, 106210. 10.1016/j.neubiorev.2025.106210

107. Themistocleous, C., Eckerström, M., & Kokkinakis, D. (2018). Identification of Mild Cognitive Impairment From Speech in Swedish Using Deep Sequential Neural Networks [10.3389/fneur.2018.00975]. *Frontiers in Neurology*, 9, 975. 10.3389/fneur.2018.00975

108. Themistocleous, C., Eckerström, M., & Kokkinakis, D. (2020). Voice quality and speech fluency distinguish individuals with Mild Cognitive Impairment from Healthy Controls. PLoS One, 15(7), e0236009. 10.1371/journal.pone.0236009

109. Themistocleous, C., Ficek, B., Webster, K., den Ouden, D.-B., Hillis, A. E., & Tsapkini, K. (2020). Automatic subtyping of individuals with Primary Progressive Aphasia. bioRxiv, 2020.2004.2004.025593. 10.1101/2020.04.04.025593

110. Themistocleous, C., Webster, K., Afthinos, A., & Tsapkini, K. (2020). Part of Speech Production in Patients With Primary Progressive Aphasia: An Analysis Based on Natural Language Processing. American Journal of Speech-Language Pathology, 1-15. 10.1044/2020_AJSLP-19-00114

111. Thompson, C. K., & Mack, J. E. (2014). Grammatical Impairments in PPA. Aphasiology, 28(8-9), 1018–1037. 10.1080/02687038.2014.912744

112. Thompson, C. K., Riley, E. A., den Ouden, D. B., Meltzer-Asscher, A., & Lukic, S. (2013). Training verb argument structure production in agrammatic aphasia: Behavioral and neural recovery patterns. Cortex, null(null), null.

113. Thompson, C. K., Shapiro, L. P., Kiran, S., & Sobecks, J. (2003). The role of syntactic complexity in treatment of sentence deficits in agrammatic aphasia: the complexity account of treatment efficacy (CATE). Journal of speech, language, and hearing research: JSLHR, 46(3), 591–607.

114. Tombaugh, T. N., & McIntyre, N. J. (1992). The Mini-Mental State Examination: A comprehensive review. Journal of the American Geriatrics Society, 40(9), 922–935. 10.1111/j.1532-5415.1992.tb01992.x

115. Tsapkini, K., Webster, K. T., Ficek, B. N., Desmond, J. E., Onyike, C. U., Rapp, B., Frangakis, C. E., & Hillis, A. E. (2018). Electrical brain stimulation in different variants of primary progressive aphasia: A randomized clinical trial. *Alzheimer’s & dementia (New York*, N. Y.), 4, 461–472. 10.1016/j.trci.2018.08.002

116. Tuomiranta, L., Laura, E., & and Laakso, M. (2025). Self-initiated self-repairs of connected speech and novel vocabulary learning during the first year of recovery from aphasia: four longitudinal case studies. Aphasiology, 39(3), 321–345. 10.1080/02687038.2024.2347386

117. Turkeltaub, P. E. (2015). Brain Stimulation and the Role of the Right Hemisphere in Aphasia Recovery. Current neurology and neuroscience reports, 15(11), 72.

118. Williams, E., Theys, C., & McAuliffe, M. (2023). Lexical-semantic properties of verbs and nouns used in conversation by people with Alzheimer’s disease. PLoS One, 18(8), e0288556. 10.1371/journal.pone.0288556

119. Wilson, S. M., DeMarco, A. T., Henry, M. L., Gesierich, B., Babiak, M., Miller, B. L., & Gorno-Tempini, M. L. (2016). Variable disruption of a syntactic processing network in primary progressive aphasia. Brain, 139(11), 2994–3006. 10.1093/brain/aww218

120. Wilson, S. M., Eriksson, D. K., Schneck, S. M., & Lucanie, J. M. (2018). A quick aphasia battery for efficient, reliable, and multidimensional assessment of language function. PLoS One, 13(2), e0192773. 10.1371/journal.pone.0192773

