## Supplementary material for "Automatic screening and characterization of patients with acquired neurological conditions from language": SD8

Acronyms, abbreviations, and terminology for neurological conditions and clinical research databases used in language and communication studies

| Acronym | Full Name | Description |
| --- | --- | --- |
| **HC** | Healthy Controls | Cognitively normal participants used as comparison groups in research studies |
| **LHD** | Language Disorder | Loss of language abilities, often arising from an ischemic stroke that blocks blood supply to an area of the brain. Most common adult communication disorder affecting ~2 million people in the US |
| **Dementia** | Cognitive Decline Disorder | Progressive neurological condition affecting memory, language, and other cognitive functions (as in your Pitt study) |
| **MCI** | Mild Cognitive Impairment | Cognitive decline greater than normal aging but not severe enough to meet dementia criteria |
| **RHD** | Right Hemisphere Damage | Brain damage affecting the right hemisphere, leading to specific cognitive-linguistic deficits |
| **TBI** | Traumatic Brain Injury | Brain injury resulting from external trauma, affecting communication and cognitive functions |
| **Pitt Study/Pitt** | Pittsburgh Corpus | The University of Pittsburgh Alzheimer's research study (part of DementiaBank) |
| **AphasiaBank** | Aphasia Research Database | The only openly available data source for spoken language and communication in aphasia. Model for other adult language databases |
| **TBI Bank** | Traumatic Brain Injury Database | Database for studying language and communication in people with traumatic brain injury, using protocols like AphasiaBank |
| **RHD Bank** | Right Hemisphere Damage Database | Database for studying language production behaviors and cognitive-linguistic deficits associated with right hemisphere brain damage |
| **DementiaBank** | Dementia Research Database | The overarching database containing your Pitt corpus data for studying language in dementia |
| **Delaware** | Delaware Corpus | A newer corpus within DementiaBank with updated protocols for dementia research |
| **DementiaBank MCI** | DementiaBank | DementiaBank protocol data for individuals with MCI, provided by the Baycrest Research Center Corpus and other centers |
