## Supplementary material for "Automatic screening and characterization of patients with acquired neurological conditions from language": SD7

**Table 1. Performance comparison of machine learning models for binary classification of patient vs. HC**

| **Model** | **HC Recall (Specificity)** | **Patient Recall (Sensitivity)** | **Balanced Accuracy** | **AUC-ROC** | **AUC-PR** |
| --- | --- | --- | --- | --- | --- |
| LR | 0.98 | 0.99 | 0.99 | 0.920 | 0.909 |
| SVM | 0.93 | 0.98 | 0.96 | 0.970 | 0.972 |
| DNN | 0.91 | 0.99 | 0.95 | 0.966 | 0.935 |
| GB | 0.88 | 0.93 | 0.90 | 0.918 | 0.925 |
| RF | 0.86 | 0.89 | 0.88 | 0.902 | 0.897 |

**Table 2. Performance comparison of machine learning models for multi-class classification of neurological conditions.**

| **Model** | **Category** | **Support** | **Precision** | **Recall** | **F1-Score** |
| --- | --- | --- | --- | --- | --- |
| SVM | LHD | 1173 | 0.94 | 0.99 | 0.96 |
|  | Dementia | 47 | 0.89 | 0.83 | 0.86 |
|  | HC | 573 | 0.94 | 0.93 | 0.94 |
|  | Other | 211 | 0.94 | 0.72 | 0.82 |
| LR | LHD | 1173 | 0.98 | 0.92 | 0.95 |
|  | Dementia | 47 | 0.83 | 0.94 | 0.88 |
|  | HC | 573 | 0.98 | 0.98 | 0.98 |
|  | Other | 211 | 0.66 | 0.87 | 0.75 |
| GB | LHD | 1173 | 0.94 | 0.92 | 0.93 |
|  | Dementia | 47 | 0.78 | 0.85 | 0.82 |
|  | HC | 573 | 0.83 | 0.88 | 0.85 |
|  | Other | 211 | 0.45 | 0.40 | 0.43 |
| DNN | LHD | 1173 | 1.00 | 0.90 | 0.94 |
|  | Dementia | 47 | 0.61 | 0.98 | 0.75 |
|  | HC | 573 | 0.97 | 0.91 | 0.94 |
|  | Other | 211 | 0.59 | 0.93 | 0.72 |
| RF | LHD | 1173 | 0.91 | 0.94 | 0.92 |
|  | Dementia | 47 | 0.80 | 0.77 | 0.78 |
|  | HC | 573 | 0.75 | 0.86 | 0.81 |
|  | Other | 211 | 0.51 | 0.23 | 0.31 |

Notes: Classification performance metrics (precision, recall, and F1-score) for five machine learning models across four categories: Left Hemisphere Damage (LHD), Dementia, Healthy Controls (HC), and Other Neurological Conditions. Support indicates the number of test samples in each category. SVM: Support Vector Machine, LR: Logistic Regression, GB: Gradient Boosting, DNN: Deep Neural Network, RF: Random Forest.
