## Supplementary material for "Automatic screening and characterization of patients with acquired neurological conditions from language": SD5

### Supplementary Table 2 Measures ranked by Effect Sizes (Partial η²).

|  | Measure | Category | F | Num DF | Den DF | p value | Partial η2 |
| --- | --- | --- | --- | --- | --- | --- | --- |
| 1.00 | Definite Indefinite Count | 87.39 | 2.00 | 431.02 | 0.00 | 0.29 | 0.26 |
| 2.00 | Cardinal Number Count | 90.88 | 2.00 | 526.94 | 0.00 | 0.26 | 0.24 |
| 3.00 | Types | 62.63 | 2.00 | 598.34 | 0.00 | 0.17 | 0.19 |
| 4.00 | CVCC | 63.29 | 2.00 | 630.04 | 0.00 | 0.17 | 0.20 |
| 5.00 | Number Plural Count | 56.81 | 2.00 | 568.22 | 0.00 | 0.17 | 0.19 |
| 6.00 | Content Words Unique | 57.43 | 2.00 | 577.53 | 0.00 | 0.17 | 0.18 |
| 7.00 | Attribute Count | 67.64 | 2.00 | 683.24 | 0.00 | 0.17 | 0.24 |
| 8.00 | 2 syllables word | 57.92 | 2.00 | 594.29 | 0.00 | 0.16 | 0.19 |
| 9.00 | Appositional modifier Count | 41.96 | 2.00 | 431.04 | 0.00 | 0.16 | 0.21 |
| 10.00 | Degree Positive Count | 48.38 | 2.00 | 499.73 | 0.00 | 0.16 | 0.17 |
| 11.00 | Adjective Count | 57.71 | 2.00 | 600.00 | 0.00 | 0.16 | 0.19 |
| 12.00 | Adjective Phrases | 57.47 | 2.00 | 597.62 | 0.00 | 0.16 | 0.19 |
| 13.00 | Adjectival modifier Count | 56.27 | 2.00 | 586.36 | 0.00 | 0.16 | 0.19 |
| 14.00 | Numeral Count | 88.21 | 2.00 | 941.26 | 0.00 | 0.16 | 0.24 |
| 15.00 | Noun Count | 52.31 | 2.00 | 570.44 | 0.00 | 0.15 | 0.17 |
| 16.00 | Expletive Count | 16.51 | 2.00 | 180.42 | 0.00 | 0.15 | 0.11 |
| 17.00 | Syllables | 53.61 | 2.00 | 591.15 | 0.00 | 0.15 | 0.18 |
| 18.00 | CVC | 53.58 | 2.00 | 593.36 | 0.00 | 0.15 | 0.17 |
| 19.00 | Content Words Total | 51.06 | 2.00 | 576.36 | 0.00 | 0.15 | 0.18 |
| 20.00 | Total Characters in Text Letters Only | 51.64 | 2.00 | 585.38 | 0.00 | 0.15 | 0.17 |
| 21.00 | Corrected TTR CTTR | 82.39 | 2.00 | 937.05 | 0.00 | 0.15 | 0.20 |
| 22.00 | Prepositional modifier Count | 51.04 | 2.00 | 581.51 | 0.00 | 0.15 | 0.18 |
| 23.00 | Prepositional Phrases | 51.79 | 2.00 | 590.31 | 0.00 | 0.15 | 0.18 |
| 24.00 | Unclassified dependent Count | 10.19 | 2.00 | 117.38 | 0.00 | 0.15 | 0.05 |
| 25.00 | Adposition Count | 51.23 | 2.00 | 593.33 | 0.00 | 0.15 | 0.18 |
| 26.00 | CV | 59.48 | 2.00 | 693.62 | 0.00 | 0.15 | 0.18 |
| 27.00 | Verb Phrases | 50.38 | 2.00 | 590.91 | 0.00 | 0.15 | 0.17 |
| 28.00 | Direct object Count | 46.34 | 2.00 | 543.58 | 0.00 | 0.15 | 0.17 |
| 29.00 | Words Tokens | 49.53 | 2.00 | 582.53 | 0.00 | 0.15 | 0.17 |
| 30.00 | Verb Count | 48.24 | 2.00 | 571.17 | 0.00 | 0.14 | 0.17 |
| 31.00 | VC | 75.21 | 2.00 | 893.24 | 0.00 | 0.14 | 0.22 |
| 32.00 | CCVCC | 68.10 | 2.00 | 812.79 | 0.00 | 0.14 | 0.23 |
| 33.00 | Function Words Total | 49.99 | 2.00 | 598.63 | 0.00 | 0.14 | 0.17 |
| 34.00 | Total Complex T units | 69.42 | 2.00 | 838.58 | 0.00 | 0.14 | 0.21 |
| 35.00 | Estimated Reading Time sec | 47.42 | 2.00 | 575.91 | 0.00 | 0.14 | 0.16 |
| 36.00 | Root Count | 48.25 | 2.00 | 586.97 | 0.00 | 0.14 | 0.17 |
| 37.00 | Total Dependent Clauses | 66.17 | 2.00 | 812.52 | 0.00 | 0.14 | 0.20 |
| 38.00 | Object of preposition Count | 46.11 | 2.00 | 568.80 | 0.00 | 0.14 | 0.17 |
| 39.00 | PronType Personal Count | 50.96 | 2.00 | 632.16 | 0.00 | 0.14 | 0.17 |
| 40.00 | Smog Index | 86.37 | 2.00 | 1,085.65 | 0.00 | 0.14 | 0.14 |
| 41.00 | Total Clauses | 44.58 | 2.00 | 560.82 | 0.00 | 0.14 | 0.14 |
| 42.00 | Total T units | 44.58 | 2.00 | 560.82 | 0.00 | 0.14 | 0.14 |
| 43.00 | Compound modifier Count | 62.32 | 2.00 | 785.44 | 0.00 | 0.14 | 0.21 |
| 44.00 | Total Complex Nominals | 45.78 | 2.00 | 577.76 | 0.00 | 0.14 | 0.15 |
| 45.00 | Gender Neuter Count | 54.46 | 2.00 | 696.14 | 0.00 | 0.14 | 0.21 |
| 46.00 | Case Nominative Count | 51.81 | 2.00 | 663.02 | 0.00 | 0.14 | 0.17 |
| 47.00 | Tense Present Count | 43.39 | 2.00 | 555.47 | 0.00 | 0.14 | 0.16 |
| 48.00 | Sentences Alphabetic Only | 44.90 | 2.00 | 575.50 | 0.00 | 0.13 | 0.16 |
| 49.00 | Determiner Count | 45.09 | 2.00 | 577.94 | 0.00 | 0.13 | 0.15 |
| 50.00 | Total Sentences | 43.97 | 2.00 | 574.15 | 0.00 | 0.13 | 0.16 |
| 51.00 | Clausal complement Count | 46.80 | 2.00 | 615.52 | 0.00 | 0.13 | 0.19 |
| 52.00 | Total Characters in Text All symbols | 43.56 | 2.00 | 573.79 | 0.00 | 0.13 | 0.15 |
| 53.00 | Particle Count | 54.87 | 2.00 | 724.84 | 0.00 | 0.13 | 0.19 |
| 54.00 | Total Words | 43.19 | 2.00 | 574.50 | 0.00 | 0.13 | 0.15 |
| 55.00 | Total Tree Height | 42.67 | 2.00 | 572.59 | 0.00 | 0.13 | 0.15 |
| 56.00 | Number Singular Count | 42.65 | 2.00 | 573.16 | 0.00 | 0.13 | 0.15 |
| 57.00 | Noun Phrases | 42.55 | 2.00 | 573.66 | 0.00 | 0.13 | 0.15 |
| 58.00 | Total Classical Yngve Load | 42.47 | 2.00 | 575.78 | 0.00 | 0.13 | 0.15 |
| 59.00 | Difficult Words | 64.47 | 2.00 | 875.73 | 0.00 | 0.13 | 0.20 |
| 60.00 | Auxiliary passive Count | 25.22 | 2.00 | 345.66 | 0.00 | 0.13 | 0.13 |
| 61.00 | PronType Article Count | 41.83 | 2.00 | 578.03 | 0.00 | 0.13 | 0.14 |
| 62.00 | CCVC | 52.40 | 2.00 | 726.34 | 0.00 | 0.13 | 0.18 |
| 63.00 | Adverbial Phrases | 49.40 | 2.00 | 687.28 | 0.00 | 0.13 | 0.17 |
| 64.00 | Adverb Count | 49.51 | 2.00 | 688.87 | 0.00 | 0.13 | 0.17 |
| 65.00 | VerbForm Finite Count | 41.19 | 2.00 | 573.45 | 0.00 | 0.13 | 0.14 |
| 66.00 | Pronoun Count | 42.40 | 2.00 | 593.60 | 0.00 | 0.12 | 0.15 |
| 67.00 | Auxiliary Count | 45.51 | 2.00 | 637.20 | 0.00 | 0.12 | 0.16 |
| 68.00 | Person Third Count | 40.62 | 2.00 | 569.61 | 0.00 | 0.12 | 0.14 |
| 69.00 | Function Words Unique | 59.34 | 2.00 | 842.94 | 0.00 | 0.12 | 0.17 |
| 70.00 | Mood Indicative Count | 40.07 | 2.00 | 577.70 | 0.00 | 0.12 | 0.15 |
| 71.00 | Adverbial clause modifier Count | 48.12 | 2.00 | 698.75 | 0.00 | 0.12 | 0.19 |
| 72.00 | Aspect Progressive Count | 48.66 | 2.00 | 707.00 | 0.00 | 0.12 | 0.18 |
| 73.00 | Nominal subject Count | 39.17 | 2.00 | 570.06 | 0.00 | 0.12 | 0.14 |
| 74.00 | VerbForm Participle Count | 51.65 | 2.00 | 760.45 | 0.00 | 0.12 | 0.19 |
| 75.00 | Gender Feminine Count | 41.57 | 2.00 | 616.11 | 0.00 | 0.12 | 0.14 |
| 76.00 | Nominal subject passive Count | 18.58 | 2.00 | 278.11 | 0.00 | 0.12 | 0.12 |
| 77.00 | Adverbial modifier Count | 57.71 | 2.00 | 871.05 | 0.00 | 0.12 | 0.19 |
| 78.00 | Adjectival complement Count | 34.18 | 2.00 | 531.31 | 0.00 | 0.11 | 0.15 |
| 79.00 | PronType Indefinite Count | 18.49 | 2.00 | 293.57 | 0.00 | 0.11 | 0.13 |
| 80.00 | NumType Cardinal Count | 30.90 | 2.00 | 493.59 | 0.00 | 0.11 | 0.13 |
| 81.00 | Possession modifier Count | 44.91 | 2.00 | 732.30 | 0.00 | 0.11 | 0.16 |
| 82.00 | V | 61.39 | 2.00 | 1,001.77 | 0.00 | 0.11 | 0.18 |
| 83.00 | Definite Definite Count | 34.31 | 2.00 | 562.37 | 0.00 | 0.11 | 0.12 |
| 84.00 | CCV | 34.86 | 2.00 | 580.36 | 0.00 | 0.11 | 0.11 |
| 85.00 | Coordinating conjunction Count | 34.42 | 2.00 | 584.10 | 0.00 | 0.11 | 0.12 |
| 86.00 | Relative clause modifier Count | 33.10 | 2.00 | 568.12 | 0.00 | 0.10 | 0.14 |
| 87.00 | ConjType Cmp Count | 32.35 | 2.00 | 559.94 | 0.00 | 0.10 | 0.11 |
| 88.00 | CVCCC | 43.22 | 2.00 | 753.69 | 0.00 | 0.10 | 0.17 |
| 89.00 | Clauses Total Clauses in Text | 33.01 | 2.00 | 580.92 | 0.00 | 0.10 | 0.13 |
| 90.00 | Case Accusative Count | 44.13 | 2.00 | 777.07 | 0.00 | 0.10 | 0.18 |
| 91.00 | PronType Relative Count | 8.13 | 2.00 | 144.32 | 0.00 | 0.10 | 0.06 |
| 92.00 | VerbForm Infinitive Count | 49.95 | 2.00 | 888.80 | 0.00 | 0.10 | 0.17 |
| 93.00 | Polarity Negative Count | 29.05 | 2.00 | 526.17 | 0.00 | 0.10 | 0.13 |
| 94.00 | Poss Possessive Count | 35.54 | 2.00 | 651.61 | 0.00 | 0.10 | 0.13 |
| 95.00 | Negation modifier Count | 28.90 | 2.00 | 536.15 | 0.00 | 0.10 | 0.13 |
| 96.00 | Open clausal complement Count | 36.00 | 2.00 | 673.58 | 0.00 | 0.10 | 0.15 |
| 97.00 | Tense Past Count | 46.29 | 2.00 | 880.03 | 0.00 | 0.10 | 0.16 |
| 98.00 | VerbType Mod Count | 3.09 | 2.00 | 60.34 | 0.05 | 0.09 | 0.04 |
| 99.00 | Aspect Perfective Count | 24.71 | 2.00 | 494.02 | 0.00 | 0.09 | 0.13 |
| 100.00 | VCC | 28.06 | 2.00 | 571.22 | 0.00 | 0.09 | 0.10 |
| 101.00 | Number modifier Count | 18.56 | 2.00 | 381.56 | 0.00 | 0.09 | 0.10 |
| 102.00 | X3 syllables word | 37.95 | 2.00 | 796.55 | 0.00 | 0.09 | 0.14 |
| 103.00 | Organization Ratio | 32.79 | 2.00 | 699.95 | 0.00 | 0.09 | 0.12 |
| 104.00 | Marker Count | 25.82 | 2.00 | 552.70 | 0.00 | 0.09 | 0.11 |
| 105.00 | Proper noun Count | 28.04 | 2.00 | 608.32 | 0.00 | 0.08 | 0.09 |
| 106.00 | Cardinal Number Ratio | 44.47 | 2.00 | 1,002.06 | 0.00 | 0.08 | 0.10 |
| 107.00 | Total Coordinate Phrases | 27.57 | 2.00 | 645.82 | 0.00 | 0.08 | 0.09 |
| 108.00 | Subordinating conjunction Count | 40.98 | 2.00 | 973.14 | 0.00 | 0.08 | 0.14 |
| 109.00 | Person Second Count | 8.67 | 2.00 | 207.57 | 0.00 | 0.08 | 0.06 |
| 110.00 | Case marker Count | 12.74 | 2.00 | 312.45 | 0.00 | 0.08 | 0.11 |
| 111.00 | PronType Demonstrative Ratio | 28.21 | 2.00 | 695.31 | 0.00 | 0.08 | 0.08 |
| 112.00 | Conjunct Count | 23.30 | 2.00 | 595.57 | 0.00 | 0.07 | 0.09 |
| 113.00 | PronType Relative Ratio | 10.40 | 2.00 | 293.84 | 0.00 | 0.07 | 0.12 |
| 114.00 | Max Sentence Length | 23.50 | 2.00 | 665.00 | 0.00 | 0.07 | 0.07 |
| 115.00 | VerbType Mod Ratio | 6.77 | 2.00 | 194.45 | 0.00 | 0.07 | 0.06 |
| 116.00 | Degree Cmp Count | 4.89 | 2.00 | 142.23 | 0.01 | 0.06 | 0.07 |
| 117.00 | Unclassified dependent Ratio | 24.81 | 2.00 | 746.48 | 0.00 | 0.06 | 0.11 |
| 118.00 | Case marker Ratio | 20.73 | 2.00 | 623.82 | 0.00 | 0.06 | 0.06 |
| 119.00 | CCVCCC | 4.49 | 2.00 | 138.59 | 0.01 | 0.06 | 0.04 |
| 120.00 | PronType Demonstrative Count | 26.26 | 2.00 | 816.11 | 0.00 | 0.06 | 0.13 |
| 121.00 | 4 syllables word | 17.87 | 2.00 | 594.80 | 0.00 | 0.06 | 0.06 |
| 122.00 | Proper noun Ratio | 20.92 | 2.00 | 761.62 | 0.00 | 0.05 | 0.09 |
| 123.00 | Person Count | 29.45 | 2.00 | 1,107.79 | 0.00 | 0.05 | 0.08 |
| 124.00 | Gunning Fog Index | 27.21 | 2.00 | 1,036.36 | 0.00 | 0.05 | 0.05 |
| 125.00 | CCCVC | 4.99 | 2.00 | 193.95 | 0.01 | 0.05 | 0.06 |
| 126.00 | Complement of preposition Count | 4.60 | 2.00 | 178.96 | 0.01 | 0.05 | 0.05 |
| 127.00 | Degree Cmp Ratio | 5.28 | 2.00 | 209.46 | 0.01 | 0.05 | 0.05 |
| 128.00 | Other Ratio | 3.62 | 2.00 | 154.89 | 0.03 | 0.04 | 0.13 |
| 129.00 | Aspect Perfective Ratio | 11.15 | 2.00 | 507.11 | 0.00 | 0.04 | 0.06 |
| 130.00 | Gender Masculine Count | 14.01 | 2.00 | 665.77 | 0.00 | 0.04 | 0.07 |
| 131.00 | Other Count | 7.06 | 2.00 | 336.10 | 0.00 | 0.04 | 0.00 |
| 132.00 | Type Token Ratio TTR | 17.86 | 2.00 | 955.88 | 0.00 | 0.04 | 0.05 |
| 133.00 | Linsear Write Formula | 16.89 | 2.00 | 931.51 | 0.00 | 0.03 | 0.08 |
| 134.00 | Person First Count | 14.60 | 2.00 | 841.25 | 0.00 | 0.03 | 0.05 |
| 135.00 | Clausal modifier of noun Count | 2.50 | 2.00 | 147.81 | 0.09 | 0.03 | 0.03 |
| 136.00 | Person Second Ratio | 6.39 | 2.00 | 421.71 | 0.00 | 0.03 | 0.10 |
| 137.00 | Mean Sentence Length in Words | 12.16 | 2.00 | 804.54 | 0.00 | 0.03 | 0.03 |
| 138.00 | Clausal complement Ratio | 5.86 | 2.00 | 394.42 | 0.00 | 0.03 | 0.03 |
| 139.00 | Dale Chall Readability Score | 14.23 | 2.00 | 981.68 | 0.00 | 0.03 | 0.07 |
| 140.00 | Aspect Progressive Ratio | 11.45 | 2.00 | 790.74 | 0.00 | 0.03 | 0.05 |
| 141.00 | Average Tree Height | 12.96 | 2.00 | 901.18 | 0.00 | 0.03 | 0.03 |
| 142.00 | Gender Masculine Ratio | 9.74 | 2.00 | 696.56 | 0.00 | 0.03 | 0.05 |
| 143.00 | PronType Indefinite Ratio | 4.28 | 2.00 | 308.78 | 0.01 | 0.03 | 0.04 |
| 144.00 | 5 syllables word | 0.43 | 2.00 | 31.61 | 0.65 | 0.03 | 0.00 |
| 145.00 | Automated Readability Index | 10.23 | 2.00 | 759.04 | 0.00 | 0.03 | 0.03 |
| 146.00 | Coleman Liau Index | 12.27 | 2.00 | 911.88 | 0.00 | 0.03 | 0.04 |
| 147.00 | Marker Ratio | 4.23 | 2.00 | 340.76 | 0.02 | 0.02 | 0.02 |
| 148.00 | Appositional modifier Ratio | 1.64 | 2.00 | 132.22 | 0.20 | 0.02 | 0.01 |
| 149.00 | Numeral Ratio | 11.22 | 2.00 | 906.21 | 0.00 | 0.02 | 0.04 |
| 150.00 | VerbForm Participle Ratio | 10.68 | 2.00 | 862.71 | 0.00 | 0.02 | 0.03 |
| 151.00 | Average Sentence Length | 9.26 | 2.00 | 790.92 | 0.00 | 0.02 | 0.02 |
| 152.00 | Object of preposition Ratio | 11.71 | 2.00 | 1,022.61 | 0.00 | 0.02 | 0.05 |
| 153.00 | Adverbial clause modifier Ratio | 5.91 | 2.00 | 547.60 | 0.00 | 0.02 | 0.04 |
| 154.00 | Root Ratio | 10.59 | 2.00 | 981.61 | 0.00 | 0.02 | 0.02 |
| 155.00 | Max Classical Yngve Load | 5.58 | 2.00 | 556.02 | 0.00 | 0.02 | 0.02 |
| 156.00 | Dative Count | 0.53 | 2.00 | 54.17 | 0.59 | 0.02 | 0.01 |
| 157.00 | Interjection Count | 10.73 | 2.00 | 1,106.90 | 0.00 | 0.02 | 0.04 |
| 158.00 | Auxiliary Ratio | 9.12 | 2.00 | 942.66 | 0.00 | 0.02 | 0.04 |
| 159.00 | Min Sentence Length | 3.51 | 2.00 | 366.69 | 0.03 | 0.02 | 0.02 |
| 160.00 | Nominal subject passive Ratio | 2.69 | 2.00 | 282.95 | 0.07 | 0.02 | 0.01 |
| 161.00 | Flesch Kincaid Grade Level | 8.33 | 2.00 | 953.12 | 0.00 | 0.02 | 0.02 |
| 162.00 | Nominal subject Ratio | 8.95 | 2.00 | 1,045.01 | 0.00 | 0.02 | 0.04 |
| 163.00 | Max Left Branching Depth | 6.98 | 2.00 | 845.08 | 0.00 | 0.02 | 0.02 |
| 164.00 | Definite Indefinite Ratio | 5.83 | 2.00 | 754.52 | 0.00 | 0.02 | 0.03 |
| 165.00 | Tense Present Ratio | 8.17 | 2.00 | 1,065.86 | 0.00 | 0.02 | 0.03 |
| 166.00 | Organization Count | 4.61 | 2.00 | 613.65 | 0.01 | 0.01 | 0.02 |
| 167.00 | Number modifier Ratio | 7.04 | 2.00 | 981.34 | 0.00 | 0.01 | 0.02 |
| 168.00 | Average Word Length in Characters | 5.02 | 2.00 | 701.95 | 0.01 | 0.01 | 0.03 |
| 169.00 | Definite Definite Ratio | 6.82 | 2.00 | 979.33 | 0.00 | 0.01 | 0.03 |
| 170.00 | CCCV | 0.89 | 2.00 | 132.36 | 0.41 | 0.01 | 0.02 |
| 171.00 | Compound modifier Ratio | 3.33 | 2.00 | 545.32 | 0.04 | 0.01 | 0.01 |
| 172.00 | Mean Classical Yngve Load | 5.10 | 2.00 | 857.25 | 0.01 | 0.01 | 0.02 |
| 173.00 | Determiner Ratio | 5.90 | 2.00 | 1,030.49 | 0.00 | 0.01 | 0.02 |
| 174.00 | Gender Neuter Ratio | 4.89 | 2.00 | 889.15 | 0.01 | 0.01 | 0.02 |
| 175.00 | Gender Feminine Ratio | 3.38 | 2.00 | 621.08 | 0.03 | 0.01 | 0.02 |
| 176.00 | Conjunct Ratio | 3.31 | 2.00 | 643.52 | 0.04 | 0.01 | 0.01 |
| 177.00 | Tense Past Ratio | 4.52 | 2.00 | 895.95 | 0.01 | 0.01 | 0.02 |
| 178.00 | Poss Possessive Ratio | 1.19 | 2.00 | 239.03 | 0.30 | 0.01 | 0.00 |
| 179.00 | Subordinating conjunction Ratio | 2.81 | 2.00 | 563.42 | 0.06 | 0.01 | 0.01 |
| 180.00 | Open clausal complement Ratio | 4.93 | 2.00 | 992.74 | 0.01 | 0.01 | 0.01 |
| 181.00 | Auxiliary passive Ratio | 2.15 | 2.00 | 443.68 | 0.12 | 0.01 | 0.01 |
| 182.00 | Adjective Ratio | 5.17 | 2.00 | 1,092.87 | 0.01 | 0.01 | 0.02 |
| 183.00 | Case Accusative Ratio | 3.80 | 2.00 | 805.38 | 0.02 | 0.01 | 0.02 |
| 184.00 | Mean Left Branching Depth | 4.11 | 2.00 | 893.19 | 0.02 | 0.01 | 0.02 |
| 185.00 | Relative clause modifier Ratio | 3.21 | 2.00 | 704.37 | 0.04 | 0.01 | 0.01 |
| 186.00 | Adjectival complement Ratio | 4.82 | 2.00 | 1,059.42 | 0.01 | 0.01 | 0.03 |
| 187.00 | Coordinating conjunction Ratio | 3.91 | 2.00 | 959.78 | 0.02 | 0.01 | 0.01 |
| 188.00 | Mood Indicative Ratio | 4.07 | 2.00 | 1,020.10 | 0.02 | 0.01 | 0.03 |
| 189.00 | Interjection Ratio | 3.41 | 2.00 | 896.34 | 0.03 | 0.01 | 0.02 |
| 190.00 | Particle Ratio | 3.57 | 2.00 | 946.73 | 0.03 | 0.01 | 0.01 |
| 191.00 | Prepositional modifier Ratio | 3.37 | 2.00 | 936.82 | 0.03 | 0.01 | 0.01 |
| 192.00 | Number Plural Ratio | 3.06 | 2.00 | 885.65 | 0.05 | 0.01 | 0.02 |
| 193.00 | Date Count | 3.90 | 2.00 | 1,134.42 | 0.02 | 0.01 | 0.02 |
| 194.00 | Date Ratio | 3.54 | 2.00 | 1,028.39 | 0.03 | 0.01 | 0.02 |
| 195.00 | Maas s TTR A2 | 2.52 | 2.00 | 889.85 | 0.08 | 0.01 | 0.01 |
| 196.00 | VerbForm Infinitive Ratio | 2.80 | 2.00 | 1,039.76 | 0.06 | 0.01 | 0.01 |
| 197.00 | Number Singular Ratio | 2.23 | 2.00 | 840.21 | 0.11 | 0.01 | 0.01 |
| 198.00 | Adposition Ratio | 2.58 | 2.00 | 983.65 | 0.08 | 0.01 | 0.01 |
| 199.00 | Flesch Reading Ease | 2.45 | 2.00 | 968.91 | 0.09 | 0.01 | 0.01 |
| 200.00 | Automated Lexical Density Content Words Total Words | 2.36 | 2.00 | 944.57 | 0.09 | 0.00 | 0.01 |
| 201.00 | Noun Ratio | 2.51 | 2.00 | 1,009.59 | 0.08 | 0.00 | 0.01 |
| 202.00 | PronType Personal Ratio | 2.45 | 2.00 | 1,050.44 | 0.09 | 0.00 | 0.01 |
| 203.00 | Polarity Negative Ratio | 1.62 | 2.00 | 713.64 | 0.20 | 0.00 | 0.01 |
| 204.00 | Person Ratio | 2.37 | 2.00 | 1,073.08 | 0.09 | 0.00 | 0.00 |
| 205.00 | Complement of preposition Ratio | 0.84 | 2.00 | 398.26 | 0.43 | 0.00 | 0.00 |
| 206.00 | Clausal modifier of noun Ratio | 0.64 | 2.00 | 309.61 | 0.53 | 0.00 | 0.00 |
| 207.00 | VerbForm Finite Ratio | 1.88 | 2.00 | 958.52 | 0.15 | 0.00 | 0.00 |
| 208.00 | Adjectival modifier Ratio | 1.24 | 2.00 | 737.12 | 0.29 | 0.00 | 0.00 |
| 209.00 | Passive Sentences Percent | 0.87 | 2.00 | 530.80 | 0.42 | 0.00 | 0.00 |
| 210.00 | Person First Ratio | 1.21 | 2.00 | 839.37 | 0.30 | 0.00 | 0.00 |
| 211.00 | Verb Ratio | 1.46 | 2.00 | 1,084.36 | 0.23 | 0.00 | 0.00 |
| 212.00 | Degree Positive Ratio | 1.30 | 2.00 | 966.65 | 0.27 | 0.00 | 0.01 |
| 213.00 | Pronoun Ratio | 1.36 | 2.00 | 1,014.27 | 0.26 | 0.00 | 0.01 |
| 214.00 | ConjType Cmp Ratio | 1.09 | 2.00 | 922.87 | 0.34 | 0.00 | 0.00 |
| 215.00 | Dative Ratio | 0.76 | 2.00 | 652.30 | 0.47 | 0.00 | 0.00 |
| 216.00 | Expletive Ratio | 0.83 | 2.00 | 721.90 | 0.44 | 0.00 | 0.00 |
| 217.00 | Adverb Ratio | 1.14 | 2.00 | 1,034.58 | 0.32 | 0.00 | 0.01 |
| 218.00 | Possession modifier Ratio | 0.77 | 2.00 | 744.18 | 0.46 | 0.00 | 0.00 |
| 219.00 | Direct object Ratio | 0.98 | 2.00 | 1,100.41 | 0.38 | 0.00 | 0.00 |
| 220.00 | Case Nominative Ratio | 0.88 | 2.00 | 1,025.57 | 0.42 | 0.00 | 0.00 |
| 221.00 | Attribute Ratio | 0.53 | 2.00 | 643.63 | 0.59 | 0.00 | 0.00 |
| 222.00 | PronType Article Ratio | 0.78 | 2.00 | 1,010.58 | 0.46 | 0.00 | 0.00 |
| 223.00 | NumType Cardinal Ratio | 0.27 | 2.00 | 553.15 | 0.76 | 0.00 | 0.00 |
| 224.00 | Adverbial modifier Ratio | 0.36 | 2.00 | 1,025.42 | 0.69 | 0.00 | 0.00 |
| 225.00 | Person Third Ratio | 0.04 | 2.00 | 1,061.46 | 0.96 | 0.00 | 0.00 |
| 226.00 | Negation modifier Ratio | 0.03 | 2.00 | 1,058.31 | 0.97 | 0.00 | 0.00 |
