## Supplementary material for "Automatic screening and characterization of patients with acquired neurological conditions from language": SD4

Best Tuned Models

Five machine learning models that have been tuned for optimal performance, in this study. The final configuration of the best models after the hyperparameter tuning is shown in (1).

(1)

{'Random Forest': Pipeline(steps=[('smote', DynamicSMOTE(k_neighbors=9, random_state=30)),

('classifier',

RandomForestClassifier(max_depth=30, min_samples_split=5,

n_estimators=300, random_state=30))]),

'SVM': Pipeline(steps=[('smote', DynamicSMOTE(k_neighbors=9, random_state=30)),

('classifier', SVC(C=100, probability=True, random_state=30))]),

'Logistic Regression': Pipeline(steps=[('smote', DynamicSMOTE(k_neighbors=7, random_state=30)),

('classifier',

LogisticRegression(C=10, max_iter=1000, penalty='l1',

random_state=30, solver='liblinear'))]),

'Gradient Boosting': Pipeline(steps=[('smote', DynamicSMOTE(random_state=30)),

('classifier',

GradientBoostingClassifier(max_depth=5, random_state=30))]),

'DNN': Pipeline(steps=[('smote', DynamicSMOTE(random_state=30)),

('classifier',

KerasClassifier(batch_size=32, epochs=300, loss='sparse_categorical_crossentropy', model=<function create_dnn_model at 0x308b8dda0>, model__dropout_rate=0.3, model__kernel_regularizer_l2=0.01, model__layers=4, model__neurons=256, optimizer=<class 'keras.src.optimizers.adam.Adam'>, optimizer__learning_rate=0.005, verbose=0))])}

Best DNN params: {'smote__k_neighbors': 5, 'classifier__optimizer__learning_rate': 0.005, 'classifier__model__neurons': 256, 'classifier__model__layers': 4, 'classifier__model__kernel_regularizer_l2': 0.01, 'classifier__model__dropout_rate': 0.3, 'classifier__epochs': 300, 'classifier__batch_size': 32}

DNN stopped at epoch: 300

--- Tuned Model Evaluation on Hold-Out Test Set ---

Each model is structured as a two-step pipeline. Namely, the first step addresses class imbalance in the data using a technique called DynamicSMOTE, and the second step is the classification algorithm itself. A fixed random_state of 30 is used across the models to ensure that the results can be reproduced.

#### Random Forest

This model first balances the data using DynamicSMOTE, considering 9 nearest neighbors to generate synthetic samples. The classifier is a RandomForestClassifier consisting of 300 trees. Each tree in the forest is limited to a maximum depth of 30 levels, and a node is split only if it contains at least 5 samples.

#### Support Vector Machine (SVM)

Similar to the Random Forest, the SVM pipeline also begins by applying DynamicSMOTE with 9 nearest neighbors. The classifier is a Support Vector Classifier (SVC) with the regularization parameter, C, set to 100. A higher C value indicates a stricter penalty for misclassifying training examples. The probability=True setting enables the model to predict class probabilities.

#### Logistic Regression

This model's pipeline starts with DynamicSMOTE using 7 nearest neighbors. The classifier is a LogisticRegression model that uses L1 regularization (penalty='l1') with a regularization strength, C, of 10. L1 regularization can also be used for feature selection by driving the coefficients of less important features to zero. The liblinear solver is employed, which is well-suited for L1 regularization, and the model is set to run for a maximum of 1000 iterations to ensure convergence.

#### Gradient Boosting

The Gradient Boosting pipeline also utilizes DynamicSMOTE, with its default parameters, to balance the data. The classifier is a GradientBoostingClassifier where the individual decision trees are constrained to a maximum depth of 5 levels.

#### Deep Neural Network (DNN)

The Deep Neural Network, which also follows the same pipeline structure.

The process begins with the application of DynamicSMOTE using its default settings to handle imbalanced data. The classifier is a KerasClassifier, which wraps a Keras deep learning model for use in a scikit-learn pipeline. The model was trained for 300 epochs, processing the data in batches of 32 samples. The sparse_categorical_crossentropy loss function was used, which is appropriate for multi-class classification problems where the labels are provided as integers. A more detailed set of optimal parameters for the DNN model was also provided:

i. SMOTE: The k_neighbors for the initial DynamicSMOTE step was set to 5.
ii. Optimizer: The Adam optimizer was used with a learning rate of 0.005.
iii. Architecture:
     a. The network consists of 4 hidden layers.
     b. Each hidden layer has 256 neurons.
     c. L2 regularization with a factor of 0.01 was applied to the kernel weights to prevent overfitting by penalizing large weights.
     d. A dropout rate of 0.3 was implemented, meaning that 30% of the neurons were randomly deactivated during each training update to further combat overfitting.
iv. Training Termination: The training process concluded after completing all 300 epochs.

### Machine Learning Performance

Random Forest shows relatively lower performance, suggesting reduced effectiveness in handling class imbalance. The robust performance across multiple metrics validates the feasibility of automated neurological assessment using quantitative linguistic features extracted from language production tasks.

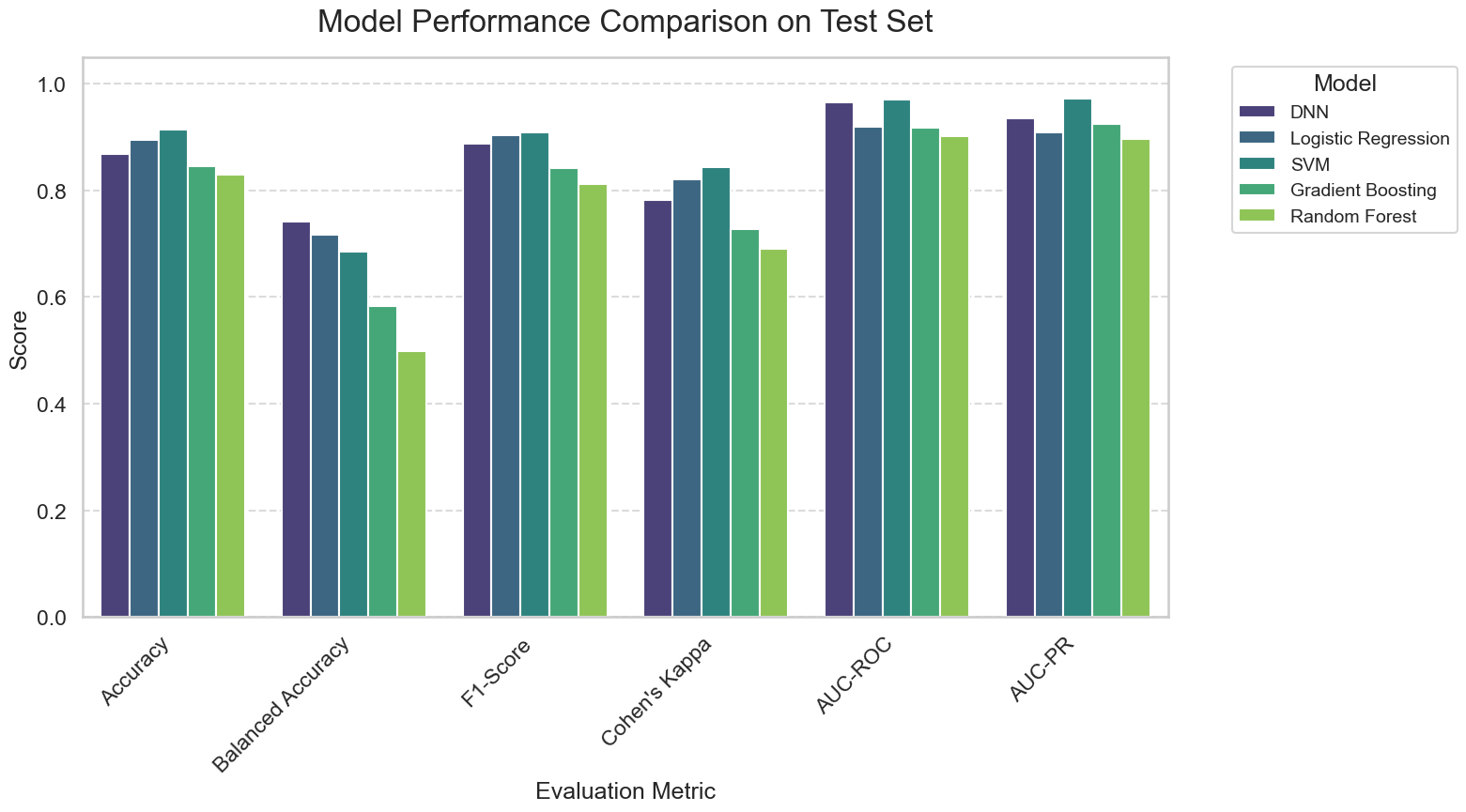

Figure 1 Comparative performance of machine learning models for neurological diagnosis classification. Performance metrics are evaluated on the independent test set across six evaluation criteria. Five machine learning algorithms are compared: Deep Neural Network (DNN, purple), Logistic Regression (dark teal), Support Vector Machine (SVM, light teal), Gradient Boosting (green), and Random Forest (light green). Evaluation metrics include Accuracy (overall classification performance), Balanced Accuracy (accuracy adjusted for class imbalance), F1-Score (harmonic mean of precision and recall), Cohen's Kappa (inter-rater agreement corrected for chance), AUC-ROC (Area Under the Receiver Operating Characteristic Curve), and AUC-PR (Area Under the Precision-Recall Curve).

Table 1 Model Performance

| Model | Accuracy | Balanced Accuracy | F1-Score | Cohen's Kappa | AUC-ROC | AUC-PR |
| --- | --- | --- | --- | --- | --- | --- |
| DNN | 0.868 | 0.741 | 0.888 | 0.783 | 0.966 | 0.935 |
| Logistic Regression | 0.895 | 0.717 | 0.905 | 0.822 | 0.920 | 0.909 |
| SVM | 0.914 | 0.685 | 0.908 | 0.844 | 0.970 | 0.972 |
| Gradient Boosting | 0.845 | 0.584 | 0.842 | 0.729 | 0.918 | 0.925 |
| Random Forest | 0.830 | 0.500 | 0.812 | 0.691 | 0.902 | 0.897 |

### Tuned Model Performance Results

#### Random Forest Model Performance

Table 2 Classification Report for Random Forest Model

| Class | Precision | Recall | F1-score | Support |
| --- | --- | --- | --- | --- |
| LHD | 0.91 | 0.94 | 0.92 | 1173 |
| Dementia | 0.8 | 0.77 | 0.78 | 47 |
| HC | 0.75 | 0.86 | 0.81 | 573 |
| MCI | 0.48 | 0.19 | 0.27 | 64 |
| RHD | 0.22 | 0.06 | 0.09 | 36 |
| TBI | 0.34 | 0.19 | 0.24 | 111 |
| accuracy |  |  | 0.83 | 2004 |
| macro avg | 0.58 | 0.5 | 0.52 | 2004 |
| weighted avg | 0.8 | 0.83 | 0.81 | 2004 |

Table 3 Confusion Matrix for Random Forest Model

| LHD | 1097 | 7 | 49 | 4 | 0 | 16 |
| --- | --- | --- | --- | --- | --- | --- |
| Dementia | 9 | 36 | 2 | 0 | 0 | 0 |
| HC | 49 | 2 | 495 | 8 | 2 | 17 |
| MCI | 12 | 0 | 34 | 12 | 3 | 3 |
| RHD | 8 | 0 | 21 | 1 | 2 | 4 |
| TBI | 33 | 0 | 55 | 0 | 2 | 21 |
|  | LHD | Dementia | HC | MCI | RHD | TBI |

#### SVM Model Performance

Table 4 Classification Report for SVM Model

| Class | Precision | Recall | F1-score | Support |
| --- | --- | --- | --- | --- |
| LHD | 0.94 | 0.99 | 0.96 | 1173 |
| Dementia | 0.89 | 0.83 | 0.86 | 47 |
| HC | 0.94 | 0.93 | 0.94 | 573 |
| MCI | 0.63 | 0.56 | 0.6 | 64 |
| RHD | 0.42 | 0.31 | 0.35 | 36 |
| TBI | 0.71 | 0.5 | 0.58 | 111 |
| accuracy |  |  | 0.91 | 2004 |
| macro avg | 0.75 | 0.68 | 0.71 | 2004 |
| weighted avg | 0.91 | 0.91 | 0.91 | 2004 |

Table 5 Confusion Matrix for SVM Model

| LHD | 1159 | 5 | 4 | 4 | 0 | 1 |
| --- | --- | --- | --- | --- | --- | --- |
| Dementia | 8 | 39 | 0 | 0 | 0 | 0 |
| HC | 38 | 0 | 531 | 1 | 0 | 3 |
| MCI | 6 | 0 | 3 | 36 | 8 | 11 |
| RHD | 4 | 0 | 7 | 6 | 11 | 8 |
| TBI | 22 | 0 | 17 | 10 | 7 | 55 |
|  | LHD | Dementia | HC | MCI | RHD | TBI |

#### Logistic Regression Model Performance

Table 6 Classification Report for Logistic Regression Model

| Class | Precision | Recall | F1-score | Support |
| --- | --- | --- | --- | --- |
| LHD | 0.98 | 0.92 | 0.95 | 1173 |
| Dementia | 0.83 | 0.94 | 0.88 | 47 |
| HC | 0.98 | 0.98 | 0.98 | 573 |
| MCI | 0.35 | 0.44 | 0.39 | 64 |
| RHD | 0.22 | 0.44 | 0.29 | 36 |
| TBI | 0.53 | 0.59 | 0.56 | 111 |
| accuracy |  |  | 0.89 | 2004 |
| macro avg | 0.65 | 0.72 | 0.67 | 2004 |
| weighted avg | 0.92 | 0.89 | 0.9 | 2004 |

Table 7 Confusion Matrix for Logistic Regression Model

| LHD | 1077 | 8 | 2 | 29 | 20 | 37 |
| --- | --- | --- | --- | --- | --- | --- |
| Dementia | 3 | 44 | 0 | 0 | 0 | 0 |
| HC | 1 | 1 | 563 | 1 | 3 | 4 |
| MCI | 5 | 0 | 0 | 28 | 21 | 10 |
| RHD | 2 | 0 | 0 | 11 | 16 | 7 |
| TBI | 13 | 0 | 7 | 12 | 14 | 65 |
|  | LHD | Dementia | HC | MCI | RHD | TBI |

#### Gradient Boosting Model Performance

Table 8 Classification Report for Gradient Boosting Model

| Class | Precision | Recall | F1-score | Support |
| --- | --- | --- | --- | --- |
| LHD | 0.94 | 0.92 | 0.93 | 1173 |
| Dementia | 0.78 | 0.85 | 0.82 | 47 |
| HC | 0.83 | 0.88 | 0.85 | 573 |
| MCI | 0.34 | 0.31 | 0.33 | 64 |
| RHD | 0.28 | 0.22 | 0.25 | 36 |
| TBI | 0.34 | 0.31 | 0.32 | 111 |
| accuracy |  |  | 0.84 | 2004 |
| macro avg | 0.58 | 0.58 | 0.58 | 2004 |
| weighted avg | 0.84 | 0.84 | 0.84 | 2004 |

Table 9 Confusion Matrix for Gradient Boosting Model

| LHD | 1084 | 8 | 20 | 20 | 5 | 36 |
| --- | --- | --- | --- | --- | --- | --- |
| Dementia | 7 | 40 | 0 | 0 | 0 | 0 |
| HC | 23 | 1 | 507 | 16 | 5 | 21 |
| MCI | 6 | 2 | 23 | 20 | 7 | 6 |
| RHD | 5 | 0 | 18 | 2 | 8 | 3 |
| TBI | 26 | 0 | 46 | 1 | 4 | 34 |
|  | LHD | Dementia | HC | MCI | RHD | TBI |

#### DNN Model Performance

Table 10 Classification Report for DNN Model

| Class | Precision | Recall | F1-score | Support |
| --- | --- | --- | --- | --- |
| LHD | 1.0 | 0.9 | 0.94 | 1173 |
| Dementia | 0.61 | 0.98 | 0.75 | 47 |
| HC | 0.97 | 0.91 | 0.94 | 573 |
| MCI | 0.4 | 0.52 | 0.45 | 64 |
| RHD | 0.17 | 0.56 | 0.26 | 36 |
| TBI | 0.49 | 0.59 | 0.53 | 111 |
| accuracy |  |  | 0.87 | 2004 |
| macro avg | 0.61 | 0.74 | 0.65 | 2004 |
| weighted avg | 0.92 | 0.87 | 0.89 | 2004 |

Table 11 Confusion Matrix for DNN Model

| LHD | 1053 | 30 | 6 | 29 | 37 | 18 |
| --- | --- | --- | --- | --- | --- | --- |
| Dementia | 1 | 46 | 0 | 0 | 0 | 0 |
| HC | 0 | 0 | 523 | 4 | 12 | 34 |
| MCI | 0 | 0 | 1 | 33 | 22 | 8 |
| RHD | 0 | 0 | 1 | 8 | 20 | 7 |
| TBI | 4 | 0 | 9 | 8 | 25 | 65 |
|  | LHD | Dementia | HC | MCI | RHD | TBI |

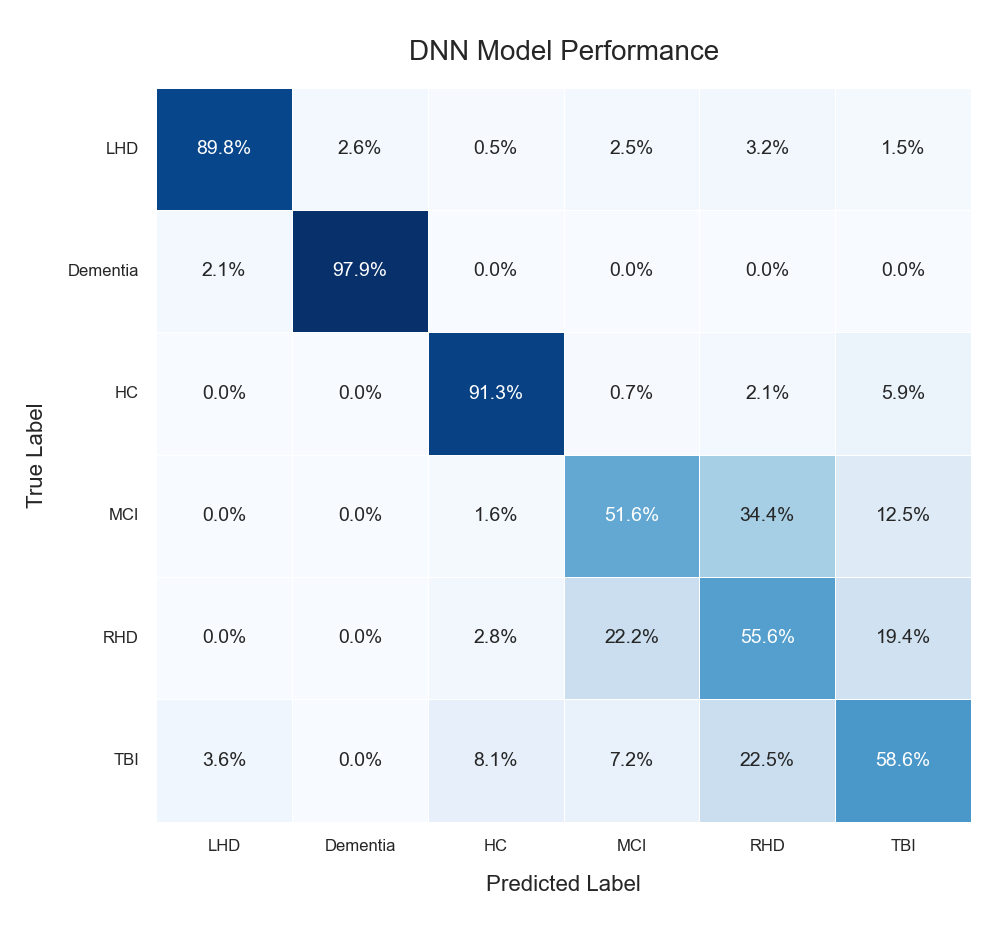

Figure 2 Confusion matrix for Deep Neural Network (DNN) model performance in neurological diagnosis classification. The matrix displays classification accuracy across six diagnostic categories: Left Hemisphere Damage (LHD), Dementia, Healthy Controls (HC), Mild Cognitive Impairment (MCI), Right Hemisphere Damage (RHD), and Traumatic Brain Injury (TBI). True labels are shown on the y-axis and predicted labels on the x-axis, with percentages representing the proportion of cases classified into each category. Diagonal elements (dark blue) indicate correct classifications, while off-diagonal elements represent misclassifications.

The model demonstrates excellent performance for certain conditions: Dementia achieves 97.9% accuracy with minimal misclassification (2.1% confused with LHD), and Healthy Controls achieve 91.3% accuracy. LHD shows robust performance at 89.8% accuracy, with most errors distributed across other neurological conditions. More challenging classifications include MCI (61.6% accuracy) and RHD (55.6% accuracy), which show greater confusion with other conditions, particularly TBI. The pattern of misclassifications reflects the clinical reality of overlapping symptoms between certain neurological conditions, with the model capturing both distinct diagnostic signatures and areas of diagnostic uncertainty that mirror clinical practice. Overall classification performance demonstrates the feasibility of automated neurological screening using quantitative linguistic analysis.

Table 12

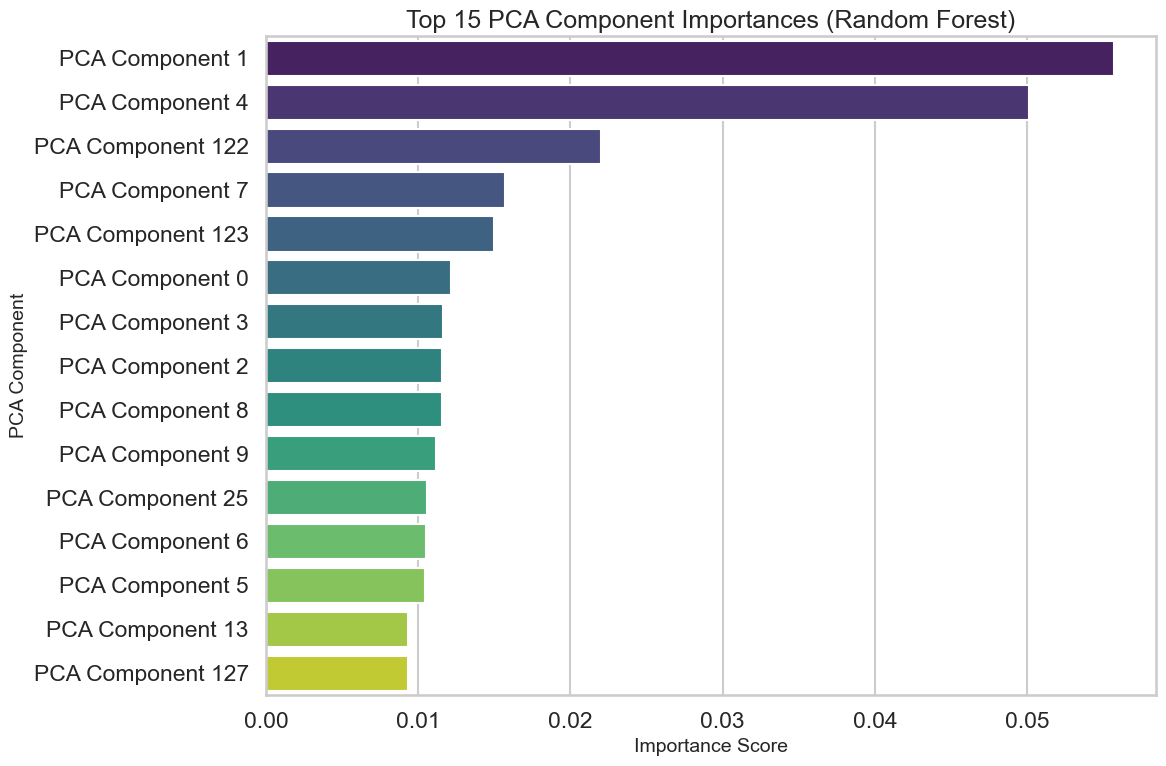

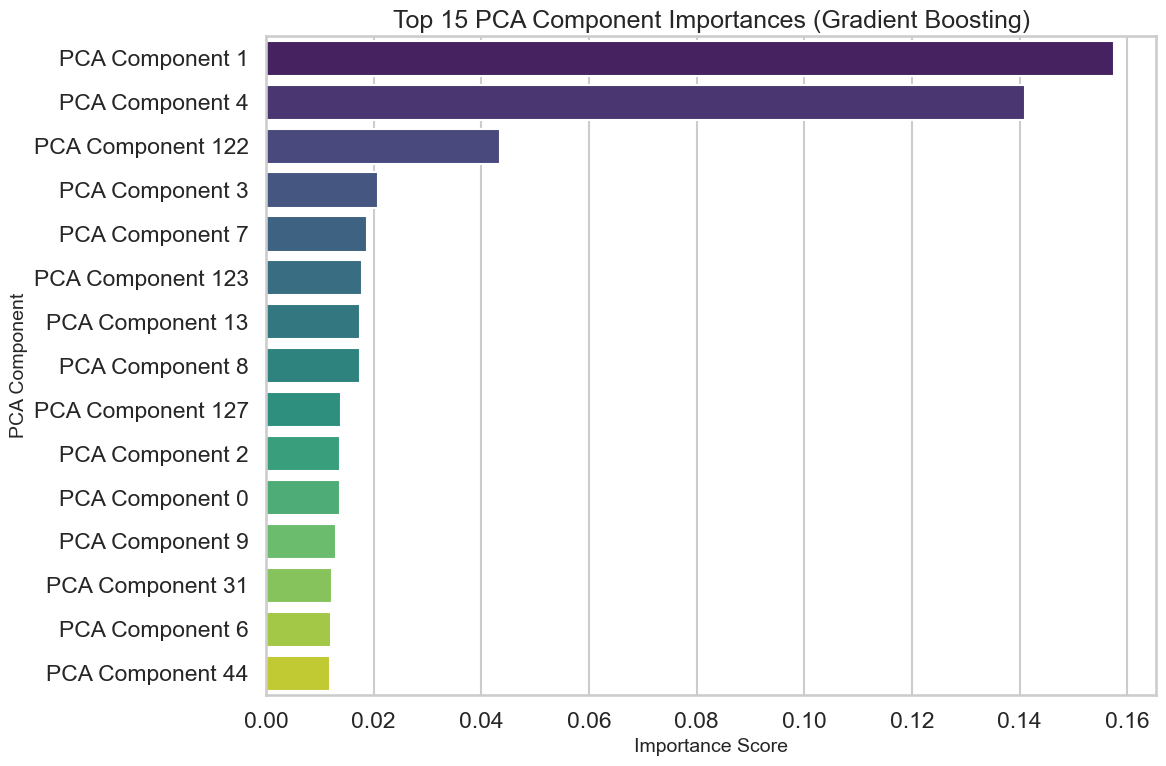
