## Supplementary material for "Automatic screening and characterization of patients with acquired neurological conditions from language": SD2

**Supplementary Data 1**

Table 1. Breakdown of Data Count by Group, Project, and Task.

|  | Diagnosis | Project | Task | Sessions |
| --- | --- | --- | --- | --- |
| 1 | Healthy Controls | NeuralHC | Cat | 358 |
| 2 | Healthy Controls | NeuralHC | Cinderella | 355 |
| 3 | Healthy Controls | NeuralHC | Cookie | 38 |
| 4 | Healthy Controls | NeuralHC | Flood | 88 |
| 5 | Healthy Controls | NeuralHC | Illness | 219 |
| 6 | Healthy Controls | NeuralHC | ImportantEvent | 174 |
| 7 | Healthy Controls | NeuralHC | Sandwich | 343 |
| 8 | Healthy Controls | NeuralHC | Speech | 18 |
| 9 | Healthy Controls | NeuralHC | Umbrella | 331 |
| 10 | Healthy Controls | NeuralHC | Window | 333 |
| 11 | Healthy Controls | Pitt | Cookie | 549 |
| 12 | Aphasia | AphasiaBank | Cat | 666 |
| 13 | Aphasia | AphasiaBank | Cinderella | 988 |
| 14 | Aphasia | AphasiaBank | Flood | 138 |
| 15 | Aphasia | AphasiaBank | ImportantEvent | 584 |
| 16 | Aphasia | AphasiaBank | Sandwich | 566 |
| 17 | Aphasia | AphasiaBank | Speech | 620 |
| 18 | Aphasia | AphasiaBank | Stroke | 589 |
| 19 | Aphasia | AphasiaBank | Umbrella | 631 |
| 20 | Aphasia | AphasiaBank | Window | 996 |
| 21 | Dementia | Pitt | Cookie | 306 |
| 22 | MCI | MCI_DementiaBank | Cat | 71 |
| 23 | MCI | MCI_DementiaBank | Cinderella | 69 |
| 24 | MCI | MCI_DementiaBank | Cookie | 62 |
| 25 | MCI | MCI_DementiaBank | Rockwell | 62 |
| 26 | MCI | MCI_DementiaBank | Sandwich | 68 |
| 27 | MCI | MCI_DementiaBank | Umbrella | 9 |
| 28 | MCI | MCI_DementiaBank | Window | 9 |
| 29 | Right Hemisphere Stroke | RHDBank | Cat | 36 |
| 30 | Right Hemisphere Stroke | RHDBank | Cinderella | 37 |
| 31 | Right Hemisphere Stroke | RHDBank | Cookie | 36 |
| 32 | Right Hemisphere Stroke | RHDBank | Sandwich | 36 |
| 33 | Right Hemisphere Stroke | RHDBank | Speech | 33 |
| 34 | Right Hemisphere Stroke | RHDBank | Stroke | 35 |
| 35 | TBI | TBIBank | Brain_Injury | 57 |
| 36 | TBI | TBIBank | Cat | 54 |
| 37 | TBI | TBIBank | Cinderella | 55 |
| 38 | TBI | TBIBank | ImportantEvent | 55 |
| 39 | TBI | TBIBank | Recovery | 58 |
| 40 | TBI | TBIBank | Sandwich | 56 |
| 41 | TBI | TBIBank | Speech | 58 |
| 42 | TBI | TBIBank | Umbrella | 54 |
| 43 | TBI | TBIBank | Window | 55 |
| Sum |  |  |  | 9955 |
