## Supplementary material for "Automatic screening and characterization of patients with acquired neurological conditions from language": SD1

### Supplementary Table 1

The measures elicited from the picture description tasks are provided below, with a working definition

#### Readability Measures

1. ***Estimated Reading Time (sec)***: Predicts time to read a text, typically calculated by dividing word count by average reading speed (e.g., 200-300 words/minute) and converting to seconds
2. ***Flesch Reading Ease***: Scores text 0–100 based on sentence length and syllables per word Higher scores mean easier reading (e.g., 90-100 = ~5th grade)
3. ***Flesch-Kincaid Grade Level***: Converts Flesch scores to U S grade levels (e.g., 8 0 = 8th-grade comprehension), using sentence length and syllables
4. ***Gunning Fog Index***: Estimates education level needed via sentence length and complex words (≥3 syllables) A score of 12 = 12th-grade level
5. ***Coleman-Liau Index***: Uses characters per word and sentences per one hundred words to determine grade level, avoiding syllable counts
6. ***Automated Readability Index (ARI)***: Relies on characters per word and sentence length to output grade levels (e.g., 10 = 10th grade)
7. ***SMOG Index***: Focuses on polysyllabic words (≥3 syllables) to gauge education level required, often used in healthcare contexts
8. ***Linsear Write Formula***: Tailored for technical writing; weights "hard" words (≥3 syllables) and sentence length, adjusting results for grade level
9. ***Passive Sentences Percent***: Tracks passive voice usage; lower percentages enhance clarity and readability
10. ***Dale-Chall Readability Score***: Uses a predefined "easy word" list and sentence length to assign grade levels (e.g., 9-10 = college level)
11. ***Difficult* *Words***: Measures percentage of complex terms (e.g., multisyllabic or rare words), with higher percentages indicating denser text

#### Phonological Measures

1. Syllable counts count of one, two, three, etc syllable words
2. Structure of syllables: a combination of Consonant (C) and Vowel (V)

#### Morphological Measures

Part of Speech Measures

1. *Adjectives:* words that describe or modify nouns or pronouns (e.g., "red," "happy")
2. *Adpositions:* words that indicate direction, location, or time (e.g., "in," "on")
3. *Adverbs:* words that modify verbs, adjectives, or other adverbs (e.g., "quickly," "very")
4. *Auxiliaries:* helping verbs that express tense, aspect, or modality (e.g., "is," "will")
5. *Coordinating conjunctions:* words that connect words, phrases, or clauses of equal importance (e.g., "and" "but")
6. *Determiners*: words that specify or limit a noun's reference (e.g., "the," "this")
7. *Interjections*: exclamatory words expressing emotion (e.g., "wow," "oh")
8. *Nouns*: words that represent people, places, things, or ideas (e.g., "dog," "city")
9. *Numerals*: words that quantify nouns or indicate order (e.g., "one," "first")
10. *Particles*: function words that modify the meaning of a sentence or verb (e.g., "not," "to")
11. Pronouns: words that replace nouns (e.g., "he," "they")
12. Proper nouns: words that name unique entities (e.g., "John," "Paris")
13. Subordinating conjunctions: words that connect dependent clauses to independent clauses (e.g., "because" "although")
14. Symbols: characters representing symbols (e.g., "$," "%")
15. Verbs: words that express actions, occurrences, or states of being (e.g., "run," "is")
16. Other: words that do not fit into any other POS category

#### Syntactic Measures

##### **Core Syntactic Relations**

1. ***Clausal modifier of noun (acl)***: A clause that modifies a noun (e.g., "The book [that I read] was fascinating")
2. ***Adjectival complement (acomp)***: An adjective functioning as the complement of a verb (e.g., "She seems [happy]")
3. ***Adverbial clause modifier (advcl)***: A clause acting as an adverb to modify a verb (e.g., "He left [because it was late]")
4. ***Adverbial modifier (advmod)***: A word or phrase modifying a verb, adjective, or adverb (e.g., "She ran [quickly]")
5. ***Adjectival modifier (amod)***: An adjective directly modifying a noun (e.g., "[Red] apple")
6. ***Appositional modifier (appos)***: A noun phrase providing additional info about another noun (e.g., "My friend [the doctor] is here")
7. ***Attribute (attr)***: A nominal predicate linked to the subject via a copula (e.g., "She is [a teacher]")

##### **Verb-Related Roles**

1. ***Auxiliary: (aux)***: A helping verb (e.g., "She [has] eaten")
2. ***Auxiliary: (passive) (aux: pass****)*: Auxiliary: verb in a passive construction (e.g., "The cake [was] baked")
3. ***Clausal complement (ccomp)***: A clause acting as the direct object of a verb (e.g., "He said [she left]")
4. ***Open clausal complement (xcomp)***: A clausal complement missing an overt subject (e.g., "She wants [to leave]")

##### **Subject/Object Roles**

1. ***Clausal subject (csubj)***: A clause acting as the subject (e.g., "[That he lied] is obvious")
2. ***Nominal subject (nsubj)***: A noun phrase acting as the subject (e.g., "[The cat] sleeps")
3. ***Nominal subject (passive) (nsubj: pass****)*: Subject of a passive verb (e.g., "[The cake] was eaten")
4. ***Direct object (dobj)***: The noun phrase directly receiving the action (e.g., "She ate [the cake]")
5. ***Object of preposition (pobj)***: Noun governed by a preposition (e.g., "in [the house]")

##### **Modifiers & Connectives**

1. ***Case: marker (case)***: A preposition/postposition marking grammatical Case: (e.g., "[in] the box")
2. ***Coordinating conjunction (cc)***: Connects words/clauses (e.g., "tea [and] coffee")
3. ***Compound modifier (compound)***: Multi-word expressions (e.g., "[coffee] cup")
4. ***Conjunct (conj)***: Elements linked by coordination (e.g., "apples [and] oranges")
5. ***Possession modifier (nmod: poss****)*: Marks possession (e.g., "[John's] book")
6. ***Prepositional modifier (prep)***: A prepositional phrase modifying a noun/verb (e.g., "the book [on the table]")

##### **Special Constructions**

1. ***Expletive (expl)***: A placeholder subject with no semantic role (e.g., "[There] is a problem")
2. ***Interjection (intj)***: An exclamation (e.g., "[Wow], that's amazing!")
3. ***Marker (mark)***: Subordinating conjunction (e.g., "She said [that] it’s true")
4. ***Negation modifier (neg)***: Negation word (e.g., "She [did not] go")
5. ***Parataxis***: Loosely connected clauses (e.g., "He left, [she stayed]")
6. ***Relative clause modifier (relcl)***: Clause introduced by a relative pronoun (e.g., "The man [who called]")

##### **Phrasal Categories**

1. ***Noun Phrases (NP)***: A phrase headed by a noun (e.g., "[The quick brown fox]")
2. ***Verb Phrases (VP)***: A phrase headed by a verb (e.g., "[jumps over the dog]")
3. ***Prepositional Phrases (PP)***: A phrase headed by a preposition (e.g., "[in the house]")
4. ***Adjective Phrases (ADJP)***: A phrase headed by an adjective (e.g., "[very happy]")
5. ***Adverbial Phrases (ADVP)***: A phrase headed by an adverb (e.g., "[quite slowly]")

##### **Miscellaneous**

1. ***Determiner (det)***: Articles or quantifiers (e.g., "[The] cat")
2. ***Number: modifier (nummod)***: Numerals modifying nouns (e.g., "[three] books")
3. ***Particle (prt)***: Verb particle in phrasal verbs (e.g., "give [up]")
4. ***Punctuation (punct)***: Marks like commas or periods
5. ***Root***: The main predicate of the sentence (e.g., the central verb)
6. ***Unclassified dependent (dep)***: A dependency that defies standard classification

Basic Syntax Measures

1. *Total Sentences*: The Number: of sentences in a text
2. *Total Words*: The total word count (tokens) in a text
3. *Average Sentence Length (tokens)*: Mean Number: of words per sentence (Total Words / Total Sentences)
4. *Minimum Sentence Length (tokens)*: Shortest sentence in the text (by word count)
5. *Maximum Sentence Length (tokens)*: Longest sentence in the text (by word count)

Syntactic Tree Metrics

1. *Average Tree Height*: Mean depth of syntactic parse trees (hierarchical structure) across sentences. Higher values indicate more embedded/clauses
2. *Total Tree Height*: Sum of the heights of all syntactic parse trees in the text (if analyzing multiple sentences)
3. *Yngve Depth*: A measure of syntactic complexity based on how deeply phrases are nested in a parse tree. For example, in *"The cat [on the mat [in the room]]"*, the second prepositional phrase increases depth, we provide the following:
   a. *Mean Yngve Depth*: Average Yngve score per sentence, measuring syntactic complexity via right-branching structures (higher = more nested phrases)
   b. *Max Yngve Depth*: Highest Yngve score in the text, indicating the most complex sentence’s nesting depth

Clause and Sentence Complexity

1. *Total Clauses*: Count of all independent and dependent clauses (e.g., *"She laughed [when he fell]"*= 2 clauses)
2. *Total T-units*: *T-units* are used to assess writing maturity; longer/more complex T-units suggest advanced syntax. We calculate the Number: of "minimal terminable units"—a main clause plus any attached subordinate clauses. Measures sentence segmentation (e.g., *"He ran [because he was late]"* = 1 T-unit)
3. *Total Complex T-units*: T-units containing at least one subordinate clause (e.g., *"She said [that it was true]"*)
4. Total Dependent Clauses: Clauses that cannot stand alone (e.g., "[After she left], it rained")

Phrase-Level Metrics

1. Total Verb Phrases (VPs): Phrases centered around a verb (e.g., "[is running quickly]")
2. *Total Coordinate Phrases*: Phrases joined by coordinating conjunctions (e.g., *"apples [and] oranges"*)
3. *Total Complex Nominals*: Noun phrases with modifiers (e.g., *"the [quick brown] fox"*) Counts nouns with adjectives, prepositional phrases, or clauses. Frequent use correlates with formal/academic writing

##### **Semantics**

##### **Numerical and Quantitative Entities**

1. ***Cardinal Number:*** Numerals (e.g., "three", "100")
2. ***Ordinal Number:*** Positions in a sequence (e.g., "first", "third")
3. ***Percentage***: Values with a percent symbol (e.g., "75%")
4. ***Monetary value***: Currency amounts (e.g., "$50", "€200")
5. ***Quantity***: Measurements with units (e.g., "5 kilograms", "10 liters")

##### **Time-Related Entities**

1. ***Date***: Absolute or relative dates (e.g., "January 1, 2023", "next Monday")
2. ***Time***: Time of day or duration (e.g., "3:00 PM", "two hours")

##### **Geographical and Political Entities**

1. ***Location***: Physical places (natural or human-made) (e.g., "Mount Everest", "Pacific Ocean")
2. ***Geopolitical entity (GPE)***: Countries, cities, states, or regions (e.g., "France", "New York")
3. ***Facility***: Man-made structures (e.g., "Kennedy Airport", "Eiffel Tower")

##### **Organizations and Groups**

1. ***Organization***: Companies, institutions, or groups (e.g., "Google", "United Nations")
2. ***Nationalities/Religious/Political groups***: Ethnicities, faiths, or parties (e.g., "Christians", "Democrats")

##### **People and Cultural Entities**

1. ***Person***: Individual names (e.g., "Marie Curie", "Elon Musk")
2. ***Language***: Named languages (e.g., "Mandarin", "Swahili")
3. ***Work of art***: Creative works (e.g., "Mona Lisa", "Hamlet")

##### **Events and Abstract Concepts**

1. ***Event***: Historical, cultural, or planned occurrences (e.g., "World War II", "Olympic Games")
2. ***Law***: Legal documents, statutes, or treaties (e.g., "Civil Rights Act", "GDPR")

##### **Commercial Entities**

1. ***Product***: Branded goods or services (e.g., "iPhone", "Tesla Model S")

### Lexical Measures

Automated.Lexical.Density..Content.Words...Total.Words.

Average.Word.Length..in.Characters.

Content.Words..Total.

Content.Words..Unique.

Corrected.TTR..CTTR.

Function.Words..Total.

Function.Words..Unique.

Maas.s.TTR..A2.

Total.Words

Type.Token.Ratio..TTR.

Types

Words..Tokens.
