## Supplementary material for "Automatic screening and characterization of patients with acquired neurological conditions from language": SD3

### Supplementary Data: Model Performance

Table 1. Performance metrics for machine learning models in neurological diagnosis classification. Five machine learning algorithms were evaluated on an independent test set using six standard classification metrics. Performance is reported as percentages for Accuracy, Balanced Accuracy, F1-Score, and Cohen's Kappa, and as decimal values converted to percentages for AUC-ROC and AUC-PR. Accuracy represents overall classification performance; Balanced Accuracy adjusts for class imbalance by averaging recall across all classes; F1-Score provides the harmonic mean of precision and recall; Cohen's Kappa measures agreement beyond chance; AUC-ROC quantifies the area under the receiver operating characteristic curve; AUC-PR represents the area under the precision-recall curve. Support Vector Machine (SVM) achieved the highest overall accuracy (91%) and Cohen's Kappa (84%), while maintaining excellent AUC scores (97% ROC, 97% PR). Deep Neural Network (DNN) demonstrated the most balanced performance profile with strong metrics across all categories. Logistic Regression showed robust performance with the second-highest accuracy (89%) and Cohen's Kappa (82%). Random Forest exhibited the lowest performance, particularly in Balanced Accuracy (50%), indicating difficulty with class imbalance. All models achieved AUC-ROC scores above 90%, demonstrating strong discriminative ability for neurological condition classification based on linguistic features.

| **Model** | **Accuracy** | **Balanced Accuracy** | **F1-Score** | **Cohen's Kappa** | **AUC-ROC** | **AUC-PR** |
| --- | --- | --- | --- | --- | --- | --- |
| DNN | 87% | 74% | 89% | 78% | 97% | 94% |
| Logistic Regression | 89% | 72% | 90% | 82% | 92% | 91% |
| SVM | 91% | 68% | 91% | 84% | 97% | 97% |
| Gradient Boosting | 84% | 58% | 84% | 73% | 92% | 92% |
| Random Forest | 83% | 50% | 81% | 69% | 90% | 90% |
